# ARCliDS: A Clinical Decision Support System for AI-assisted Decision-Making in Response-Adaptive Radiotherapy

**DOI:** 10.1101/2022.09.23.22280215

**Authors:** Dipesh Niraula, Wenbo Sun, Jionghua (Judy) Jin, Ivo D. Dinov, Kyle Cuneo, Jamalina Jamaluddin, Martha M. Matuszak, Yi Luo, Theodore S. Lawrence, Shruti Jolly, Randall K. Ten Haken, Issam El Naqa

## Abstract

**Background:** Involvement of many variables, uncertainty in treatment response, and inter-patient heterogeneity challenge objective decision-making in dynamic treatment regime (DTR) in oncology. Advanced machine learning analytics in conjunction with information-rich dense multi-omics data have the ability to overcome such challenges. We have developed a comprehensive artificial intelligence (AI)-based optimal decision-making framework for assisting oncologists in DTR. In this work, we demonstrate the proposed framework to Knowledge Based Response-Adaptive Radiotherapy (KBR-ART) applications by developing an interactive software tool entitled **A**daptive **R**adiotherapy **Cli**nical **D**ecision **S**upport (ARCliDS).

**Methods:** ARCliDS is composed of two main components: Artificial RT Environment (ARTE) and Optimal Decision Maker (ODM). ARTE is designed as a Markov decision process and modeled via supervised learning. Given a patient’s pre- and during-treatment information, ARTE can estimate treatment outcomes for a selected daily dosage value (radiation fraction size). ODM is formulated using reinforcement learning and is trained on ARTE. ODM can recommend optimal daily dosage adjustments to maximize the tumor local control probability and minimize the side effects. Graph Neural Network (GNN) is applied to exploit the inter-feature relationships for improved modeling performance and a novel double GNN architecture is designed to avoid unphysical treatment response. Datasets of size 117 and 292 were available from two clinical trials on adaptive RT in non-small cell lung cancer (NSCLC) patients and adaptive stereotactic body RT (SBRT) in hepatocellular carcinoma (HCC) patients, respectively. For training and validation, dense data with 297 features were available for 67 NSCLC patients and 110 features for 71 HCC patients. To increase the sample size for ODM training, we applied Generative Adversarial Network to generate 10,000 synthetic patients. The ODM was trained on the synthetic patients and validated on the original dataset.

**Results:** Double GNN architecture was able to correct the unphysical dose-response trend and improve ARCliDS recommendation. The average root mean squared difference (RMSD) between ARCliDS recommendation and reported clinical decisions using double GNNs were 0.61 ± 0.03 Gy/frac (mean±sem) for adaptive RT in NSCLC patients and 2.96 ± 0.42 Gy/frac for adaptive SBRT HCC compared to the single GNN’s RMSDs of 0.97 ± 0.12 Gy/frac and 4.75 ± 0.16 Gy/frac, respectively. Overall, For NSCLC and HCC, ARCliDS with double GNNs was able to reproduce 36% and 50% of the good clinical decisions (local control and no side effects) and improve 74% and 30% of the bad clinical decisions, respectively.

**Conclusion:** ARCliDS is the first web-based software dedicated to assist KBR-ART with multi-omics data. ARCliDS can learn from the reported clinical decisions and facilitate AI-assisted clinical decision-making for improving the outcomes in DTR.

## Introduction

Optimal decision-making in Knowledge Based Response-Adaptive Radiotherapy (KBR-ART) is a difficult task^1^. The difficulties arise from a slew of factors, such as, involvement of many variables, uncertainty in treatment response, and inter-patient heterogeneity ^2^. In the absence of a quantitative framework, clinical decisions are primarily influenced by physician’s professional experiences, which may result in inter-physician variability. Thus, there is a need for a robust and user-friendly clinical decision-support tool for objective decision-making in KBR-ART that is data-driven and consistent^3^.

Adaptive Radiotherapy Clinical Decision Support (ARCliDS) is a web-based software tool for AI-assisted optimal decision-making in KBR-ART^4–6^ and potentially other oncology applications involving dynamic treatment regime (DTR)^7,8^. ARCliDS provides a quantitative approach to overcome the decision-making difficulties via a set of data analytics algorithms, which include feature selection of important variables, statistical ensemble for representing uncertainties of treatment response, and, most importantly, integration of information-rich dense multi-omics datasets for capturing inter-patient heterogeneity^9–11^. ARCliDS combines all the above data analytics capabilities and presents a user-friendly interface for evaluating relevant clinical use cases. Moreover, it is complementary to the current treatment planning system; the integration may facilitate an introduction of multi-omics information into the treatment planning workflow.

DTR including adaptive RT (ART)^12^ are designed for treatment personalization. A popular ART paradigm and implementation is to adapt treatment plans to accommodate during-treatment anatomical changes due to weight loss, tumor regression and/or diminution of the volume of surrounding normal tissue and organ at risk (OAR). A complementary ART paradigm is KBR-ART which provides a response-based adaptive framework for personalizing RT as shown in Figure 1, where the response assessment is not limited to observing anatomical changes. It is divided into three phases: *Pre-Treatment Assessment, Treatment Response Evaluation* (evaluation phase) and *Treatment Adaptation* (adaptation phase). In the pre-treatment phase, a patient’s disease and condition is assessed and a treatment plan is tailored. In the evaluation phase, a patient’s treatment response is evaluated by comparing pre and mid treatment multi-omics information changes. Based on the treatment responses, the patient’s associated outcome probabilities are estimated. In the adaptation phase, treatment planning is adapted for a personalized and an optimal outcome. Two endpoints are considered: tumor control and normal tissue complication. The goal of KBR-ART is to maximize tumor control probability (TCP) and minimize normal tissue complication probability (NTCP).

**Figure 1:**
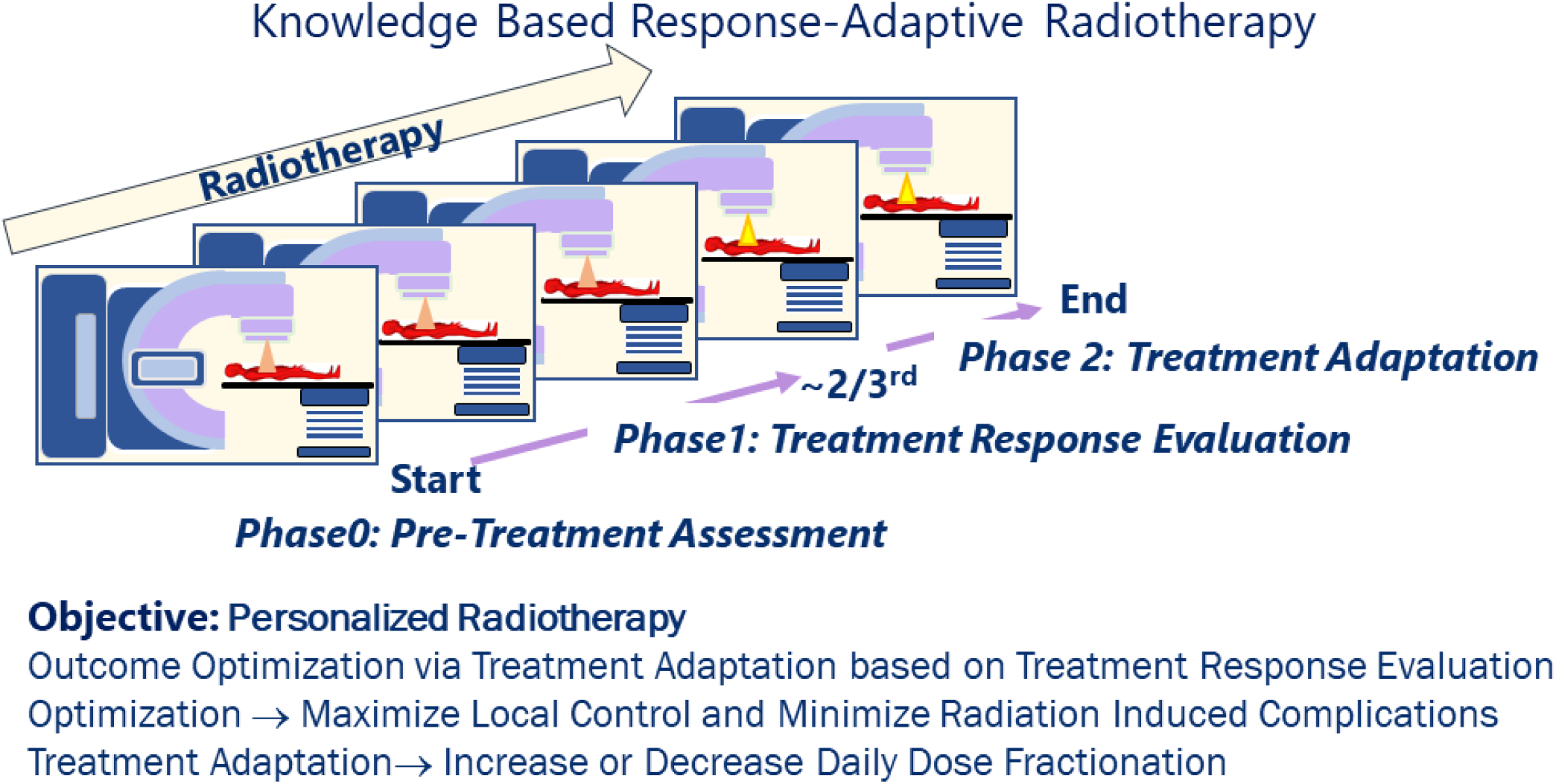
Knowledge Based Response-Adaptive Radiotherapy (KBR-ART). In KBR-ART, a pre-treatment assessment is conducted in phase 0 and appropriate treatment plan is tailored. Then patients’ treatment response is evaluated in Phase 1, and an optimal treatment adaptation is planned and executed in Phase 2.

To demonstrate the potential of ARCliDS, two clinical use cases are presented. In both studies, the evaluation time was around 1 month. The first use case is based on the UMCC (University of Michigan Cancer Center) 2007-123 phase II dose escalation clinical trial NCT01190527 ^13^, where inoperable or unresectable non-small cell lung cancer (NSCLC) patients were administered with 30 daily dose fractions. The patients received roughly 50 Gy [Gray = J/Kg] equivalent dose in 2 Gy fractions (EQD2) in the evaluation phase and up to a total dose of 92 Gy EQD2 in the adaptation phase. The evaluation phase lasted for roughly two-thirds of the 6-week treatment period. In the second clinical use case, patients with hepatocellular carcinoma (HCC) received adaptive SBRT in clinical trials NCT01519219, NCT01522937, and NCT02460835^14^. In the evaluation phase, patients received 3 daily dose fractions followed by 1 month break, and in the adaptation phase, a suitable sub-population of the patients received 2 additional daily doses.

A large sample size that is representative of the true population is preferred for all data driven and statistical modeling. However, due to financial, feasibility, and ethical reasons, obtaining a large dataset in medical field is often impractical. In our case, a dataset of size 117 and 292 were available for NSCLC patients and HCC patients, respectively. Dense multi-omics data with 297 features were available for only 67 NSCLC patients and 110 features for 71 HCC patients. These datasets, albeit on the smaller size, are unique as KBR-ART is still in its clinical trial phase and hence the largest multi-omics datasets for KBR-ART. So, although the sample size looks small, the information-rich dense multi-omics dataset is the largest of its kind.

Under the current United States Food and Drug Administration (FDA) definition and guidelines, ARCliDS is categorized as a Software as a Medical Device (SaMD)^6^. SaMD is defined as software intended to be used for medical purposes independently in contrast to software intended to drive a hardware medical device (software in a medical device). This definition was recently adopted by FDA to include AI software^15,16^ which can automatically learn from user cases and continuously update after deployment, as opposed to traditional software, which stays fixed after deployment (excluding version update). Therefore, ARCliDS also has two modes of operation: *Operation Mode* and *Learning Mode* as shown in Figure 2. After the initial training, both modes can run simultaneously (online learning) in the clinic.

**Figure 2:**
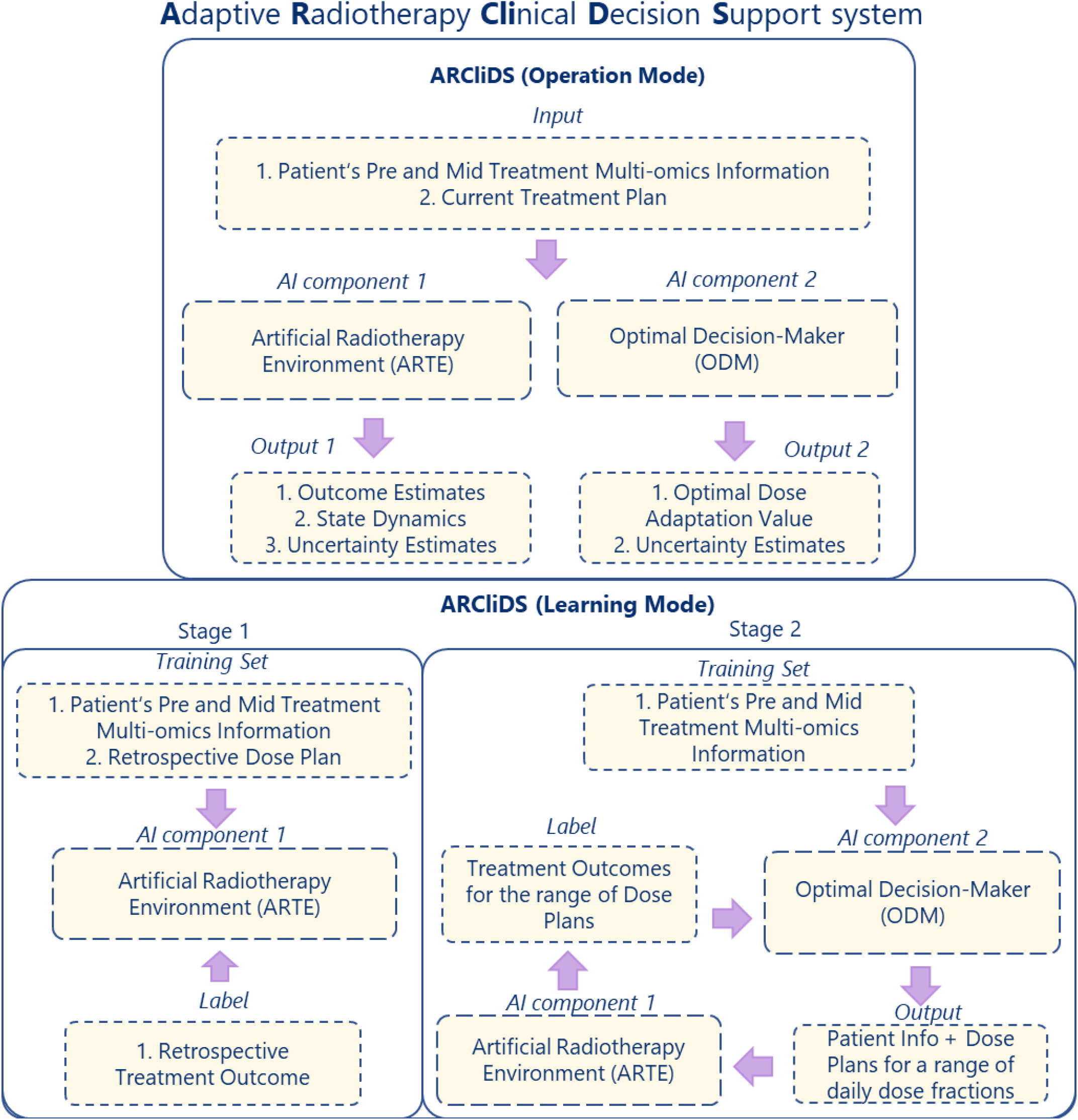
ARCliDS Blueprint. ARCliDS is composed of two AI components: (1) Artificial Radiotherapy Environment (ARTE) and (2) Optimal Decision-Maker (ODM). ARCLiDS learns ARTE via supervised learning. ARTE is then utilized in planning and teaching ODM via reinforcement learning. In the operation mode, ARTE outputs State Dynamics and RT Outcome estimates while ODM outputs the optimal dose adaptation recommendation. Both ARTE and ODM present uncertainty estimates.

ARCliDS is composed of two main AI components. The first component is the Artificial Radiotherapy Environment (ARTE) for estimating the predicted outcome and the second component is the Optimal Decision-Maker (ODM) for decision-making. In Operation Mode, ARCliDS asks for a patient’s pre and mid treatment multi-omics information, and current treatment plan. It feeds that information into ARTE and ODM, and obtains outcome estimates, state dynamics, and the optimal dose adaptation value. All of the estimated results come with associated uncertainty. The results are presented in two main plots: outcome space spanned by TCP and NTCP, and population distribution plots as further explained in the Graphical User Interface (GUI). During the Learning Mode, ARTE is trained first on the available data, and then ODM is trained on the ARTE. The details of the training are presented in the Methods section and SM.

ARCliDS presents a significant improvement to Tseng et al.’s ^17^ and Niraula et al.’s ^5^ methods. The improvement comes from the graphical representation of patients’ features. Convolution neural networks (CNNs) are known to perform well because they exploit the feature locality of images^18^. In other words, pixels at neighboring areas of an image are correlated, and CNN architectures can capture those correlations. Graph Neural Networks (GNNs) are similar except they exploit the non-local relationship between feature values^19^. Computationally, GNNs use fewer network connections compared to fully connected NNs, which help in learning by reducing redundancies. From another perspective, information from one feature only goes to its neighboring features. In this work, we have borrowed the feature graph from Luo et al.’s work on multi-objective Bayesian Network^20^ which identified the most important features related to RT outcome of interest by finding the Markov Blanket of the outcomes. Details of feature selection procedure are presented in Supplementary Materials (SM) section S1. For NSCLC and HCC, we were able to (coincidentally) select 13 important features.

The materials in this manuscript are arranged as follows. We begin by introducing ARCliDS’ GUI. We then present the details of ARTE and ODM in the Methods section. ARTE is further divided into the descriptions of patient state, transition function (TF), and RT outcome estimator (RTOE). We have proposed a linear-quadratic-linear (LQL) type TF. This is an improvement from Niraula et al.’s^5^ work which employed a linear-quadratic (LQ) type function. LQL model is a generalization of LQ model that covers both RT and SBRT. Furthermore, for the RTOE, we applied GNN to general logistic function guided double NN architecture, which was developed by Niraula et al.^5^ To improve the robustness of ODM, we have replaced the online learning approach with the Planning and Learning approach^21^. We trained and validated ARCliDS in two different cancer sites and different treatment types to show its versatility. We conclude the manuscript with a discussion and future steps.

## Methods

### I. Graphical User Interface

We have designed ARCliDS as a Web Application (app) using R Shiny as shown in Figure 3. The app consists of 4 main panels: Data Input Panel, Outcome Space, Population Distribution Plot and Report Print. Beside these, there are accessibility tools such as help information, user guide, documentation, zooming, and printing option.

**Figure 3:**
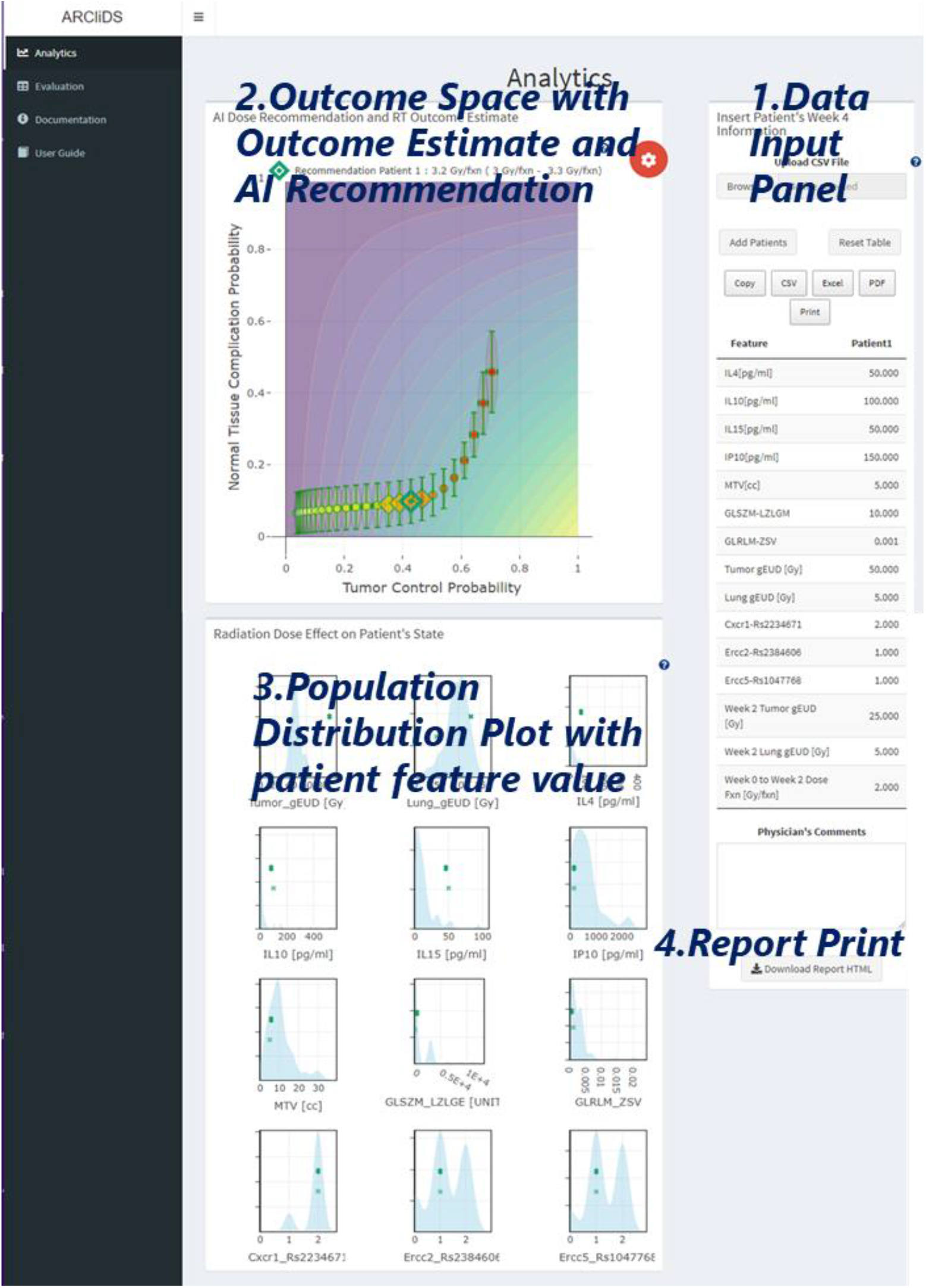
ARCliDS graphical user interface (GUI). The GUI consists of the Data Input Panel, Outcome Space spanned by TCP and NTCP, historic Population Distribution Plots, and Report Print.

#### 1. Data Input Panel

Patient Data can be input manually or via a data file. Multiple patients’ states can be input for visual comparison. The inputs can be saved or printed if necessary. There is a dedicated space for Physician notes.

#### 2. Outcome Space

We present the AI recommendations in the Output Space. The Output Space is spanned by TCP in the x-axis and NTCP in the y-axis. We contoured and colored it with the Reward Function, providing additional insight on the AI’s Decision Making. Given a patient’s information, it shows treatment outcome for a range of daily dose fractions and marks the treatment outcome for the optimal dose recommendation. It provides uncertainty assessment for both the outcome estimate and AI recommendation.

#### 3. Population Distribution Plot

Knowing the patient’s state value and its relative position to the population, provides information on patient’s “whereabouts”. To accommodate a comparison on the feature level, we have included histograms for each feature and patient’s state value atop.

#### 4. Report Print

We have designed a report printing in html format. The interactive nature of the report, even outside the app, makes it much easier to communicate with other users.

### II. Artificial RT Environment (ARTE)

Radiation damages both cancer cells and normal tissue cells. To quantify the relationship between the applied radiation and the treatment response, we consider the radiation absorbed by tumor and surrounding normal tissue, and the probabilities of tumor control (TCP) and normal tissue complication (NTCP).

The absorbed radiation is spatially non-uniform, so it is generally converted to a homogenous dose value by weighted-averaging of the treatment sites from treatment planning. Generalized equivalent uniform dose (gEUD) is one such metric^22^. It is expected that for a fixed radiation site, gEUD must increase with increasing applied radiation as shown in Figure 4. We have assumed a linear-quadratic-linear (LQL)^23^ type monotonic proportionality relationship (S1) as presented in the SM, which further results in the following two relationships for KBR-ART, i.e.,

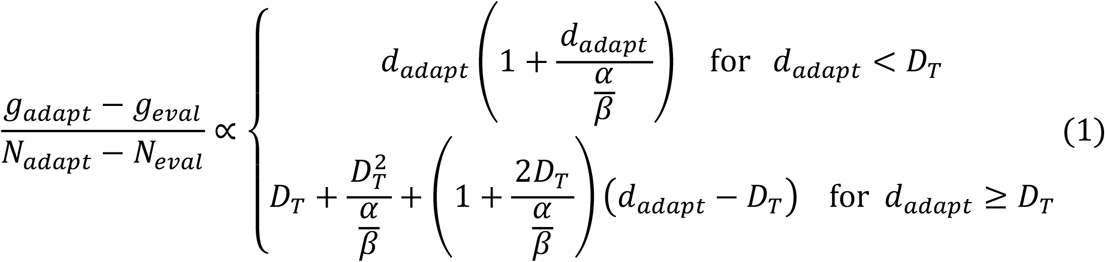

**Figure 4:**
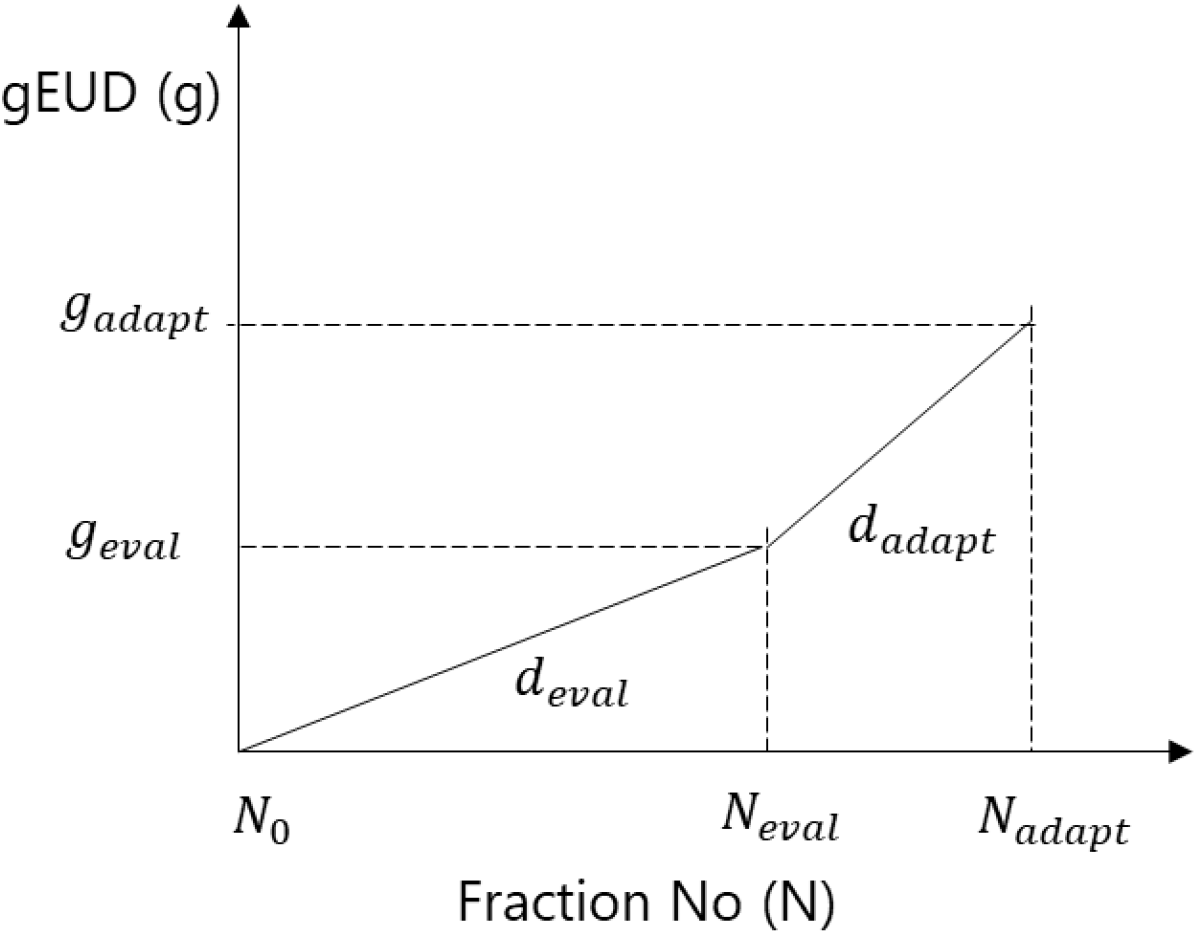
Transition Function for gEUD. Here, the KBR-ART regimen is divided into three time points, pre, mid, and post treatment, denoted by the daily dose fraction, *d*, number *N*_0_, *N*_*eval*_, and *N*_*adapt*_, respectively. The treatment period between *N*_0_ and *N*_*eval*_ is the Evaluation Phase and between *N*_*eval*_ and *N*_*adapt*_ is the Adaptation Phase.

And,

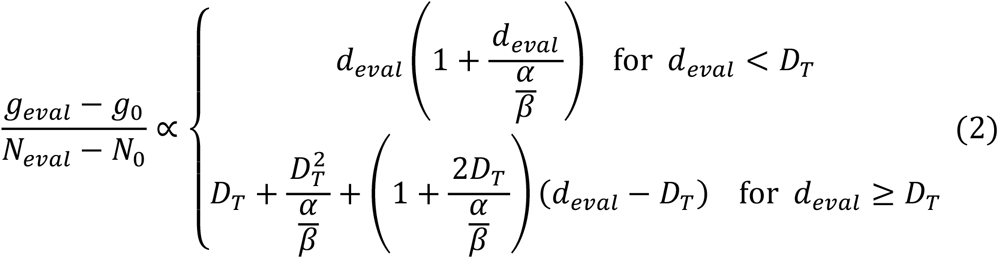

where, *g* stands for gEUD, *N* for *n*th daily dose fractions, *d* for dose fractions, *D*_*T*_ for threshold doses, and *α*/*β* ratio is a tissue-specific parameter. The subscript 0, eval, and adapt of *N* and *g* corresponds to pre-, mid-, and after-treatment, respectively while *d*_*eval*_ and *d*_*adapt*_ corresponds to applied daily dose fractionations during the evaluation phase and adaptive phase, respectively. Dividing the relationships (1) and (2) yields four equations for *g*_*adapt*_ as listed in SM Table S1.

With the assumption that the increment of radiation increases both TCP and NTCP, we have applied a sigmoid shape generalized logistic function to represent the outcome probability as follows,

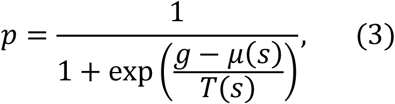

where the patient-specific parameters *μ* and *T* are functions of their multi-omics state. By applying patient’s pre and mid treatment multi-omics information, the above dose-response relationship captures inter-patient heterogeneity.

Applying the equations from Table S1 and Eq. (3), ARTE is built as a Markov Decision Process (MDP) as shown in Figure 5. ARTE takes in patient’s state (*s, c*) and daily dose fractionation (*d*) as the input and returns patient’s next state (*s*’) and outcome (*tcp, ntcp*) as the output. The state dynamics is modeled by the TF and the associated outcome by the RTOE.

**Figure 5:**
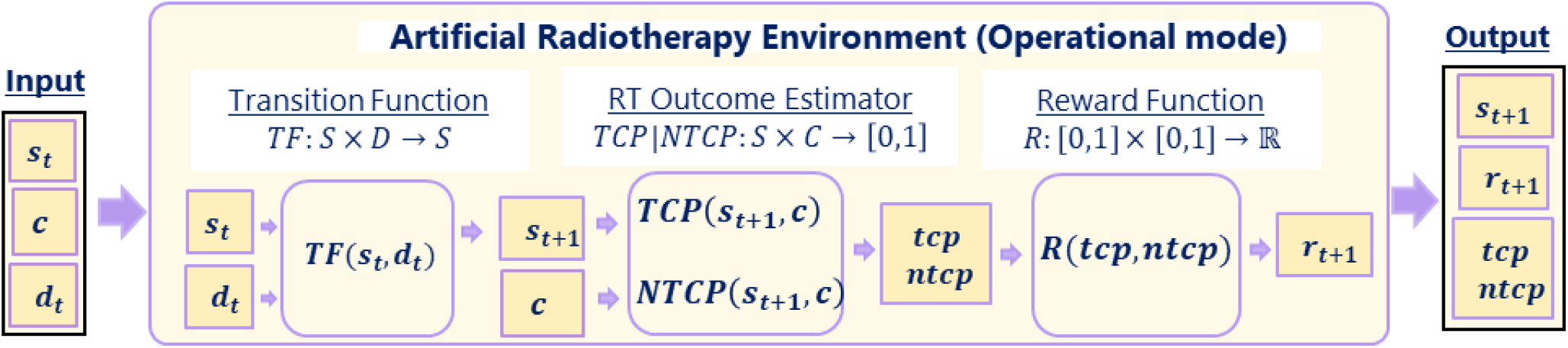
Workflow of Artificial Radiotherapy Environment (ARTE). ARTE is composed of three functions: Transition Function (TF), RT Outcome Estimator (RTOE), and Reward Function. For patients in state *s*_*t*_ that have been administered dose *d*_*t*_, TF predicts the resulting state *s*_*t*+1_. RTOE estimates *tcp* and *ntcp* for a patient in state *s*_*t*+1_ and covariate *c*. The reward function *R* assigns a reward *r*_*t*+1_ to the tuple (*tcp,ntcp*), so that optimal reward corresponds to maximal *tcp* and minimal *ntcp*. Overall, given *s*_*t*_, *c*, and *d*_*t*_, ARTE yields *s*_*t*+1_, *r*_*t*+1_, *tcp* and *ntcp*.

### A. Patient States

Patient State, *S* ⊂ ℝ^*k*^, represents a patient’s information at a given time. It consists of patient’s features such as dosimetric, clinical, radiomics, genomics, and imaging information as listed in Tables S3 and S10. MDP assumes that patient’s state at time t+1 only depends on patient’s state and dose, *D* ⊂ ℝ, at time t. In KBR-ART, three time points are relevant as shown in Figure 4: (i) pretreatment (t=0), (ii) mid treatment (t = eval), and (iii) post treatment (t = adapt).

Previously, deep regression models^5,17^ were applied to learn state transitions for all the patient features. In this work, however, to reduce modeling error, we consider the non-dosimetric variables at any time points as predictors for the treatment outcome and directly use the clinically measured values. We only estimate the state transition for dosimetric variables since the treatment outcome largely depends on the radiation. Hence, we have divided the patients states into time-varying dosimetric variables, *S* ⊂ ℝ^*m*^, and a set of other fixed multi-omics covariates, *C* ⊂ ℝ^*k*−*m*^. A complete patient’s state is given by (*s, c*) ∈ *S* × *C*.

### B. Transition Function (TF)

*TF: S* × *D* → *S*, predicts the next state, *s*_*t*+ 1_ for patients in state, *s*_*t*_, under given dose, *d*_*t*+ 1_. We have used two TFs for the dosimetric variables, tumor gEUD and normal tissue gEUD. We empirically found that deep model-based TF for gEUDs does not always maintain the causal monotonic relationship. Thus, we applied the well-known LQL based relationship from Eq. (S1) to guarantee an increasing monotone relationship between the dose applied and dose absorbed as presented in Eq. (1) and Eq. (2).

### C. RT Outcome Estimator (RTOE)

RTOEs, *TCP*: *S* × *C* → [0,1] and *NTCP*: *S* × *C* → [0,1], estimate *tcp* and *ntcp* for the patient’s state, (*s*_*t*+ 1_, *c*). In this work, we have applied GNN as the RTOE as shown in Figure 6. Each patient is assigned with a graph of features and then a binary classification is learned on the graph level. We first applied a single GNN for RTOE. While the performance is improved drastically compared to a fully connected classifier, the single GNN was found to not respect the monotonicity between the dose value and the outcome probability. To meet the monotonic relationship, we applied a double GNN architecture with a generalized logistic function named as generalized logistic function guided double GNN (GloGD-GNN).

**Figure 6:**
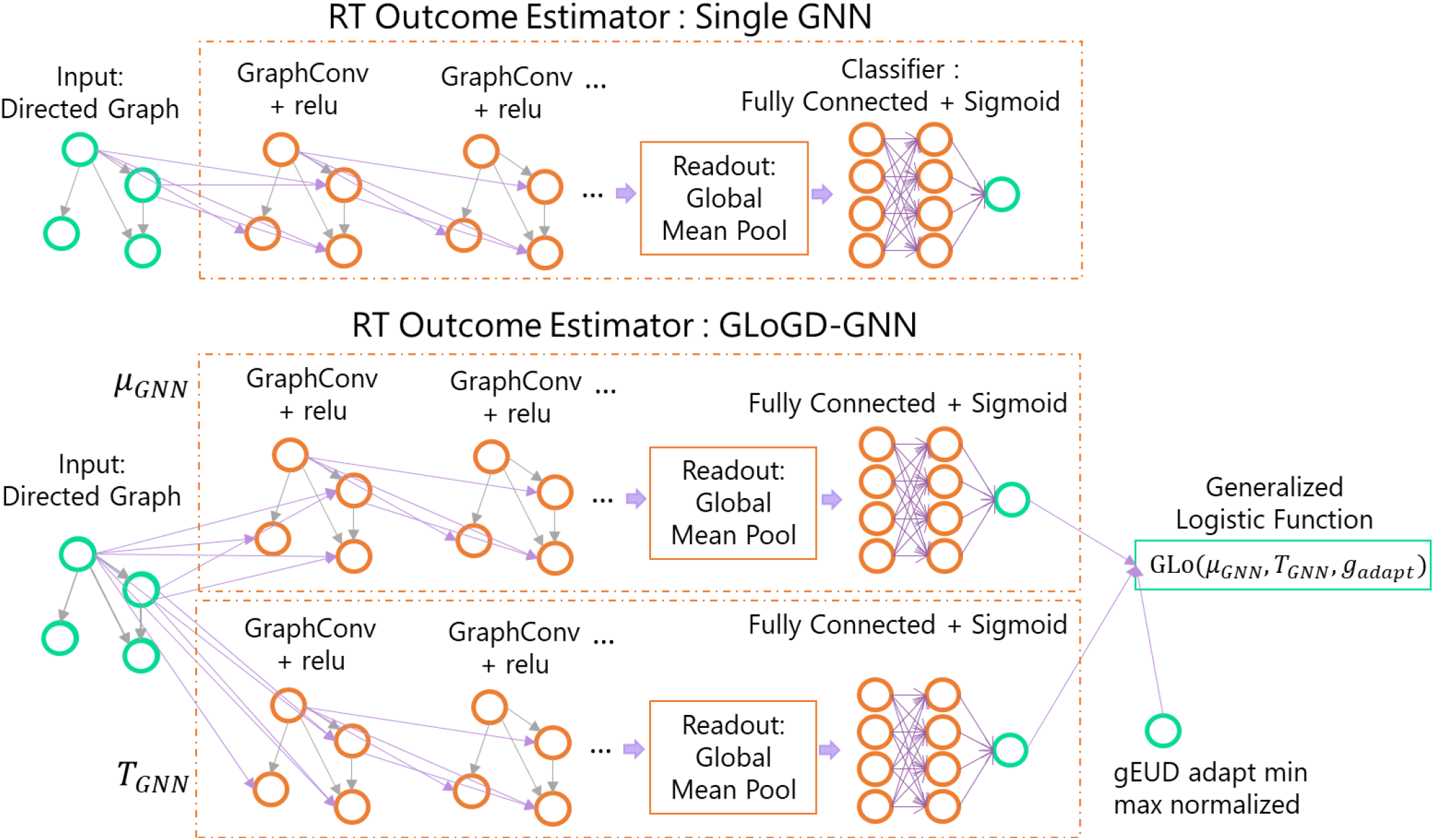
Two GNN based RT Outcome Estimator Architectures. A Single Graph Neural Network (GNN) has an input layer for graph input, graph convolution layers for graph embedding, a global mean pool layer, and a fully connected classifier layer. Generalized logistic function guided double GNN (GLoGD-GNN) has two Single GNNs fed into a 2-parameter logistic function. GLoGD-GNN takes in gEUD as the argument.

### D. Reward Function

Reward function, *R*: [0,1] × [0,1] → ℝ, assigns a value to the (*tcp, ntcp*) pair. The reward function is selected such that its optimization results in maximal *tcp* and minimal *ntcp*. We have adopted, *R* = *tcp*(1 − *ntcp*), reward function for ARCliDS. As seen from Figure 7, it is smallest at the negative outcomes, {(*tcp, ntcp*)} = {(0, 0), (0, 1), (1, 1)}, and largest at the positive outcome, (*tcp, ntcp*) = (1, 0).

**Figure 7:**
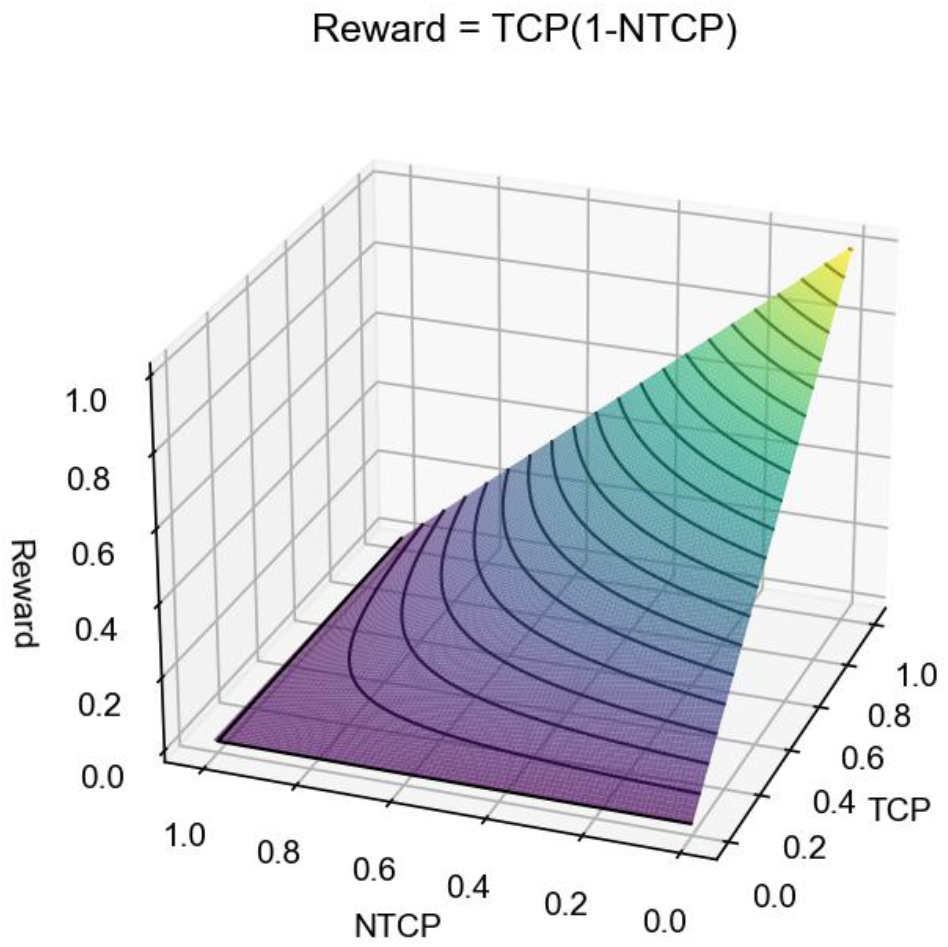
Reward Function 3D Contour Plot. The reward function, *r* = *tcp*(1 − *ntcp*), smoothly raises toward its maximum value at (*tcp, ntcp*) = (1,0). AI’s goal is to find doses that will result in the maximum reward.

Additionally, a goal is defined. By default, the goal can be defined as *tcp* > 50% and *ntcp* < 50%, which rounds to positive outcome. Furthermore, goal based on population endpoints can be added. For NSCLC, a goal of *tcp* > 70% and *ntcp* < 17.2 %, ^17^ and for HCC, *tcp* > 90% and *ntcp* < 25 % is added.

Combining the reward and goal, the reward scheme for NSCLC and HCC is defined as following,

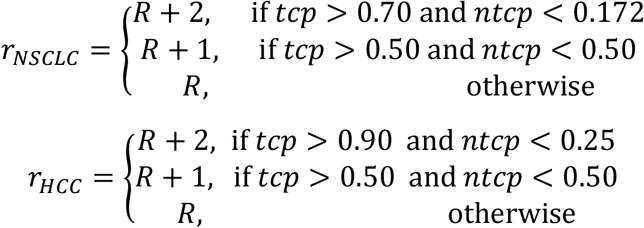

Note that goals might be unattainable for some patients.

### III. Optimal Decision Maker (ODM)

We utilized a deep reinforcement learning algorithm for training the ODM. ODM is composed of a Q (quality) function, *DQN*: *S* × *C* → ℝ^*d*^, and a selector. Given a patient’s state *s*_*t*_, deep Q-net generates a set of q-values, *Q* ⊂ ℝ^*d*^, for a range of dose. Q-Net maps k-dimensional state space to d-dimensional action (dose-decision) space. During operation mode, it simply follows greedy policy and selects the dose 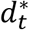 having the maximum q-value. For training, we have adopted a model-based RL paradigm that is divided into two phases: *Planning* and *Learning*, as shown in Figure 8. During Planning, an exhaustive search is carried out where all patient’s states (*s*_*t*_, *c*) and the range of adaptive dose *d*_*t*_ are fed into the ARTE and the resulting states *s*_*t*+1_, and rewards *r*_*t*+1_ are saved into the Memory. During Learning, the DQN is trained via double DQN algorithms^24^ using the Memory as a one-step optimization problem.

**Figure 8:**
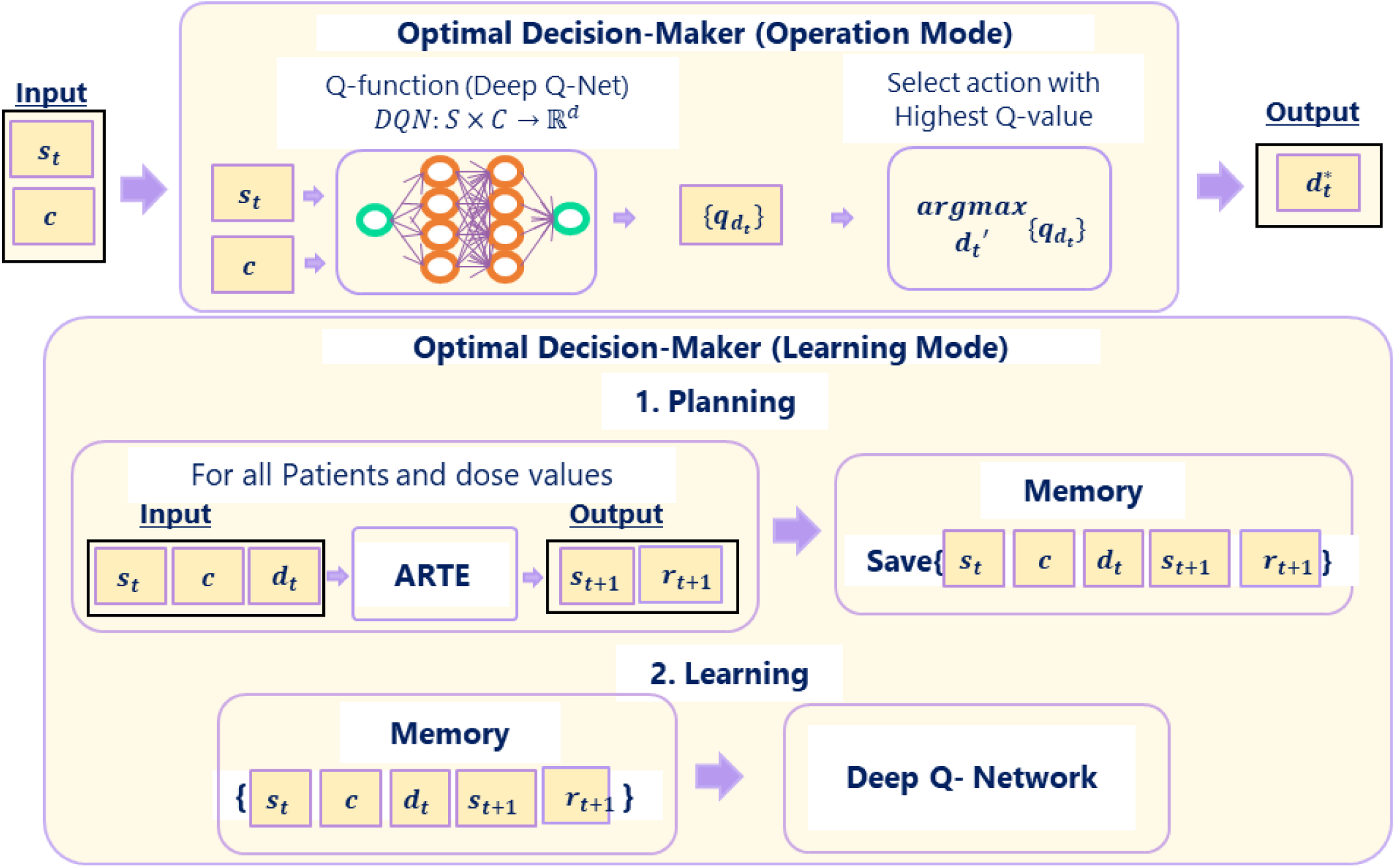
Workflow of Optimal Decision-Maker (ODM). ODM is composed of a deep Q-network (DQN) and decision selector. Given a state (*s*_*t*_, c), DQN yields a q (quality) value for the range of adaptive dose. The selector greedily selects the dose with the highest q-value. The ODM is trained by following the model-based reinforcement learning paradigm. In the Planning Phase, the ODM saves next states {*s*_*t*+1_} and associated rewards {*r*_*t*+1_} for all patient’s state {(*s*_*t*_,*c*)} and the range of adaptive dose {*d*_*t*_}. In the Learning phase, a double DQN algorithm is applied on the memory.

### IV. Uncertainty Estimate via Statistical Ensemble

Model uncertainty is estimated using Statistical Ensembling. The statistical ensemble technique trains several identical models and finds averages and deviations of the prediction. This method estimates uncertainty purely based on the trained model. Additionally, this also helps with desensitizing ARCliDS to the noise associated with the stochastic optimization algorithm used by NNs. NNs utilize a large number of randomly initiated weights and as a result, learned weights are different from model to model^25^. ARCliDS presents the average prediction *μ* as an expected value, and the covariance *cov* as an uncertainty estimation as shown in Figure 9. For ODM, standard deviation σ is used as the uncertainty estimate.

**Figure 9:**
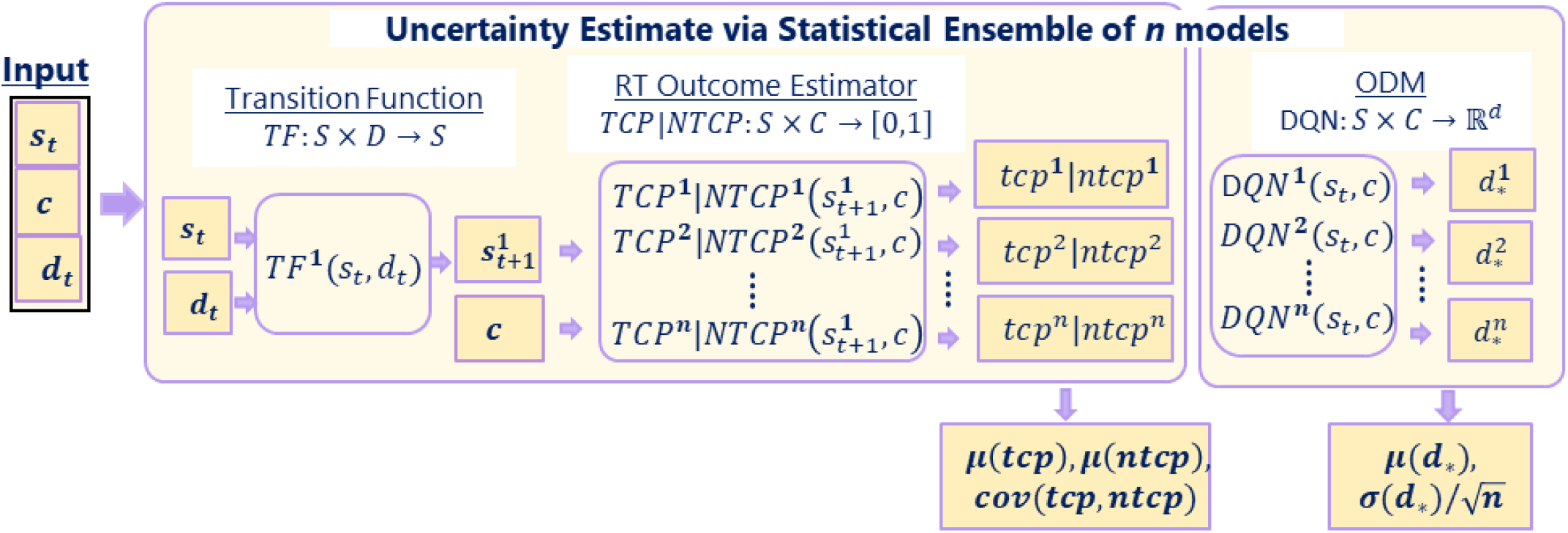
Model Uncertainty via Statistical Ensemble. We trained *n* identical models. The mean and covariance (or standard deviation) of the output distribution captures the model output and model uncertainty. For RTOE’s probability outputs, we present the uncertainty estimates as the covariance and for ODM dose recommendations, we present the uncertainty estimates as the standard error of mean.

### V. Analysis

ARCliDS was trained and validated on two different use cases from two different types of RT treatments. The first use case is an adaptive RT clinical trial of NSCLC patients, and the second use case is an adaptive SBRT clinical trial. After feature selections, we first built five ARTEs for each disease using the dataset. We then generated 10,000 synthetic patients using a generative adversarial network (GAN)^26^. Five ODMs were trained using the five ARTEs and 4,000 randomly chosen patients from the pool of the 10,000 synthetic patients. The trained ODM models were then validated on the original dataset.

Since there is no ground truth of what an optimal radiation dosage for a certain outcome would be, our evaluations are based on two metrics. The first metric is root mean square difference (RMSD) value between the ODM recommendation and the retrospective clinical decision used in treatment planning. However, since RMSD is a symmetric metric, i.e., it cannot differentiate a higher dose from a lower dose recommendation compared to the clinical decision, we separated the positive and negative clinical outcomes for additional insight. For the positive clinical outcome, a lower RMSD indicates agreement with the good clinical decisions. For the bad clinical outcome, additional comparison is needed. For this purpose, we have adopted a second metric for self-evaluation as presented in Table 1. The self-evaluation scheme is also based on the assumption that increasing radiation results in a higher value for both TCP and NTCP. Using this assumption, we can further evaluate the recommendations for patients with negative clinical outcome.

**Table 1.** Self-Evaluation Scheme. Evaluation scheme for AI recommendation is based on the positive relation between radiation dose and treatment outcomes, i.e., both TCP and NTCP increases with an increase in radiation dose. Here TC and NTC are clinical treatment outcome. TC= 1 and NTC = 0 are the only clinically positive outcome. For a patient with known treatment outcome, we can evaluate an AI recommendation by comparing it with the retrospective clinical decision. For instance, for a patient with TC = 0 and NTC = 0, a higher dose recommendation is good, while for a patient with TC = 1 and NTC = 1, a lower dose recommendation is good, and for a patient with TC=0, NTC=1, a lower dose recommendation is good. For the clinically positive cases, we cannot judge for sure if a recommendation is good unless it is within a window of the clinical dose decision. We have set the window to be 10% of the maximum dose used in the modeling.

| Self-Evaluation Scheme |  |  |  |  |  |  |  |
| --- | --- | --- | --- | --- | --- | --- | --- |
| Positive Clinical Outcome |  |  |  | Negative Clinical Outcome |  |  |  |
| TC | NTC | Relation | Remark | TC | NTC | Relation | Remark |
| 1 | 0 | $\vert AI - Cl\vert \leq 10\% \text{ of } D_{max}$ | Good | 0 | 0 | $AI \leq Cl$ | Bad |
| 1 | 0 | $\vert AI - Cl\vert > 10\% \text{ of } D_{max}$ | Not Sure | 0 | 0 | $AI > Cl$ | Good |
| 0: no event, 1: event<br>**Abbreviations:** AI, ARClIDS recommendation; Cl, Clinical decision; TC, Tumor control; NTC, radiation-induced normal tissue complication; $D_{max}$: Maximum Dose Value used in modeling | | | | 0 | 1 | $AI < Cl$ | Good |
| | | | | 0 | 1 | $AI \geq Cl$ | Bad |
| | | | | 1 | 1 | $AI < Cl$ | Good |
| | | | | 1 | 1 | $AI \geq Cl$ | Bad |

Additionally, we present a comparison for two different ARCliDS models. The first is built with Single GNN as RTOE and fully connected double deep Q-network as ODM (Single GNN RTOE+ DDQN ODM) and the second with GloGD GNN as RTOE and fully connected double deep Q-network as ODM (GLoGD GNN RTOE + DDQN ODM).

## Results

The main results are summarized and presented in Figures 10 and 11 and in SM sections S7.5 and S8.5. For the NSCLC patients, the overall RMSDs between the two ARCliDS models’ average recommendation and reported clinical decisions, ordered as GLoGD GNN RTOE + DDQN ODM and Single GNN ROTE +DDQN ODM, were 0.61 ± 0.03 Gray/fraction [Gy/frac] (mean±sem) vs 0.97 ± 0.12 Gy/frac, respectively. The overall Self-Evaluation results were Good: 55% vs 39%, Bad: 13% vs 21%, and Not Sure: 13% vs 40%, respectively. The RMSDs for patients with positive clinical outcomes were 0.66 ± 0.02 Gy/frac vs 0.96 ± 0.11 Gy/frac respectively, and the Self-Evaluation results were Good: 36% vs 18%, and Not Sure: 82% vs 64%, respectively. The RMSDs for patients with negative clinical outcomes were 0.55 ± 0.05 Gy/frac vs 0.97 ± 0.12 Gy/frac, respectively, and the Self-Evaluation results were Good: 74% vs 59%, and Bad: 26% vs 41%, respectively.

**Figure 10.**
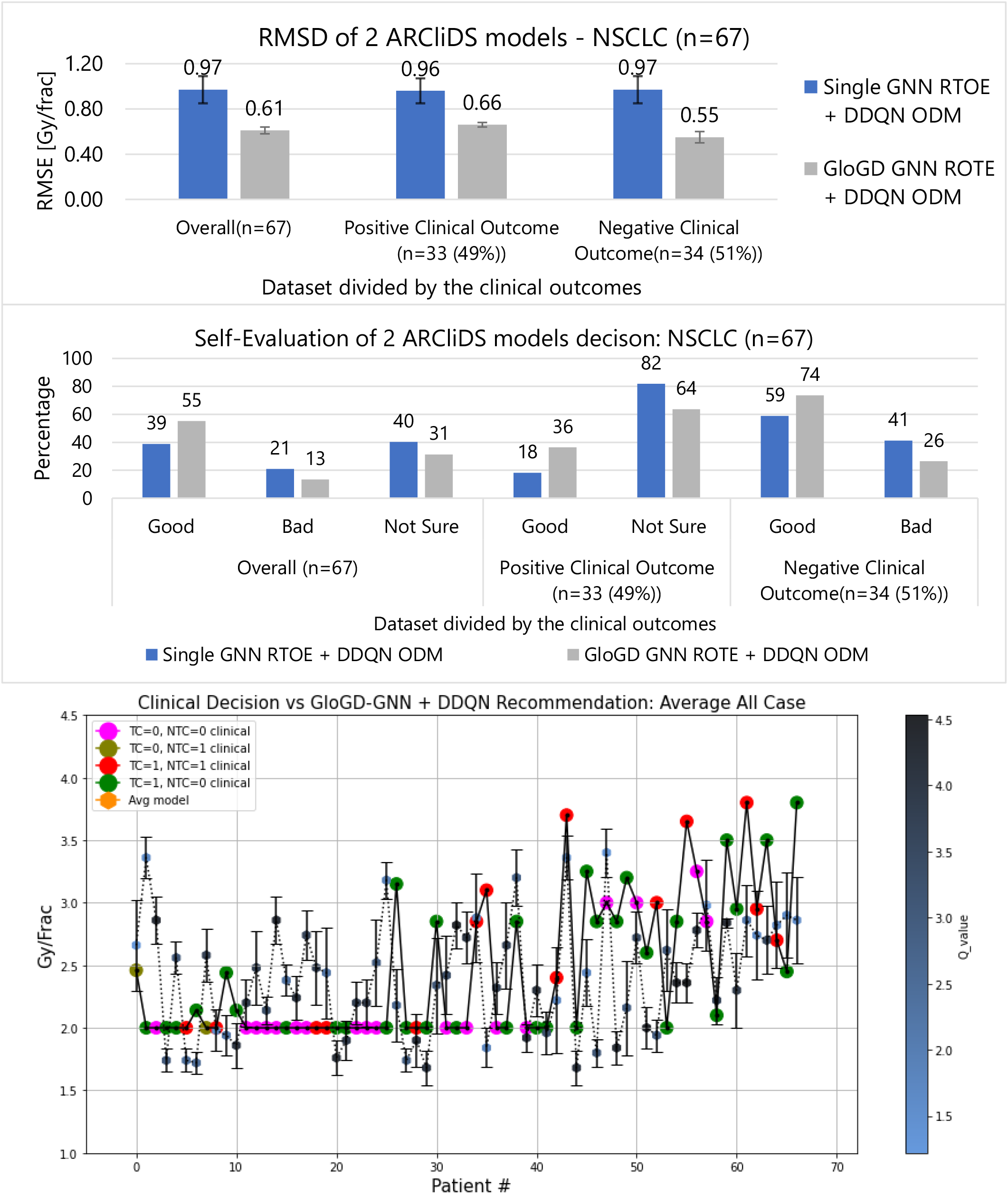
Comparison and analysis of 2 ARCliDS models trained and validated on Adaptive RT NSCLC patients. The top bar diagram presents RMSD, the middle bar diagram presents Self-Evaluation, and the bottom plot presents a visual comparison between the ARCliDS recommendation and clinical decision. The clinical decisions are color coded with the outcomes and the ARCliDS recommendations are color coded with the respective q-value. Qualitatively, the q-value can be considered as the AI confidence in its recommendations.

**Figure 11.**
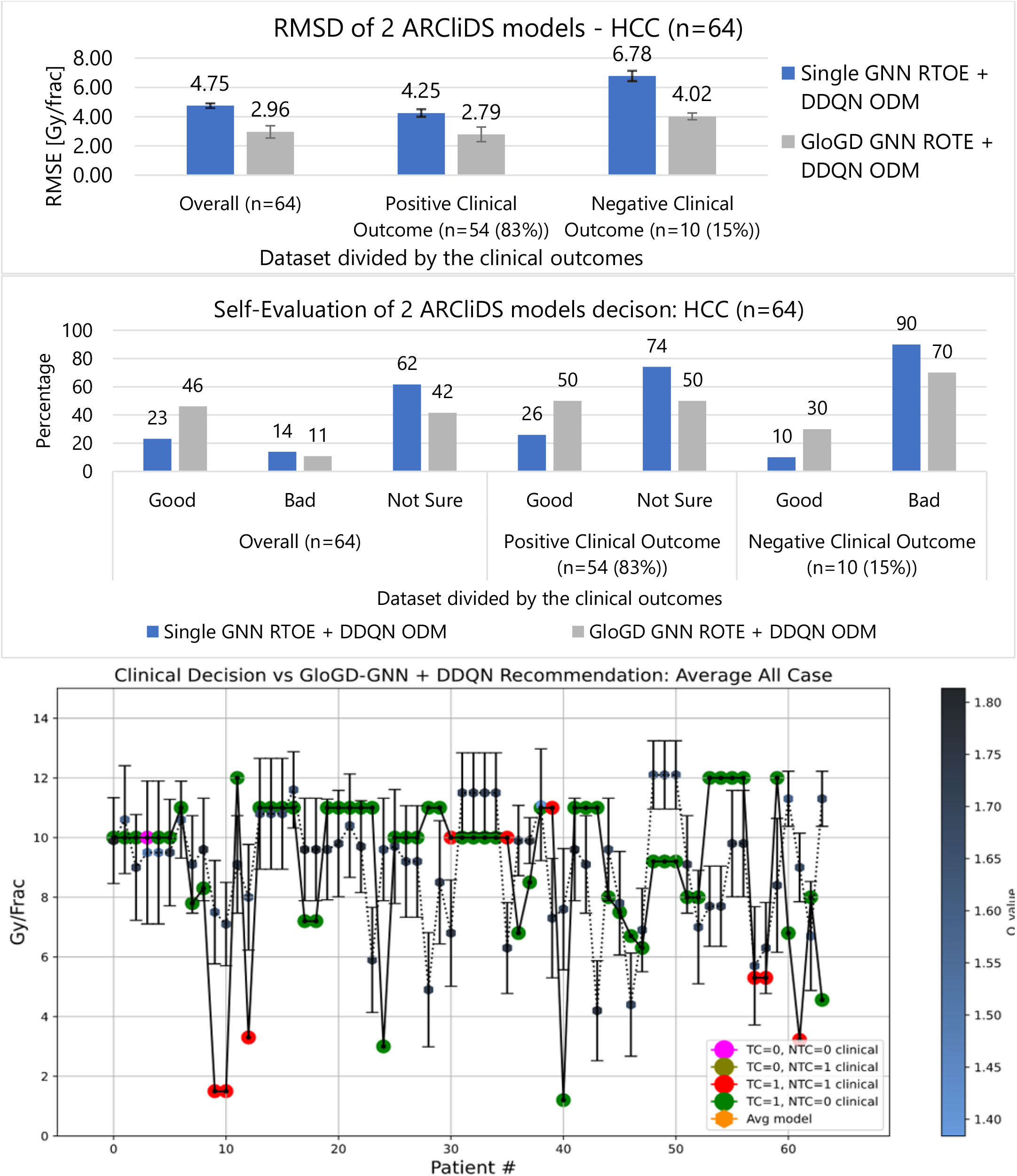
Comparison and analysis of 2 ARCliDS models trained and validated on Adaptive RT NSCLC patients. The top bar diagram presents RMSD, the middle bar diagram presents Self-Evaluation, and the bottom plot presents a visual comparison between the ARCliDS recommendation and clinical decision. The clinical decisions are color coded with the outcomes and the ARCliDS recommendations are color coded with the respective q-value. Qualitatively, the q-value can be considered as the AI confidence in its recommendations.

For the HCC patients, the overall RMSDs were 2.96 ± 0.42 Gy/frac vs 4.75 ± 0.16 Gy/Frac, respectively. The overall Self-Evaluation results were Good: 46% vs 23%, Bad: 11% vs 14%, and Not Sure: 42% vs 62%, respectively. The RMSD for patients with positive clinical outcomes were 2.79 ± 0.50 Gy/frac vs 4.25 ± 0.26 Gy/frac, respectively, and the Self-Evaluation results were Good: 50% vs 26%, and Not Sure: 50% vs 74%, respectively. The RMSD for patients with negative clinical outcomes were 4.02 ± 0.23 Gy/frac vs 6.78 ± 0.35 Gy/frac, respectively, and the Self-Evaluation results were Good: 30% vs 10%, and Bad: 70% vs 90%, respectively.

## Discussion

To our knowledge, there are software for ART^27,28^ but ARCliDS is the first interactive software dedicated to KBR-ART that will be available through a web portal. In this work, we have shown its applicability to adaptive RT and SBRT. However, ARCliD’s underlying technology can be generalized to any other DTR to optimize sequential decision-making with multi-omics data for deciding the order of treatments, including multi-modality treatment, given that an artificial treatment environment can be sufficiently modeled.

We have implemented tools such as GAN and GNN and invented novel techniques such as GloGD-GNN to overcome data-related issues for developing ARCliDS. We applied GAN to learn the underlying patient’s feature distribution and generated 10,000 synthetic patients for training the ODM. We adopted GNN for modeling RTOE as exploiting the inter-relationship between the features can improve model prediction. Mathematically, the inter-relationship can be represented by a directed graph *G*(*V, E*) where the nodes *V* represent patient features and edges *E* represent the relationships. Analyzing the inter-feature relationships before feeding it to the NN reduces the number of connections and hence simplify the learning process. As a novel approach, we applied GNN on the feature space as opposed to the sample space. As shown in the SM, every patient is represented by a directed graph of features, set by the treatment and disease type. RTOE is then designed as a graph classification problem where the node value differs from patient to patient.

As seen from Figures 10 and 11, the models in descending order according to the RMSD and Self-Evaluation measures, for both NSCLC and HCC, are GLoGD GNN RTOE + DDQN ODM, and Single GNN RTOE +DDQN ODM. As expected, correction of RTOE with GLoGD architecture helps to maintain the monotonic relationship between the outcome probability and daily dose fractionation. An example is provided in Figures S7, S8, S20, and S21 and a detailed AUROCC analysis is presented in the SM.

Our framework has some limitations. Clinically, RT dose adaptation can be performed in different ways: (1) change dose per fractions, and (2) change the number of fractions. For SBRT, the former is suitable, however for some diseases and modalities the latter may be more appropriate. For instance, when RT is combined with chemotherapy, increasing the number of fractions is preferred. Our framework only covers the former. ARCliDS uses several biomarkers such as cytokines as predictors. Due to the lack of standardization, biomarker levels of the same blood sample measured in two labs can be quite different also known as batch effect. So, biomarker levels of external dataset must be carefully examined before applying ARCliDS. For dosimetric predictor, we have used gEUD, however, for lung and liver, mean dose could also be applied. Another limitation is the number of NTCP’s considered in ARCliDS. In practice there may be more than 1 normal tissues of interest. For NSCLC, heart and lungs are the dose-limiting organs at risk (OAR). For HCC, although liver is usually the main OAR, in some patient, who has tumors near the intestine, intestine is also considered during designing the treatment plan. Finally, beside data-related shortcomings, ARCliDS prediction and recommendation uncertainty, which is based on statistical ensembles, can be improved by training more models; however, this will require more computational power and time.

Although we have the largest dataset of its kind, a larger sample size and balanced dataset will improve ARCliDS performance. We dedicate subsequent paragraphs to discuss data-related limitations, methods we implemented to overcome those limitations, and other possible solutions.

The learning of an environment model is the bottle neck of ARCliDS. For learning a good ARTE, a sufficient sample size and a balanced dataset are necessary. In the adaptive HCC patient’s cohort, only 1 patient did not achieve local control. As a result, RTOE for TCP had an unusually high AUROCC uncertainty. Although we applied class-imbalance correction techniques such as SMOTE and weighted loss function, we witnessed that such techniques fall short in correcting a highly imbalanced dataset. In addition, the toxicity count was also low --there were only 7 patients that showed toxicity. While this is a clinically desirable result, it hinders the learning process and hampers model generalizability. To make the matter worse, patients with highest liver gEUD didn’t show toxicity as shown in Figure S19. This reflects inter-patient heterogeneity, where some patients have poor pre-treatment liver function, who at a higher risk of toxicity for lower dose. Nevertheless, we performed a hyperparameter search for maximizing the generalizability of ARTE.

High noise-to-signal ratio due to inter-patient heterogeneity becomes even more pronounced with a small sample size. The medical field is especially doomed with a small sample size primarily due to the privacy issue^29^. Such issues make it difficult to learn correct trends in purely data-driven learning. We found that Single-GNN RTOE predicted unphysical trends between daily dose fractionation and TCP/NTCP. For correction, we applied a GLoGD-GNN architecture to infuse prior knowledge into the data-driven technique. We found that it corrects the trend and can also increase the model predictability. Alternatively, distributive learning features such as federated learning can be added to ARCliDS to overcome the small sample size issue. In federated learning approach only the model parameters are shared and data stays within the firewall of individual institutions^30^.

Sample size issue in training ODM can also be overcome by using synthetic patients. Since ODM of ARCliDS learns via model-based reinforcement learning, computationally the task of ODM is to learn ARTE. This can be considered as an interpolation problem in a continuous feature space. This problem can be tackled using brute-force by exhaustively selecting patient’s state. However, this assumes a uniform distribution which is generally not true. Therefore, we applied generative adversarial network (GAN) to learn the underlying patient’s feature distribution, as shown in SM and generated synthetic patient states for training the ODM. In principle, a conditional GAN^31^ can be applied to generate patients states distribution along with the outcome, however, a low sample size coupled with severe class-imbalance makes it impossible to correctly learn the underlying conditional probability distribution.

We found that the RMSD values for adaptive SBRT in HCC was higher than adaptive RT in NSCLC. There are three reasons for the higher RMSD value: (1) A larger range of adaptive dose values was explored for SBRT, i.e., 1 to 15 Gy/frac compared to 1.5 to 4 Gy/frac; given that the sample sizes are comparable, the datapoints for HCC are much sparser resulting in higher interpolation error; (2) most of the patients with a clinically negative outcome for SBRT received a lower adaptive dose than the positive case; this can confuse the RTOE, which assumes higher doses results in higher TCP and NTCP; (3) due to class-imbalance, the corrected GLoGD-GNN RTOE yielded a flatter monotonic relation than expected that did not spanned the whole probability space; we observed that the AI agent failed to satisfy the population-based goal of TCP > 90% and NTCP < 25%. So, we could set the computation goal of TCP >50% and NTCP < 50%. A smaller RMSD value can be achieved with a large sample size and well-balanced dataset.

In conclusion, we have built a user-friendly software for AI-assisted clinical decision-making and demonstrated its performance in adaptive RT. The underlying technology behind the software is generalizable to other sequential decision-making tasks in oncology. We employed GNNs to exploit the inter-feature relationship. We trained and validated our software in two different treatment types for two different diseases. We repeated the training and validation for 2 different models to test our hypothesis of improved model performance. The results confirmed our hypothesis. Statistical Ensemble was adopted to assess the model uncertainty.

## Data Availability

The training data analyzed in this study are obtained from the University of Michigan. Restrictions apply to the availability of these data, which were used under the data sharing protocol for this study. ARCliDS is publicly available at https://arclids.shinyapps.io/ARCliDS for a limited time.

## Contributors

I.E.N. conceived the study. I.E.N., R.K.T.H., M.M.M., J.J., and I.D.D, supervised the project. T.S.L., S.J., M.M.M., R.K.T.H., and K.C supervised and took part in the original clinical trials. D.N. designed and developed the software, wrote the programming codes, and carried out model training and validation. K.C. collected the data, provided clinical insight, and evaluated the software. Y.L. conducted feature selection. D.N., W.S., and J.J. contributed to the algorithmic development of the underlying methodologies. All authors carefully analyzed the methods and results. D.N. drafted the manuscript and all authors critically read and contributed to the final version.

## Supplementary Materials

### S1. Feature Selection and Graph Building via Markov Blanket Approach

In this work, we adopted the feature graph from Yi et al.’s work on multi-objective Bayesian networks for radiotherapy outcome prediction^1^. The steps of building an appropriate multi-objective Bayesian network (MO-BN) for joint prediction of TCP and NTCP include large-scale feature selection and network structure learning. The first step intends to identify important features from a high-dimensional dataset by exploring extended Markov blankets (MBs) of TCP and NTCP. An MB is an inner family found by constraint-based algorithms such as incremental association Markov blanket (IAMB) and Hiton approaches. The MB contains all variables carrying information about TCP and NTCP that cannot be obtained from any other variables. For each member in the MB of TCP and NTCP, a next-of-kin MB for this member can also be derived, which is combinedly known as extended MB.

The second step is to combine the important features from the extended MBs and search for the best stable MO-BN for joint prediction. After accommodating radiobiologically plausible relationships based on reported literature, Tabu Search is employed to generate a stable MO-BN structure from 300 randomly generated bootstrap samples. Bayesian Dirichlet equivalent (BDe) that provides an inherent penalty for model complexity is used as a scoring function to obtain the stable MO-BN. However, an initial stable network may not be the best model for joint TCP and NTCP prediction due to unimportant or redundant features in it. So, a leaf node is found out by increasing the arc threshold in the MO-BN generation. After removing the leaf node, a stable MO-BN can be generated again from Tabu Search. The process continues until the resulting MO-BN reaches its maximal prediction performance based on cross-validation. The network structure found for NSCLC and HCC are presented in Figures S5 and S18.

### S2. ARCliDS Training Sequence

1. Train Artificial Radiotherapy Environment (ARTE) via supervised learning using patient’s feature and dose plan as the input, and corresponding clinical outcomes as the label.
  a. Model Transition Function (TF) to take in mid-treatment feature (*s*_*eval*_) and predict after-treatment feature (*s*_*adapt*_) for the adaptation daily dose fractionation (*d*_*adapt*_)

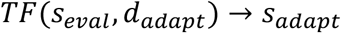

*More details are presented in Section S4*.
  b. Train RT Outcome Estimators (RTOE) to estimate treatment outcome (*tcp, ntcp*) for the predicted *s*_*adapt*_ and covariate *c* as a binary classifier as shown in Figure S1.

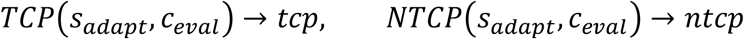

*More details are presented in Section S5*.
  c. Design Reward Function that yields reward *r* which increases with increasing *tcpi*and decreases with increasing *ntcp*. For instance:

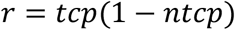
2. Train Optimal Decision Maker (ODM) via deep reinforcement learning on a trained ARTE from step 1.
  a. Inputs *s*_*eval*_, *c, and d*_*adapt*_ to the ARTE and store *s*_*adapt*_, *tcp, ntcp* and *r* for all GAN-generated synthetic patients

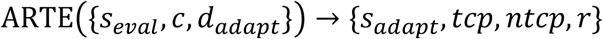

*More details on GAN are presented in Section S6*.
  b. Train ODM via Deep Q-learning as a one-step optimization process, i.e., one training episode for all patients representing the adaptive phase.

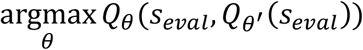

*More details are presented in Section S7*.

**Figure S1:**
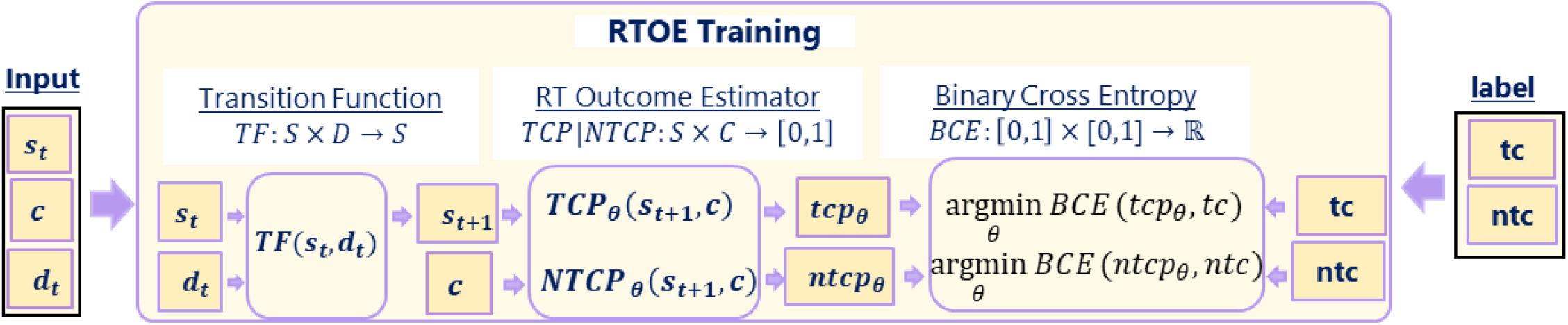
Training RTOE as a binary classification. Here *θ* represent tunable weights, which is learned by minimizing the binary cross entropy loss function. In the case of graph neural networks, the features are input as a directed graph *G*(*V, E*), where the nodes, *V*, represents the features, *X* ⊂ ℝ^*k*×1^ and edges, *E*, represents the inter-feature connections. Edges, *E*, are mathematically represented by adjacency matrix, *A* ⊂ ℝ^*k*×*k*^. During feedforward, the signal propagation is multiplied by the adjacency matrix given by *H*^*i*^ = σ(*AH*^*i*−1^*θ*^*i*^) for zero bias, where *σ* is the activation function, *θ*_*i*_, is the weight of the ith hidden layer, and *H*_*i*−1_ is the matrix containing i-1st layer embeddings. Notice that multiplying by *A* preserves the only important inter-feature connections and eliminates computational redundancies; each node embedding is computed only once in contrast to fully connected neural network. For our case, features *X* is a concatenation of dosimetric variable, *s*, and other multi-omics covariate, *c*. Note, each patient is represented by a graph in the feature space and the binary classification is performed in the sample space as a graph classification problem^2^.

### S3. Transition Function for generalized equivalent uniform dose (gEUD)

GEUD absorbed by a patient must increase monotonically with increasing daily dose fractionation based on known principles of radiation biology^3^. To enforce this monotonic relation, we use a function of the form,

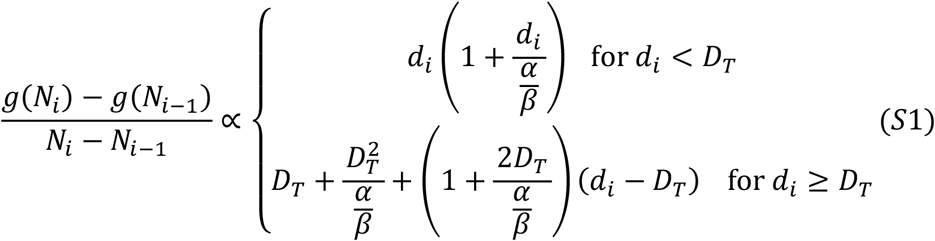

where, *i* is the *i*^th^ daily dose fractionation, *g* stands for gEUD, N stands for number of daily dose fractions, *d* stands for amount of dose fractions, *D*_*T*_ stands for threshold dose related to the linear-quadratic-linear (LQL) model, and *α*/*β* ratio is a parameter that differentiates tissue type.

**Figure S2:**
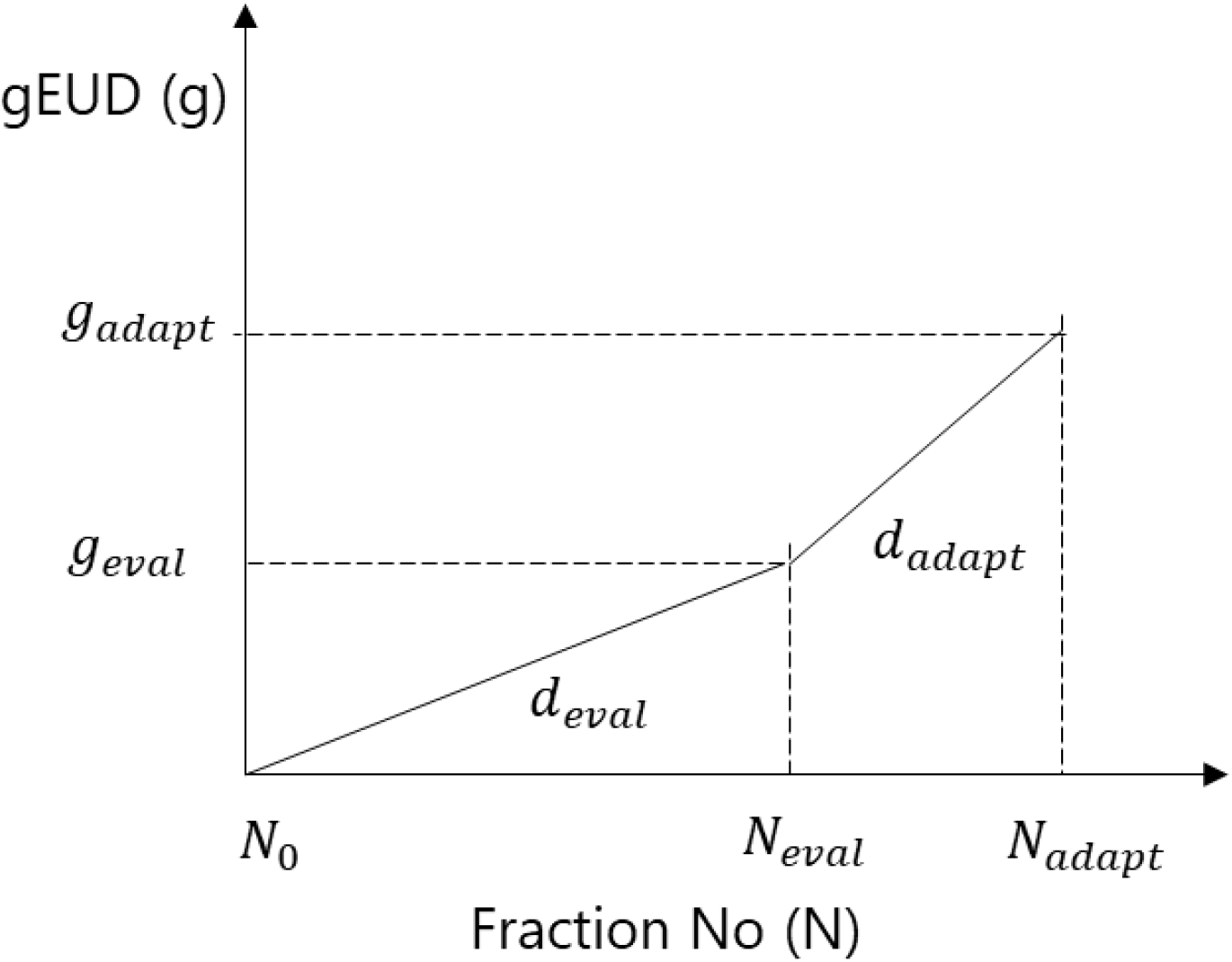
Transition Function for gEUD. Eval stands for the Evaluation Phase and adapt stands for the Adaptation Phase. Repeated from Figure 4 from the main text for clarity.

As shown in Figure S2, for our case, two relations arise from relation (S1), as follows:

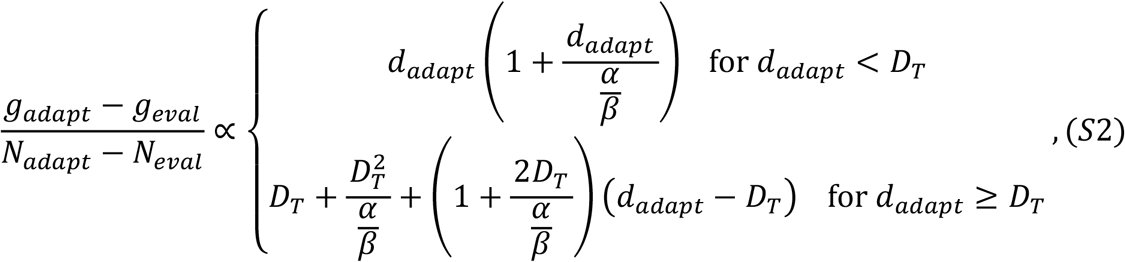

And,

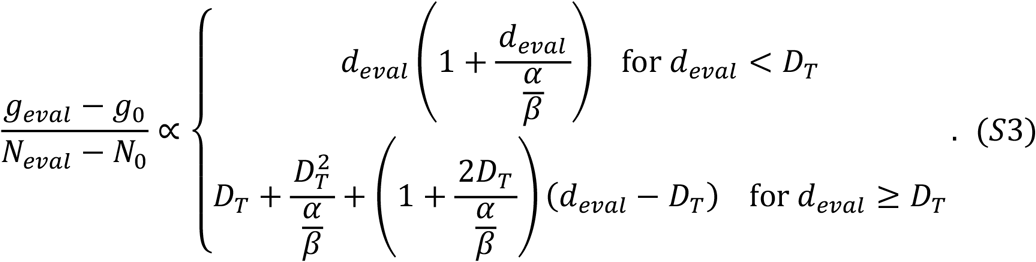

where, *g* stands for gEUD, *N* for *n*th daily dose fractions, *d* for dose fractions, *D*_*T*_ for threshold doses, and *α*/*β* ratio is a tissue-specific parameter. The subscript 0, eval, and adapt of *N* and *g*corresponds to pre-, mid-, and after-treatment, respectively while *d*_*eval*_ and *d*_*adapt*_ corresponds to applied daily dose fractionations during the evaluation phase and adaptive phase, respectively.

Furthermore, four scenarios can arise. The four different gEUD transition functions can be derived by taking the ratios of relations (S2) and (S3) as presented in Table S1.

**Table S1:** Transition Function for final gEUD (i.e., *g*_*adapt*_)

| $g_{adapt} =$ | $d_{adapt} < D_T$ | $d_{adapt} \geq D_T$ |
| --- | --- | --- |
| $d_{eval} < D_T$ | $g_{eval} \left( 1 + \left( \frac{n_{adapt}}{n_{eval}} \right) \left( \frac{d_{adapt}}{d_{eval}} \right) \left( \frac{d_{adapt} + \frac{\alpha}{\beta}}{d_{eval} + \frac{\alpha}{\beta}} \right) \right)$ | $g_{eval} \left( 1 + \left( \frac{n_{adapt}}{n_{eval}} \right) \left( \frac{\gamma d_{adapt} - D_T^2}{d_{eval} \left( d_{eval} + \frac{\alpha}{\beta} \right)} \right) \right)$ |
| $d_{eval} \geq D_T$ | $g_{eval} \left( 1 + \left( \frac{n_{adapt}}{n_{eval}} \right) \left( \frac{d_{adapt} \left( d_{adapt} + \frac{\alpha}{\beta} \right)}{\gamma d_{eval} - D_T^2} \right) \right)$ | $g_{eval} \left( 1 + \left( \frac{n_{adapt}}{n_{eval}} \right) \left( \frac{\gamma d_{adapt} - D_T^2}{\gamma d_{eval} - D_T^2} \right) \right)$ |
| $n_{adapt} \equiv N_{adapt} - N_{eval}, \quad n_{eval} \equiv N_{eval} - N_0, \quad \gamma \equiv \frac{\alpha}{\beta} + 2D_T, \quad g_0 = 0, \quad N_0 = 0$ | | |

Note: For traditional RT, only the first scenario is relevant. However, for SBRT, all four scenarios come into play.

### S4. RT Outcome Estimator (RTOE)

#### S4.1 Generalized Logistic Function Guided Double GNN (GLoGD-GNN) for Monotonic TCP/NTCP

Likewise, both TCP and NTCP must increase monotonically with increasing radiation dose. Due to the patient heterogeneity related noise in the data, RTOE composed of a single GNN classifier could not adequately represent the monotonic relationship. Thus, we developed a guided dual GNN architecture named GLoGD-GNN, which comprises of two GNNs, *μ*_*GNN*_ and *T*_*GNN*_ that is fed into a generalized logistic function along with the *g*_*adapt*_. GloGD-GNN can be summarized as follows,

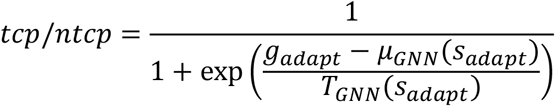

where, *μ*_*GNN*_ and *T*_*GNN*_ are GNN’s with sigmoid output layer and takes in *s*_*adapt*_ as input. GEUD *g*_*adapt*_ is min-max normalized before feeding it to the logistic function. During training, the weights of the GNN’s are updated alternately; when one is training, the other is kept frozen.

#### S4.2 RTOE Hyper Parameter (HP) Tuning and Validation

We performed a grid search for HP tuning. We used Adam optimizer for the optimization and Area Under the Receiver Operating Characteristic Curve (AUROCC) as the performance metric. We applied 10-fold stratified shuffle 80-20 split on the imbalanced dataset, where the data sets were stratified according to the binary outcome so that each test sets contained the same ratios of the outcome class. In addition, we randomly oversampled the minority class of each training split. Both training and validation dataset were batched and randomly sampled during model training.

For the Single GNN architecture, we searched the following HP space:

Optimizer learning rate = [0.0001, 0.0005, 0.001, 0.005],

Number of nodes = [256, 128, 64], Training Epoch = [50, 100, 200, 300]

For the GloGD GNN architecture, the HP search space was as follows:

Optimizer learning rate for mu GNN = [0.0001, 0.0005, 0.001],

Optimizer learning rate for T GNN = [0.0001, 0.0005, 0.001],

Number of nodes = [256, 128, 64], Training Epoch = [100, 200, 300, 400]

After selecting the best HP that corresponded to the best average test AUROCC score, we reran validation using the best HP to test the reproducibility. The best HP and all of the original datasets were then used to train five RTOE models. The results from validation on the NSCLC and HCC datasets are summarized in Figure S3.

**Figure S3:**
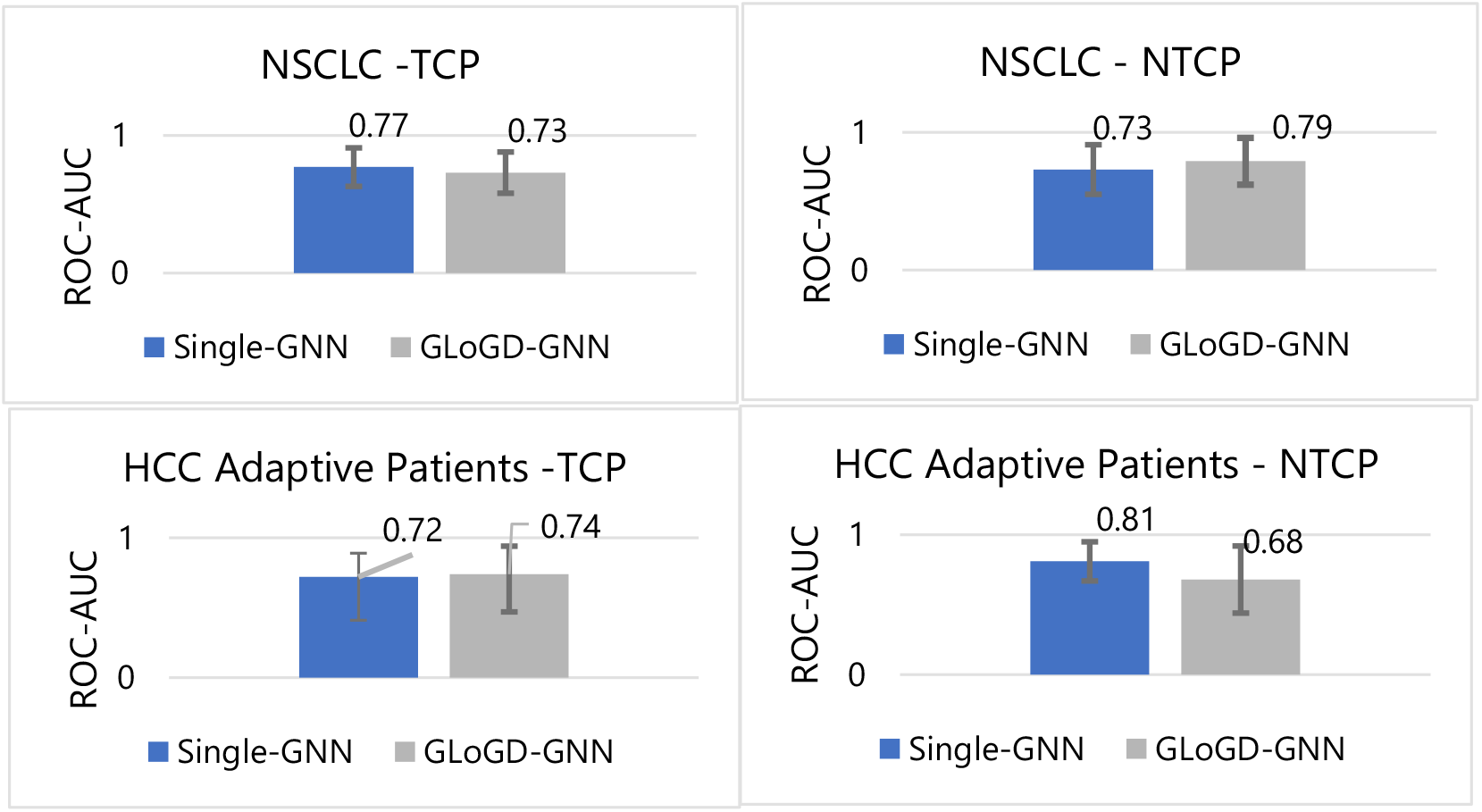
RTOE model performance for NSCLC and HCC data in Area Under the Receiver Operating Characteristic Curve (AUROCC). The mean AUROCC value is the area under the mean true positive rate curve, while the standard deviation is calculated from the AUROCC of 10 individual model output.

### S5. Synthetic Patients via Generative Adversarial Network (GAN)

To extend the sample size of our dataset, we generated 10,000 synthetic patients via Wasserstein Generative Adversarial Network with Gradient Penalty (WGAN-GP)^4,5^. We compared the distribution of synthetic patient with the original patient population data using the Jensen Shannon Divergence (JSD) metric as shown in the subsequent sections. JSD value of 0 means complete overlap and 1 means complete separation. We did not perform any statistical hypothesis test on the learned distribution as it is not necessary for the training of ODM. In principle, a uniform distribution works just as fine, however, with increase in computational complexity. Nevertheless, we ascertained the similarity by visual inspection and JSD. A comparison between the original and generated datasets are presented in Figures S13 and S26.

We applied a four-layer deep neural network with 256 nodes for both the discriminator and generator. We designed the generator to take in a 64-dimension normally distributed random numbers. We applied ADAM optimizer for the training with the learning rate of 1E-4, *β*1 of 0.5, and *β*2 of 0.9 and trained the GAN for 500 epochs. To maintain the stability by keeping discriminator ahead, we trained the discriminator 5 times for generator’s every training epoch. As in the original work, we used a Gradient Penalty weight of 10 for our training.

### S6. Double Deep Q-Learning for Optimal Decision Maker (ODM)

ODM was trained using Double deep Q-learning (DDQN) algorithm. DDQN uses two neural networks, namely policy Q-net and target Q-net. This approach helps in correcting the overestimation of the q-values. We used a 5 layer deep 256 node wide architecture. For training parameters, batch size was set to 128, gamma (discount factor) to 0.8, Polyak factor to 0.99 (used for updating the target net’s parameter) and Adam optimizer’s learning rate to 0.0005.

For robustness, a planning and learning scheme were applied. Since in clinical setting, the physicians only get to select one action, we treated the problem as a one-step optimization problem, i.e., all episodes were terminated after one step. So, we performed an exhaustive search (i.e., explored all actions), before training the q-network. Note that this results in a greedy algorithm.

In the planning phase, 4000 out of 10,000 synthetic patients were randomly selected and their next states for all possible actions were exhaustively found using ARTE and stored. Additionally, the reward values and binary information on where they met the goal outcome were also stored. Then the DDQN was trained for 300 epochs. Huber loss was used as the loss function for updating the policy Q-net and Polyak update us used for updating the target Q-net.

The update rule for double deep Q-net learning is as follows,

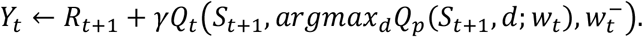

Here, *Y*_*t*_ is the target value, *γ* is the discount factor, *w*_*t*_ is the weights of policy net *Q*_*p*_, and 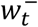 is the weights of the target network *Q*_*t*_. The policy net is updated with the target value every step via stochastic gradient descent. However, the target net is kept almost fixed, only updating in small increment by copying parameters from target net via Polyak averaging, i.e.

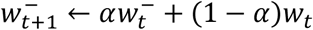

where *α* is the polyak factor.

For quantifying model uncertainty (or decision confidence), we trained 5 identical models.

**Figure S4:**
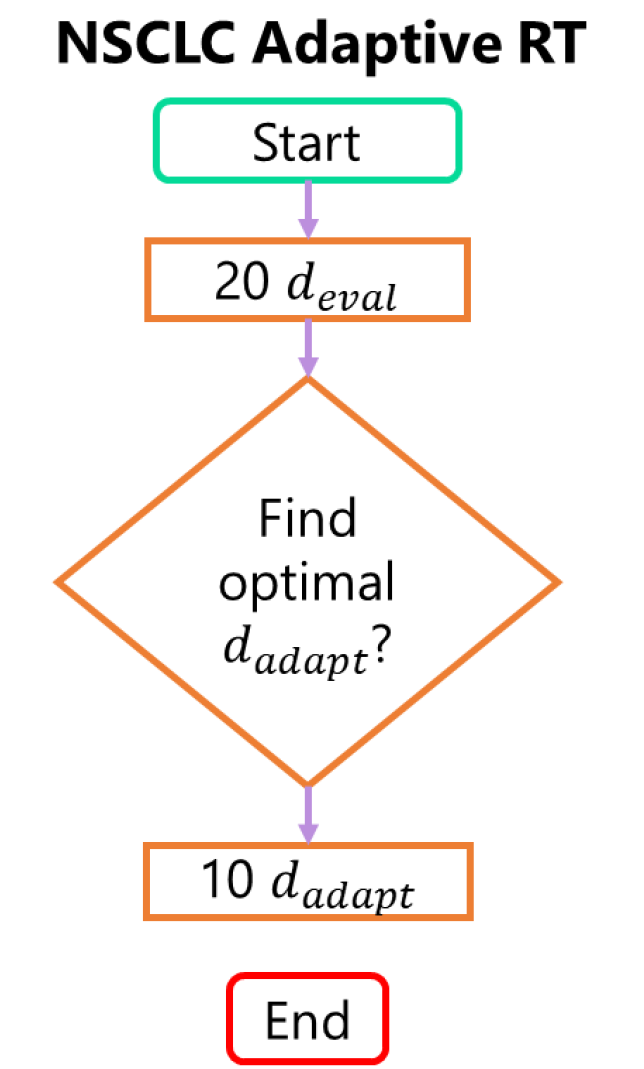
Clinical decision-making in NSCLC KBR-ART. In the evaluation phase, patients received 20 daily doses of preplanned fractions. After that the patients are evaluated, and an adaptive dose plan is designed and administered in 10 daily doses.

### Use Case 1: NSCLC

#### S7.1 Data Description

Dense multi-omics information on 117 patients were available. Out of that, 67 patients had complete information.

**Table S2:** NSCLC Patient Characteristics

| Variable | Category | Patient Count (n =67) |
| --- | --- | --- |
| Sex | Male | 51 |
|  | Female | 16 |
| Age | Median (Q1-Q3) | 66 (59-73) |
|  | Range | 55 – 85 |
| Stage | I | 4 |
|  | II | 6 |
|  | III | 57 |
| Smoking | Yes | 65 |
|  | No | 6 |
| COPD | Yes | 28 |
|  | No | 39 |
| CVD | Yes | 22 |
|  | No | 45 |
| Hypertension | Yes | 41 |
|  | No | 26 |
| Histology | Adenocarcinoma | 15 |
|  | Squamous Cell Carcinoma | 27 |
|  | Large Cell | 0 |
|  | Poorly Differentiated | 25 |
| Chemo | Yes | 57 |
|  | No | 10 |
| Outcome | LC = 0 | 20 |
|  | LC = 1 | 47 |
|  | RP2 = 0 | 51 |
|  | RP1 = 0 | 16 |
| COPD: Chronic obstructive pulmonary disease<br>CVD: Cardiovascular Diseases<br>LC: local control<br>RP2: Radiation induced pneumonitis of grade 2 or higher |  |  |

**Figure S5:**
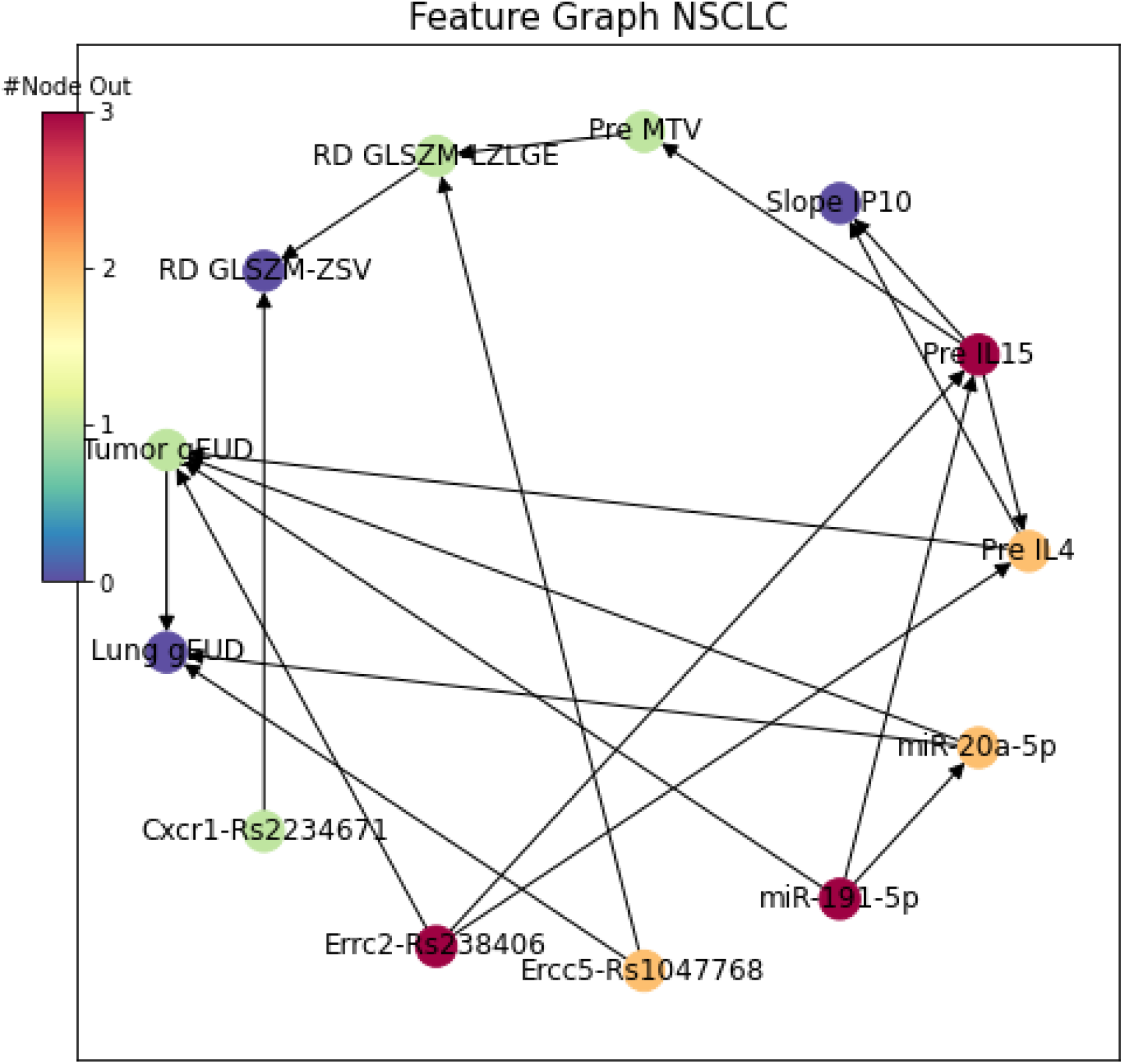
Directed graph showing the inter-relation between the NSCLC patient’s features. The nodes, which represent features, are color coded with the number of outgoing relationships. Pre stands for pre-treatment observation, RD and slope stands for relative difference and change in feature value between pretreatment and mid-treatment observation, respectively.

**Figure S6:**
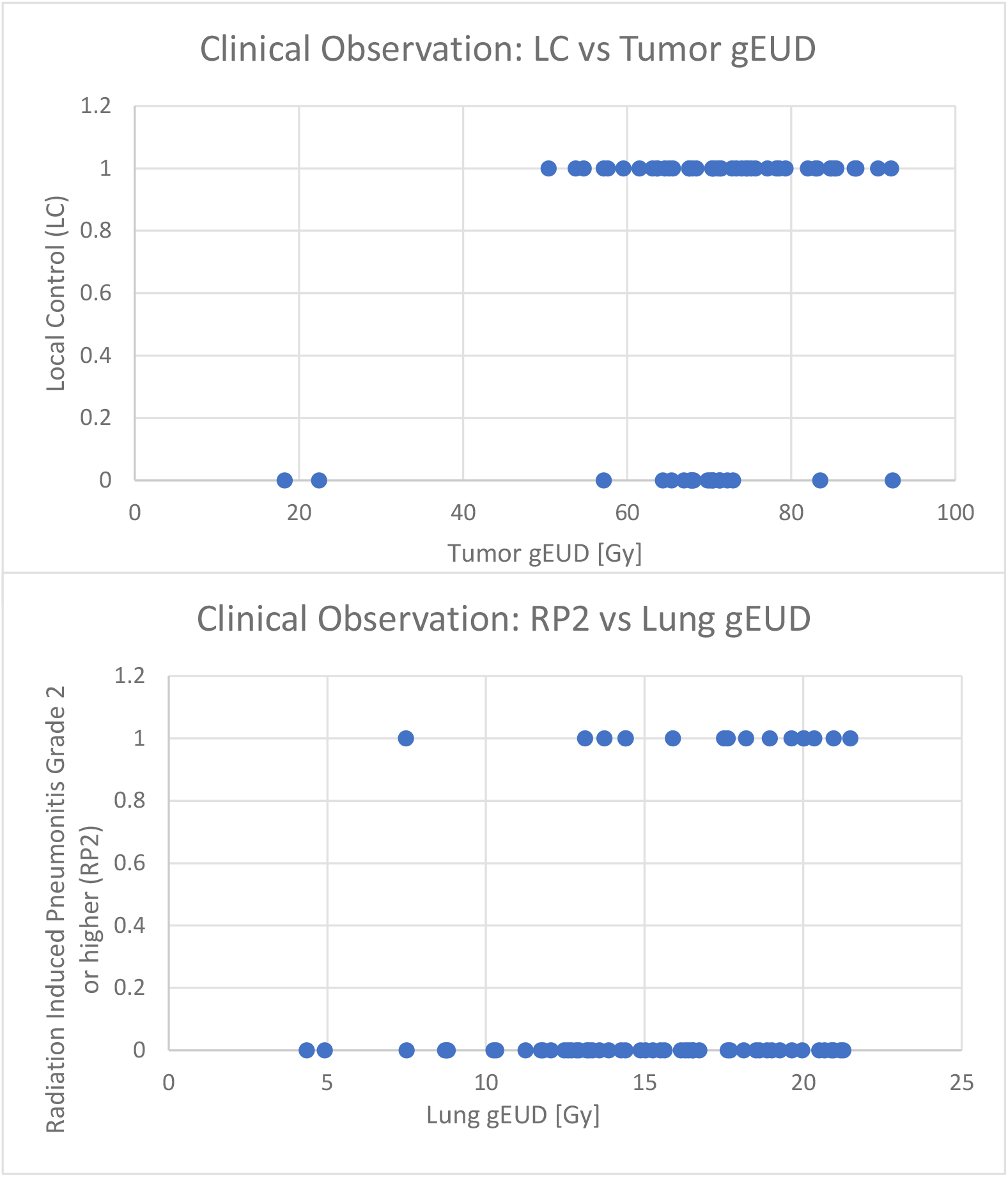
Plots showing NSCLC population tumor gEUD vs observed local control and lung gEUD vs observed radiation induced pneumonitis of grade 2 or higher. This plot captures inter-patient heterogeneity which shows patient’s diverse treatment response.

**Table S3:** NSCLC Patients’ feature description.

| Patient Variable | Biological/Clinical Characteristics |
| --- | --- |
| **Cytokines/Signaling molecule** |  |
| **IL4** | Interleukin 4 is a Th2 cytokines: (i) Regulates antibody production, hematopoiesis, and inflammation. (ii) Promotes the differentiation of naïve helper T cells into Th2 cells. (iii) Decreases the production of Th1 cells.<sup>6-8</sup> |
| **IL15** | Interleukin 15 is a Th2 cytokines: (i) Induces activation and cytotoxicity of NK (natural killer) cells. (ii) Activates macrophages. (iii) Promotes proliferation and survival of T and B- lymphocytes and NK cells.<sup>6-8</sup> |
| **IP10** | IP10 (Interferon gamma-induced protein 10) is secreted in response to IFN-γ by various cells including monocytes, endothelial and fibroblasts. (i) Acts as chemoattractant for monocytes/macrophages, T cells, NK (natural killer) cells, and dendritic cells. (ii) Promotes T cell adhesion to endothelial cells. (iii) Antitumor activity (iv) Inhibition of bone marrow colony formation (v) Angiogenesis.<sup>6-8</sup> |
| **CD4 T helper cells** are lymphocytes that strongly modulate the response of the immune system against cancer cells proliferation and tumor growth. They are classified into Th1 and Th2 cells. Th1 and Th2 cells generate Th1 and Th2 immune response by releasing cytokines. *Th1 immune response is proinflammatory and Th2 immune response is anti-inflammatory.*<br>**IFN-γ:** Th1 cytokines: (i) Enhances the microbicidal function of macrophages. (ii) Promotes the differentiation of naïve helper T cells into Th1 cells. (iii) Activates polymorphonuclear leukocytes, cytotoxic T cells, and NK cells. <sup>6</sup> |  |
| **Tumor PET Imaging features/ Radiomics** |  |
| **MTV** | Metabolic tumor volume was delineated from PET imaging using a method combining the tumor/aorta ratio auto segmentation and CT anatomy-based manual editing.<sup>9</sup> |
| **GLSZM-LZLGE** | Radiomics features: the large zone low gray-level emphasis (LZLGE) feature of a gray-level size zone matrix (GLSZM) is defined as $\sum_{i=1}^{N_g} \sum_{j=1}^{L_z} \frac{j^2 p(i,j)}{i^2}$. refer to Appendix A5 of Carrier-Valliere's thesis <sup>10</sup> for the Notations. |
| **GLSZM-ZSV** | Radiomics features: the zone-size variance (ZSV) feature of a gray-level size zone matrix (GLSZM) is defined as $\frac{1}{N_g \times L_z} \sum_{i=1}^{N_g} \sum_{j=1}^{L_z} (jp(i,j) - \mu_j)^2$, refer to Appendix A5 of Carrier-Valliere's thesis <sup>10</sup> for the Notations. |
| **Dosimetry** |  |
| **Tumor gEUD** | Generalized equivalent uniform dose (gEUD) of tumor converted from EQD2 (Equivalent Dose at standard 2 Gy per fraction) dose distribution: $gEUD = (\sum_i v_i eqd_2^a)^{\frac{1}{a}}$ and $eqd2 = N_{frac} \times d \times \frac{d+\alpha/\beta}{2+\alpha/\beta}$ where $\alpha/\beta = 10$Gy, is the radiation fractionation sensitivity of cell, a = -10 is an organ specific parameter, and $v$ is the fractional organ volume obtained from the 3D dose distribution <sup>11</sup> |
| **Lung gEUD** | Generalized equivalent uniform dose (gEUD) of lung converted from EQD2 (Equivalent Dose at standard 2 Gy per fraction) dose distribution: $gEUD = (\sum_i v_i eqd_2^a)^{\frac{1}{a}}$ and $eqd2 = N_{frac} \times d \times \frac{d+\alpha/\beta}{2+\alpha/\beta}$ where $\alpha/\beta = 4$Gy, and a =1 <sup>11</sup> |
| **Genetics** |  |
| **Cxcr1-Rs2234671** | A SNP in the gene cxcr1, also known as Interleukin 8 receptor alpha (IL8RA), related to radiation induced toxicity in non-small cell lung cancer. <sup>12</sup> |
| **Ercc2-Rs238406** | A SNP in the gene ercc2 known to repair DNA excision and related to risk of lung cancer <sup>13</sup> |
| **Ercc5-Rs1047768** | A SNP in the gene ercc5 also known to repair DNA excision and related to lung cancer susceptibility <sup>14</sup> |
| **SNP:** Single nucleotide polymorphism is a substitution of single nucleotide that occurs at a specific position in the genome via mutation. |  |
| **MicroRNA** |  |
| **miR-191-5p** | miR-191 is abnormally expressed in many cancer-types which regulate vital cellular processes such as cell proliferation, differentiation, apoptosis, and migration by targeting important transcription factors, chromatin remodelers, and cell cycle associated gene. <sup>15</sup> |
| **miR-20a-5p** | miR-20a has been found to be associated with lung cancer. It is encoded by a gene located on chromosome 13q31. It is involved in several oncogenic processes like cellular proliferation, angiogenesis, and apoptosis. <sup>16</sup> |
| MicroRNAs are small non-coding RNAs that negatively regular target gene expression through mRNA degradation or translation inhibition. <sup>17</sup> |  |

#### S7.2 GLoGD-GNN for Monotonic TCP/NTCP

**Figure S7:**
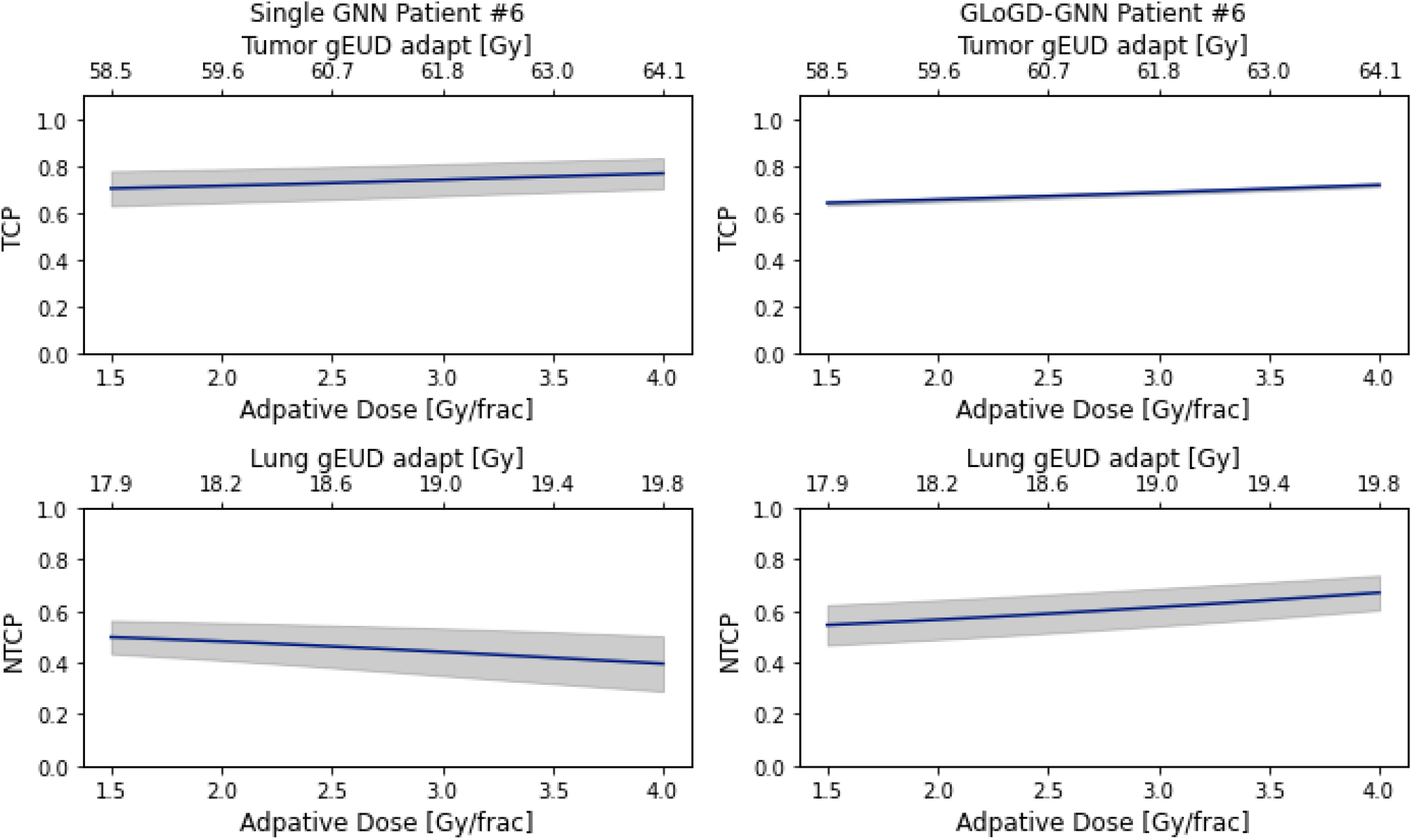
NSCLC outcomes estimate for adaptive dose ranging from 1.5 to 4.0 Gy/frac. The GLoGD-GNN correction successfully established a monotonic relationship for the NTCP. The model uncertainty is obtained from an ensemble of five RTOE model and presented as ±1 standard deviation. Note: the sample/patient IDs in the figures are arbitrary, unidentifiable, and meaningless and they were not known to anyone outside the research.

**Figure S8:**
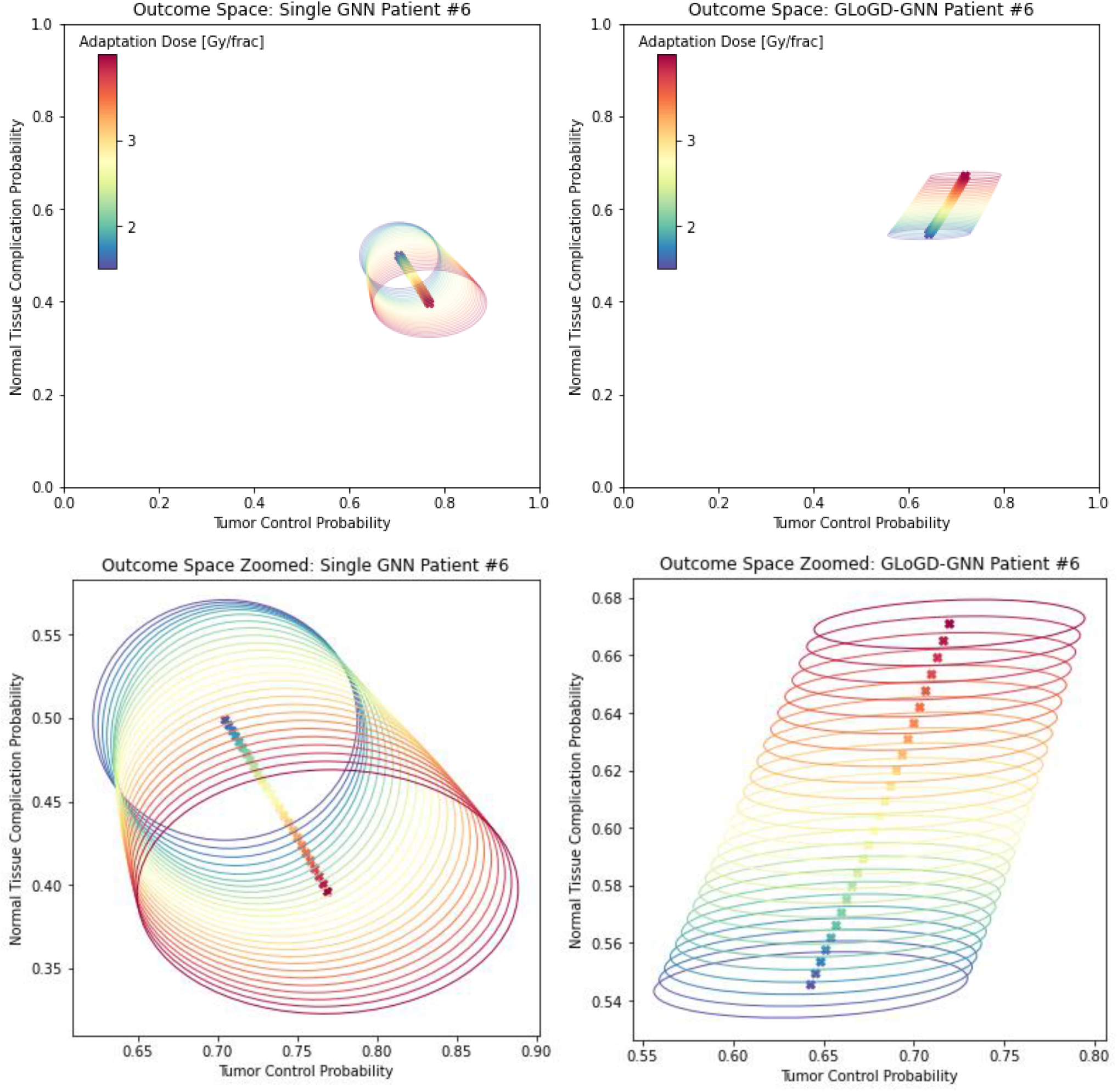
NSCLC outcomes estimate for adaptive dose ranging from 1.5 to 4.0 Gy/frac in the outcome space spanned by TCP and NTCP. Model uncertainty is obtained from an ensemble of five RTOE model and presented as an eclipse set by the Covariance matrix. In the left figure, NTCP is decreasing with increasing dose value. In the right figure, GLoGD-GNN correctly flipped the dose order i.e., NTCP is increasing with increasing dose value. Note: the sample/patient IDs in the figures are arbitrary, unidentifiable, and meaningless and they were not known to anyone outside the research.

#### S7.3 RTOE Hyper Parameter Tuning

Tables S4-S7 lists the top 10 performing HP for the NSCLC dataset followed by receivers operating characteristics (ROC) for the best performing HP (marked in bold) shown in Figures S4-S12. To check for the reproducibility, ROC were generated by retraining the models with the best performing HP. The reported AUROC values are in the mean±stdev format, where the mean AUROCC value is the area under the mean true positive rate curve, while the standard deviation is calculated from the AUROCC of 10 individual model output.

**Table S4:** Top 10 HP for **NSCLC TCP (LC)** with **Single GNN**

| HP # | Opt lr | Node | Epoch | Training AUC |  | Avg Validation AUC |  |
| --- | --- | --- | --- | --- | --- | --- | --- |
|  |  |  |  | Mean | Stdev | Mean | Stdev |
| **1** | **0.0001** | **128** | **50** | **0.85** | **0.03** | **0.79** | **0.10** |
| 2 | 0.0001 | 64 | 200 | 0.88 | 0.02 | 0.76 | 0.12 |
| 3 | 0.0001 | 64 | 100 | 0.84 | 0.02 | 0.76 | 0.12 |
| 4 | 0.0001 | 128 | 100 | 0.88 | 0.02 | 0.76 | 0.10 |
| 5 | 0.0001 | 64 | 300 | 0.91 | 0.01 | 0.76 | 0.12 |
| 6 | 0.0001 | 256 | 50 | 0.88 | 0.03 | 0.76 | 0.11 |
| 7 | 0.0005 | 64 | 50 | 0.87 | 0.03 | 0.75 | 0.12 |
| 8 | 0.0001 | 64 | 50 | 0.82 | 0.02 | 0.75 | 0.14 |
| 9 | 0.0001 | 256 | 100 | 0.93 | 0.01 | 0.75 | 0.13 |
| 10 | 0.0005 | 128 | 50 | 0.93 | 0.02 | 0.74 | 0.13 |

**Figure S9:**
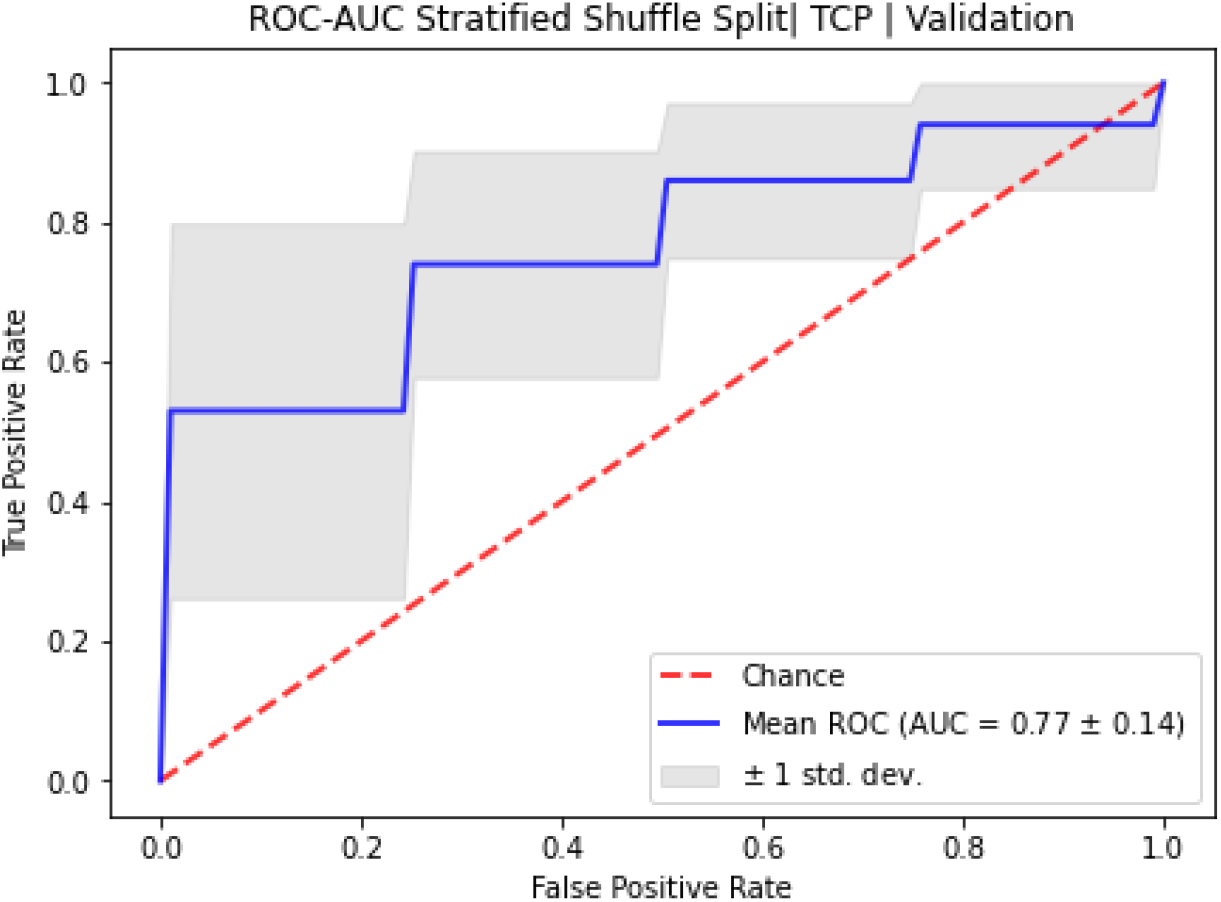
10-fold stratified shuffle 80-20 split ROC for NSCLC RTOE of TCP modeled with Single GNN architecture.

**Table S5:** Top 10 HP for **NSCLC NTCP (RP2)** with **Single GNN**

| HP # | Opt lr | Node | Epoch | Training AUC |  | Avg Validation AUC |  |
| --- | --- | --- | --- | --- | --- | --- | --- |
|  |  |  |  | Mean | Stdev | Mean | Stdev |
| **1** | **0.0001** | **64** | **50** | **0.77** | **0.04** | **0.75** | **0.13** |
| 2 | 0.0001 | 128 | 50 | 0.83 | 0.04 | 0.72 | 0.17 |
| 3 | 0.0001 | 64 | 200 | 0.86 | 0.03 | 0.67 | 0.18 |
| 4 | 0.0001 | 64 | 100 | 0.82 | 0.03 | 0.67 | 0.18 |
| 5 | 0.0001 | 128 | 100 | 0.88 | 0.03 | 0.66 | 0.18 |
| 6 | 0.0001 | 256 | 50 | 0.87 | 0.02 | 0.64 | 0.18 |
| 7 | 0.0005 | 64 | 50 | 0.88 | 0.03 | 0.64 | 0.17 |
| 8 | 0.005 | 128 | 200 | 0.99 | 0.02 | 0.63 | 0.18 |
| 9 | 0.0001 | 64 | 300 | 0.91 | 0.03 | 0.62 | 0.16 |
| 10 | 0.001 | 64 | 50 | 0.93 | 0.02 | 0.62 | 0.18 |

**Figure S10:**
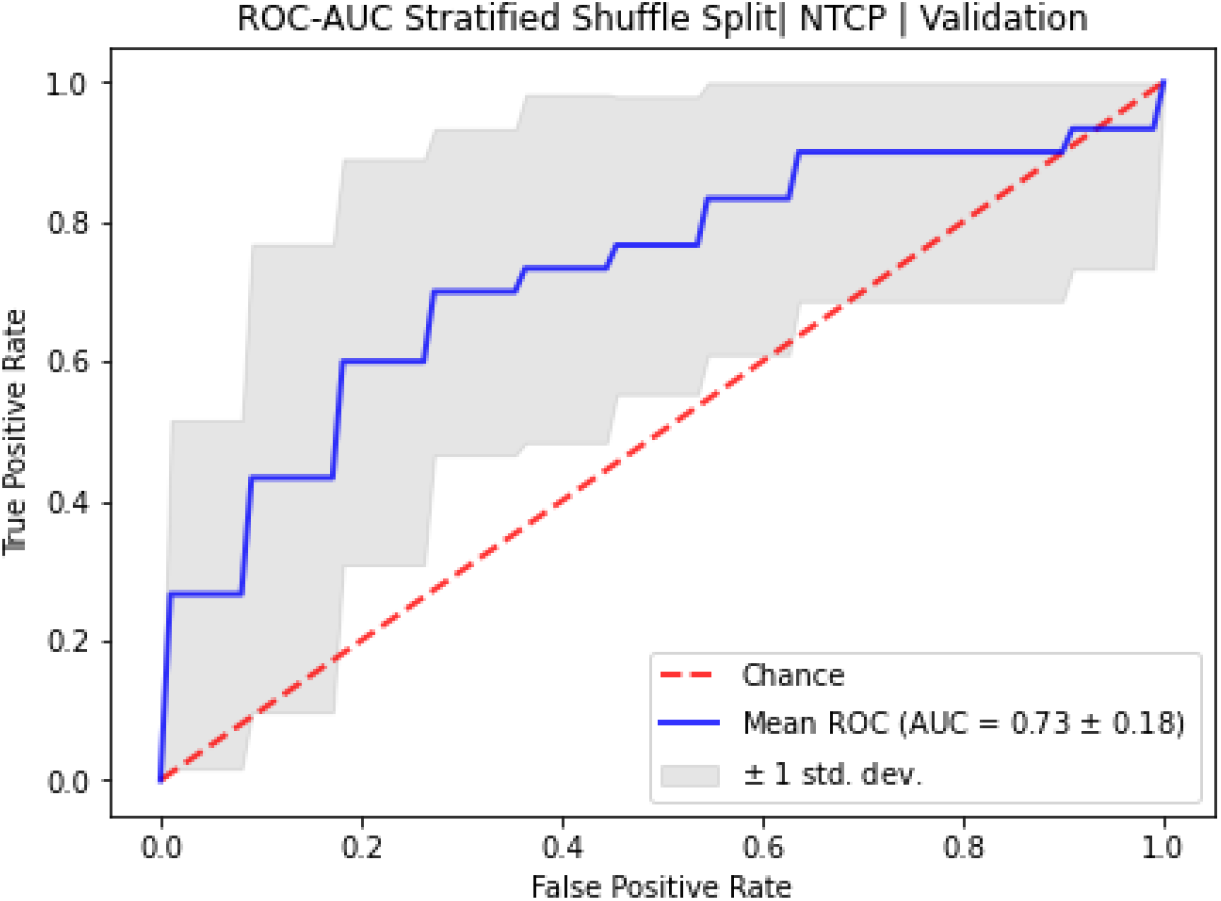
10-fold stratified shuffle 80-20 split ROC for NSCLC RTOE of NTCP modeled with Single GNN architecture.

**Table S6:**
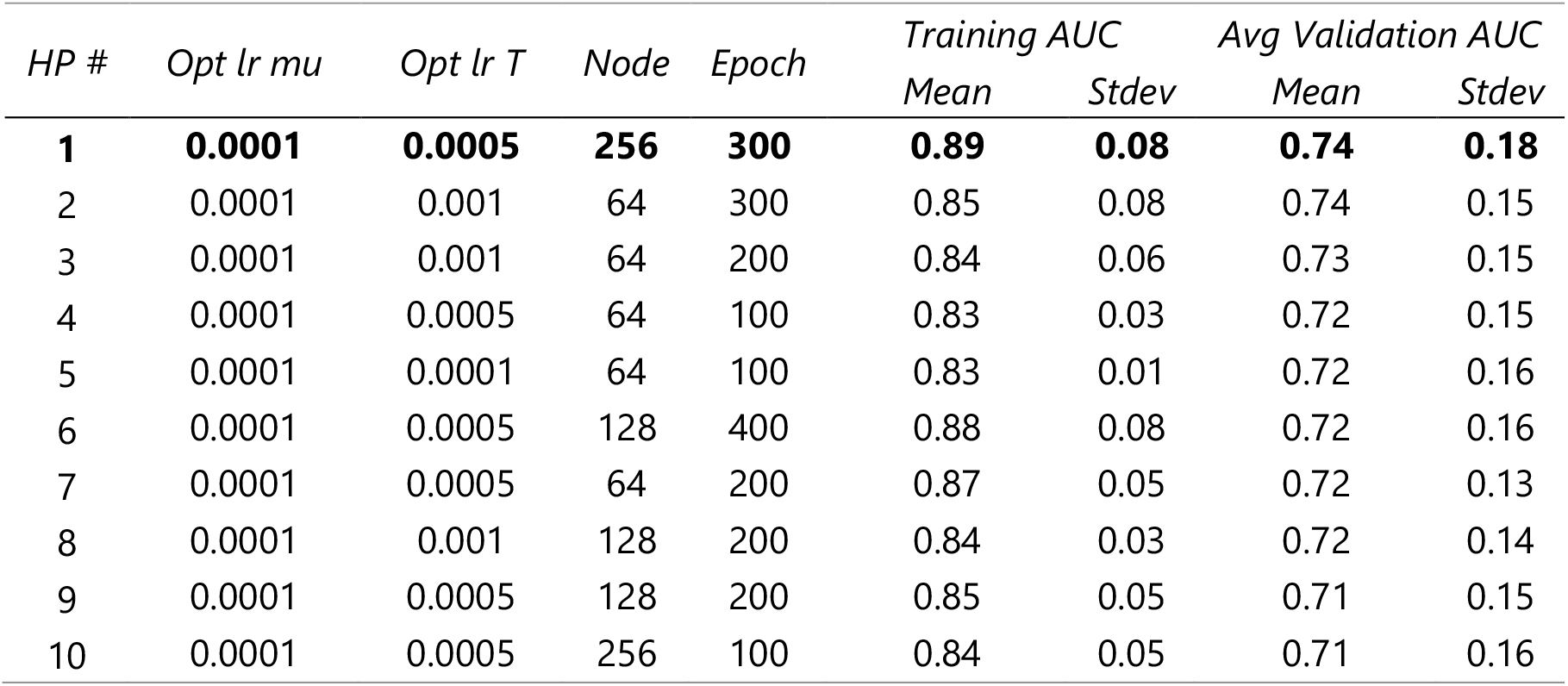
Top 10 HP for **NSCLC TCP (LC)** with **GLoGD-GNN**

**Figure S11:**
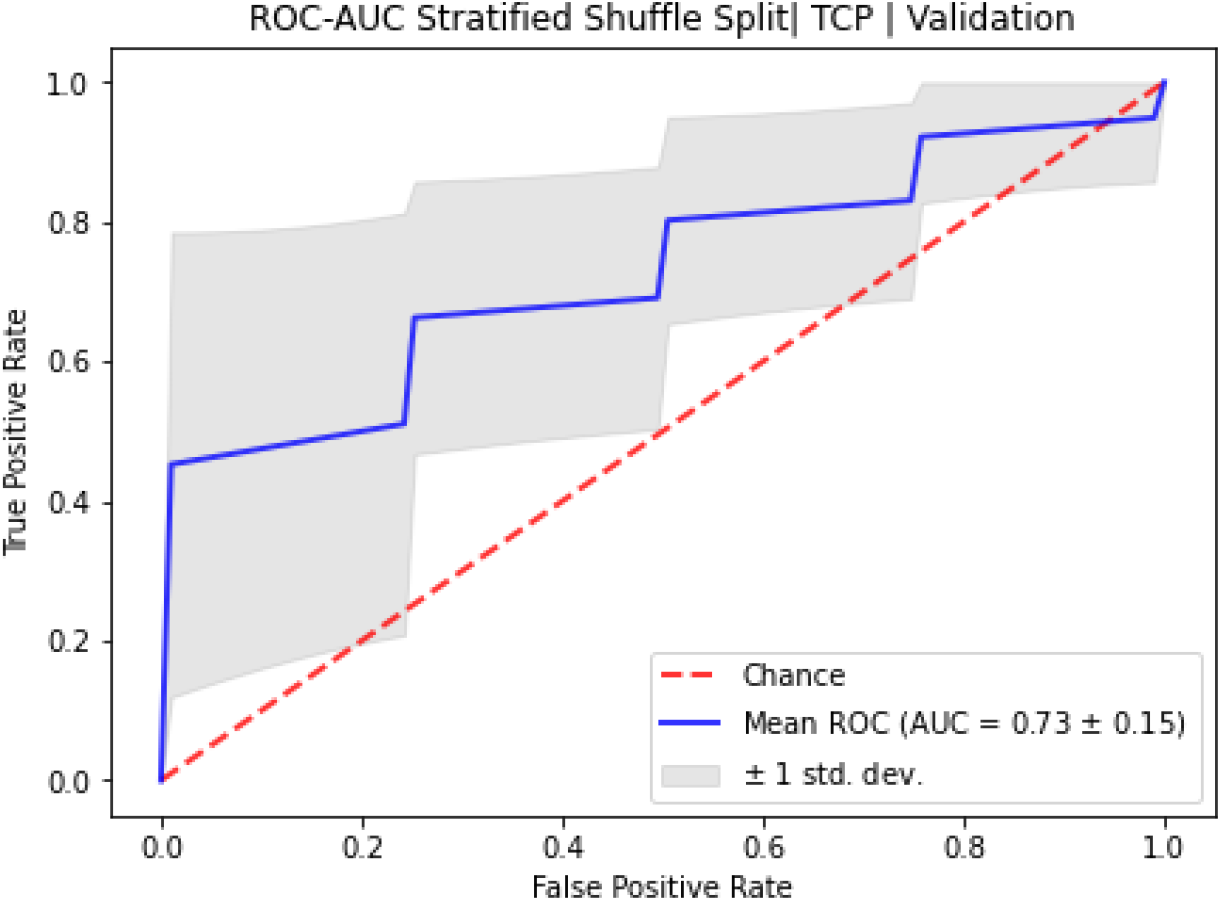
10-fold stratified shuffle 80-20 split ROC for NSCLC RTOE of TCP modeled with GLoGD-GNN architecture.

**Table S7:** Top 10 HP for **NSCLC NTCP (RP2)** with **GLoGD-GNN**

| HP # | Opt lr mu | Opt lr T | Node | Epoch | Training AUC |  | Avg Validation AUC |  |
| --- | --- | --- | --- | --- | --- | --- | --- | --- |
|  |  |  |  |  | Mean | Stdev | Mean | Stdev |
| **1** | **0.0001** | **0.001** | **128** | **100** | **0.84** | **0.05** | **0.80** | **0.13** |
| 2 | 0.0001 | 0.001 | 256 | 400 | 0.89 | 0.04 | 0.78 | 0.17 |
| 3 | 0.0001 | 0.001 | 256 | 300 | 0.85 | 0.11 | 0.76 | 0.19 |
| 4 | 0.0001 | 0.0001 | 256 | 400 | 0.88 | 0.07 | 0.76 | 0.19 |
| 5 | 0.0001 | 0.001 | 64 | 100 | 0.85 | 0.05 | 0.75 | 0.16 |
| 6 | 0.0001 | 0.0005 | 256 | 400 | 0.85 | 0.11 | 0.74 | 0.16 |
| 7 | 0.0001 | 0.0005 | 256 | 300 | 0.84 | 0.11 | 0.74 | 0.19 |
| 8 | 0.0001 | 0.0005 | 128 | 400 | 0.83 | 0.10 | 0.74 | 0.20 |
| 9 | 0.0001 | 0.001 | 256 | 100 | 0.84 | 0.08 | 0.74 | 0.16 |
| 10 | 0.0001 | 0.001 | 128 | 200 | 0.84 | 0.05 | 0.73 | 0.20 |

**Figure S12:**
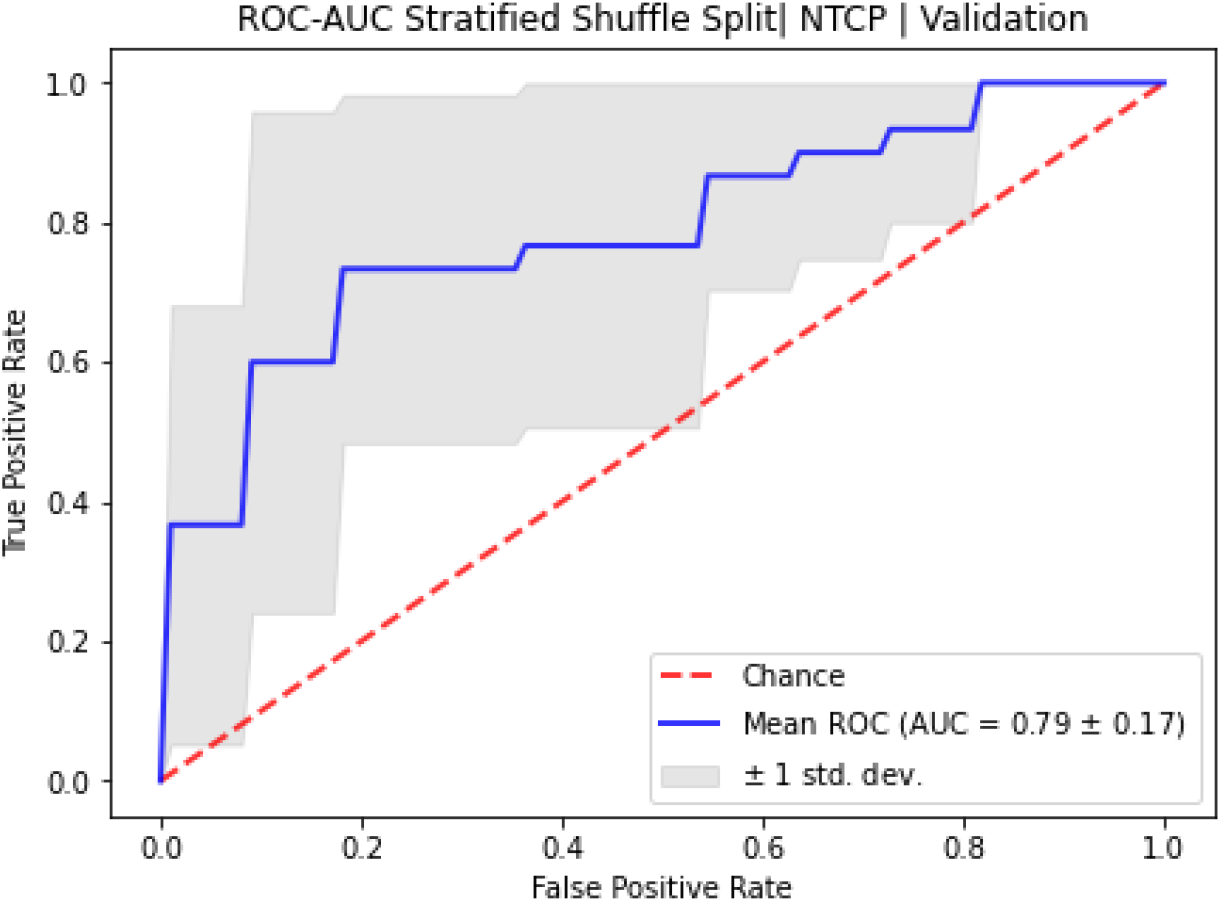
10-fold stratified shuffle 80-20 split ROC for NSCLC RTOE of NTCP modeled with GLoGD-GNN architecture.

#### S7.4 Synthetic Patient Generation via WGAN-GP

**Figure S13:**
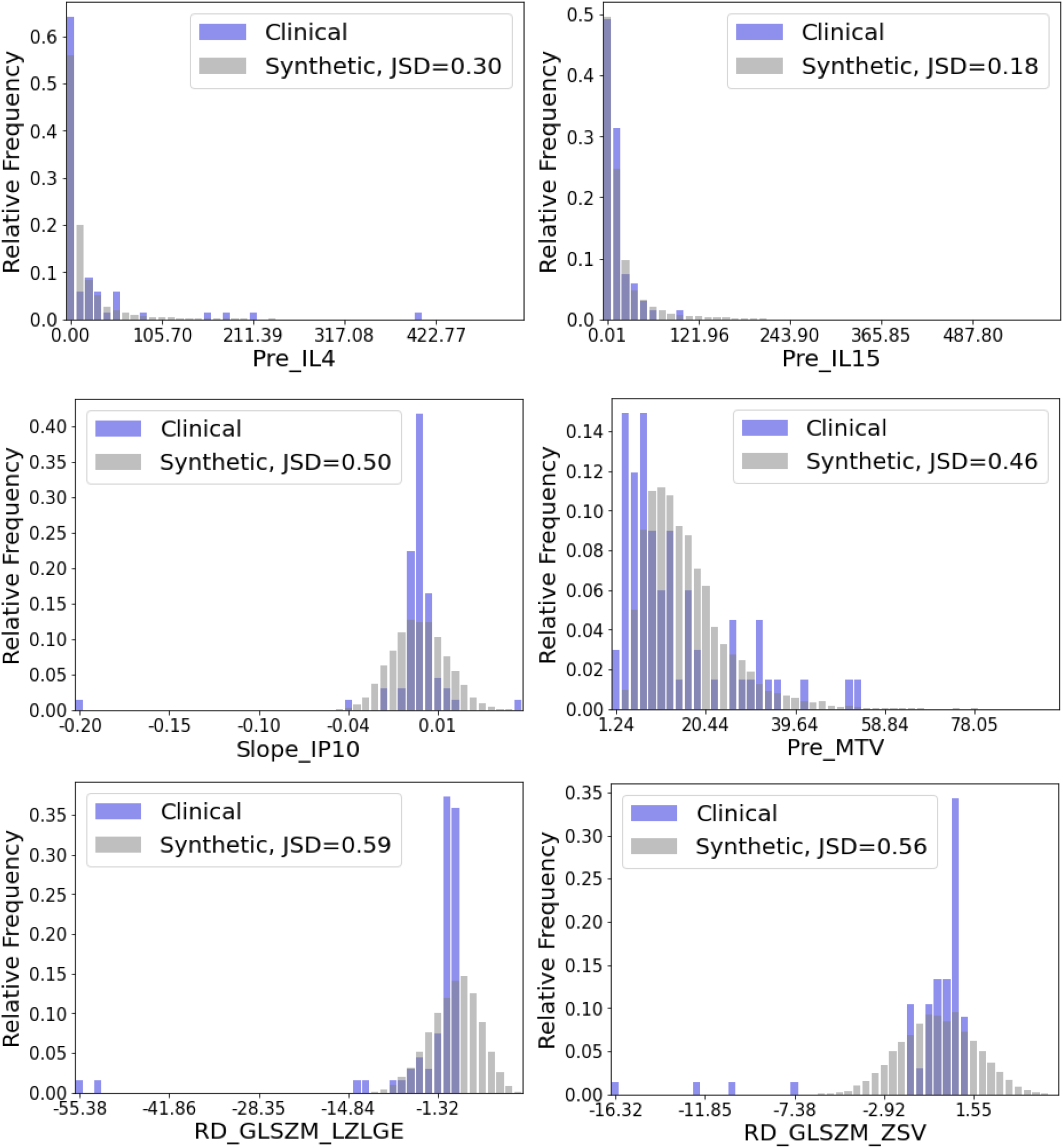
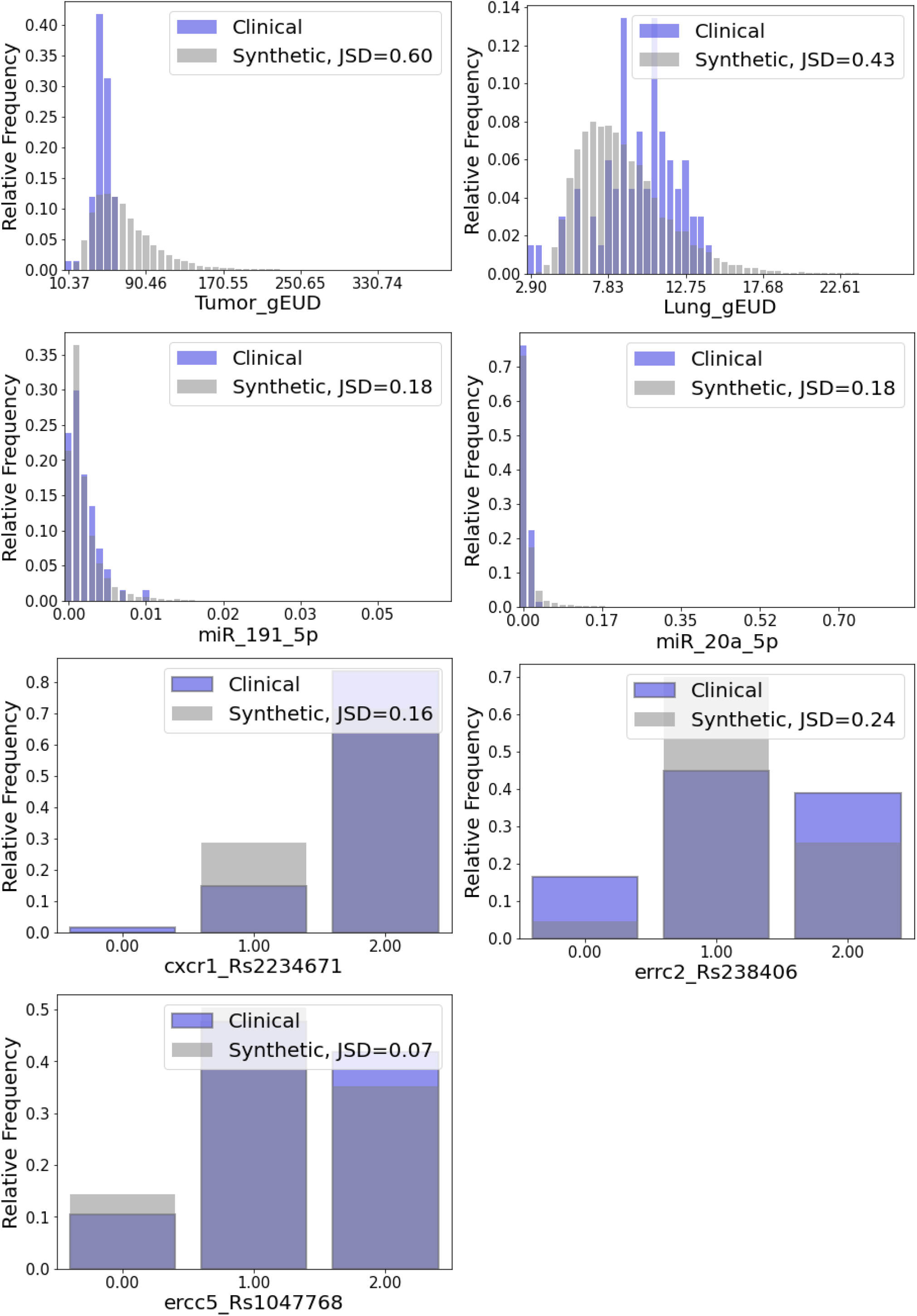
Distribution comparison of generated and original NSCLC dataset. Jensen Shannon Divergence metric between the distributions is provided for further insight on the differences.

#### S7.5 ODM Decision Analysis

We trained the models on only the synthetic data and set aside the entire clinical dataset for testing. To further test the generalizability, each model was trained with 4000 out of 10,000 randomly chosen synthetic patients. After learning, the models were tested on the clinical data. We compared two model architectures as listed in Table S8 in terms of Root Mean Square Difference (RMSD) and Mean Absolute Difference (MAD) calculated with respect to the reported clinical decisions. For a level comparison, all architectures were trained under identical conditions.

**Table S8:** ODM Decision Analysis Results for NSCLC

| **Error**<br>**[Gy/frac]** | **Overall (n=67)** |  | **Positive Clinical**<br>**Outcome (n=33 (49%))** |  | **Negative Clinical**<br>**Outcome (n=34 (51%))** |  |
| --- | --- | --- | --- | --- | --- | --- |
|  | RMSD | MAD | RMSD | MAD | RMSD | MAD |
| **Single GNN RTOE**<br>**+ DDQN ODM** | 0.97 ± 0.12 | 0.85 ± 0.11 | 0.96 ± 0.11 | 0.85 ± 0.11 | 0.97 ± 0.12 | 0.84 ± 0.11 |
| **GloGD GNN ROTE**<br>**+ DDQN ODM** | 0.61 ± 0.03 | 0.51 ± 0.03 | 0.66 ± 0.02 | 0.58 ± 0.02 | 0.55 ± 0.05 | 0.43 ± 0.04 |

\*Error ± SEM | Error between average recommendation and clinical decisions| ensemble of 5 models

| **Self-Evaluation**<br>**[Count (%)]** | **Overall (n=67)** |  |  | **Positive Clinical**<br>**Outcome (n=33 (49%))** |  | **Negative Clinical**<br>**Outcome (n=34 (51%))** |  |
| --- | --- | --- | --- | --- | --- | --- | --- |
|  | Good | Bad | Not Sure | Good | Not Sure | Good | Bad |
| **Single GNN RTOE +**<br>**DDQN ODM** | 26 (39%) | 14 (21%) | 27 (40%) | 6 (18%) | 27 (82%) | 20 (59%) | 14 (41%) |
| **GloGD GNN ROTE +**<br>**DDQN ODM** | 37 (55%) | 9 (13%) | 21 (31%) | 12 (36%) | 21 (64%) | 25 (74%) | 9 (26%) |

**Figure S14:**
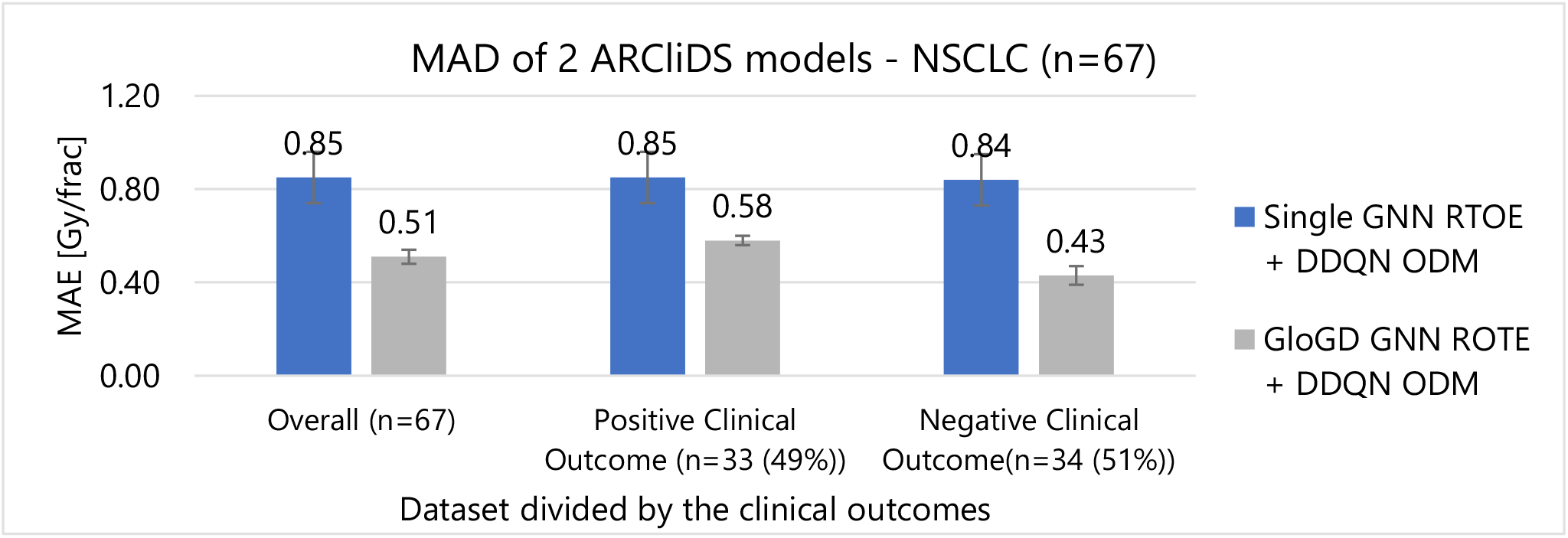
Mean Absolute Difference (MAD) of ARCliDS’s two model architecture for NSCLC patients grouped together according to the outcomes.

##### S7.5.1 DDQN trained on Single GNN RTOE - NSCLC

**Figure S15:**
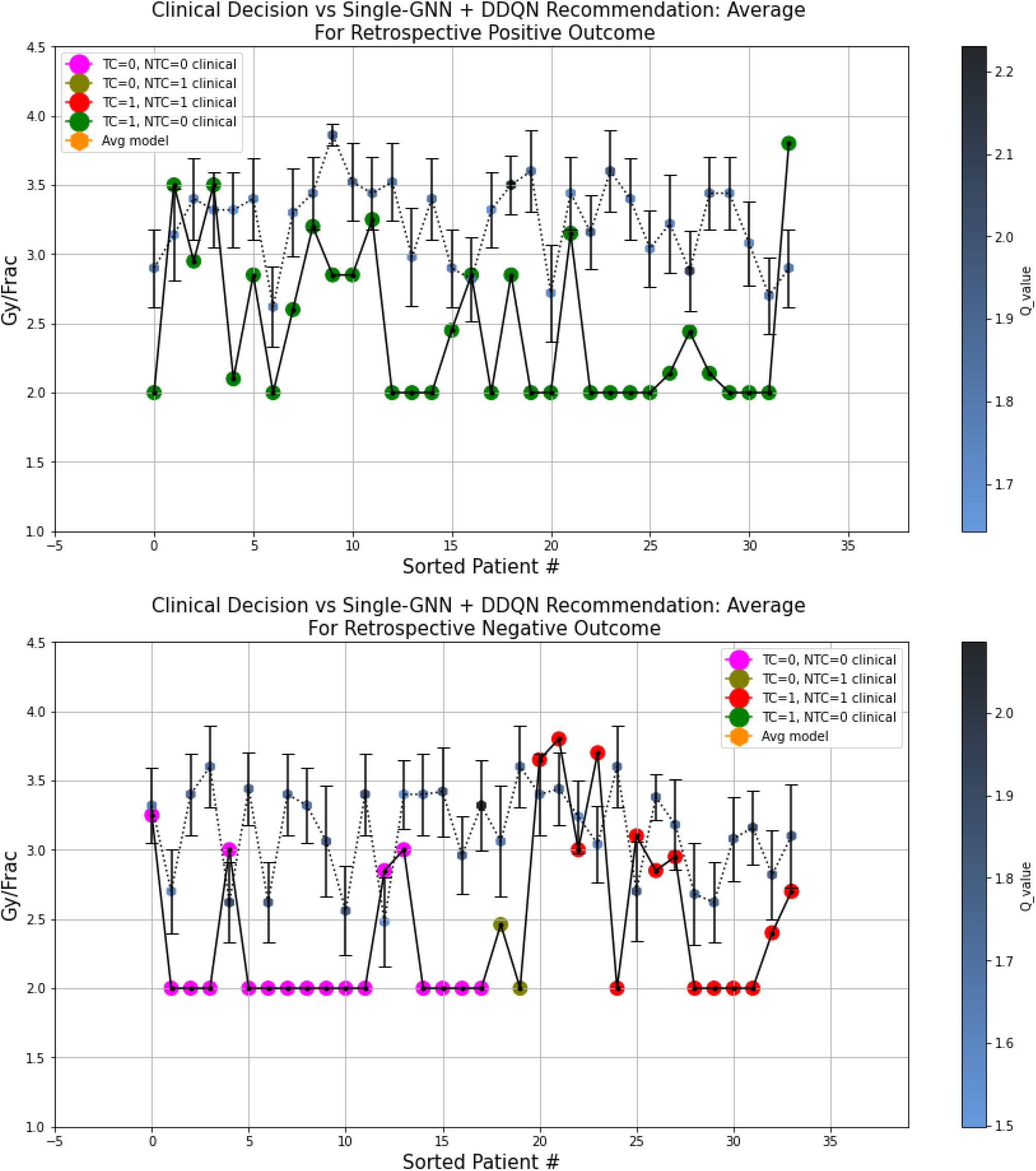
A visual comparison between the AI recommendation generated by the Single-GNN RTOE + DDQN ODM architecture and clinical decision for 2 groups of NSCLC patients divided according to the clinical outcomes. The clinical decisions are color coded with the outcomes and the ARCliDS recommendations are color coded with the respective q-value. Qualitatively, the q-value can be considered as the AI confidence in its recommendations.

##### S7.5.2 DDQN trained on GLoGD-GNN RTOE- NSCLC

**Figure S16:**
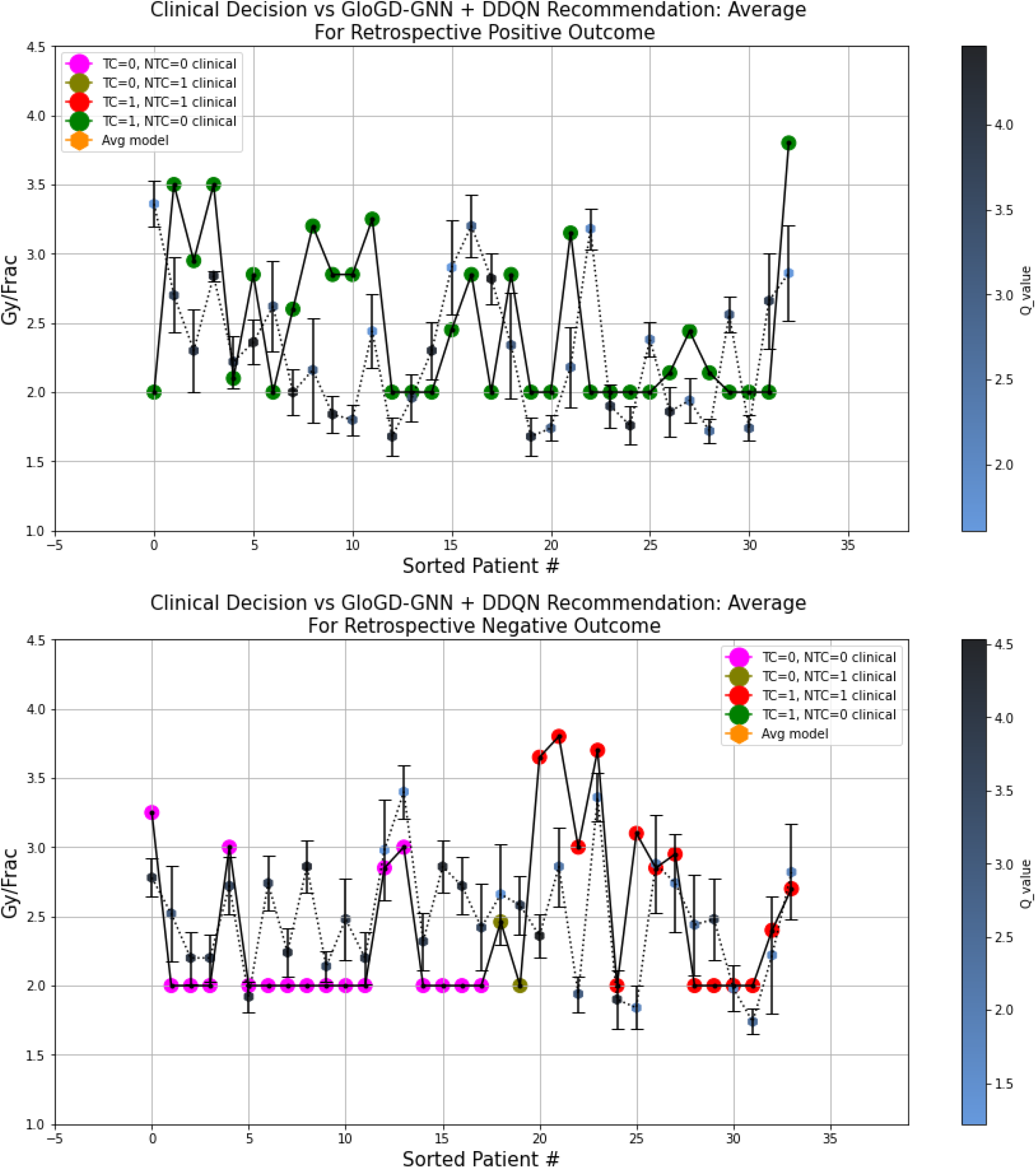
A visual comparison between the AI recommendation generated by the GLoGD-GNN RTOE + DDQN ODM architecture and clinical decision for 2 groups of NSCLC patients divided according to the clinical outcomes. The clinical decisions are color coded with the outcomes and the ARCliDS recommendations are color coded with the respective q-value. Qualitatively, the q-value can be considered as the AI confidence in its recommendations.

### Use Case 2: HCC

**Figure S17:**
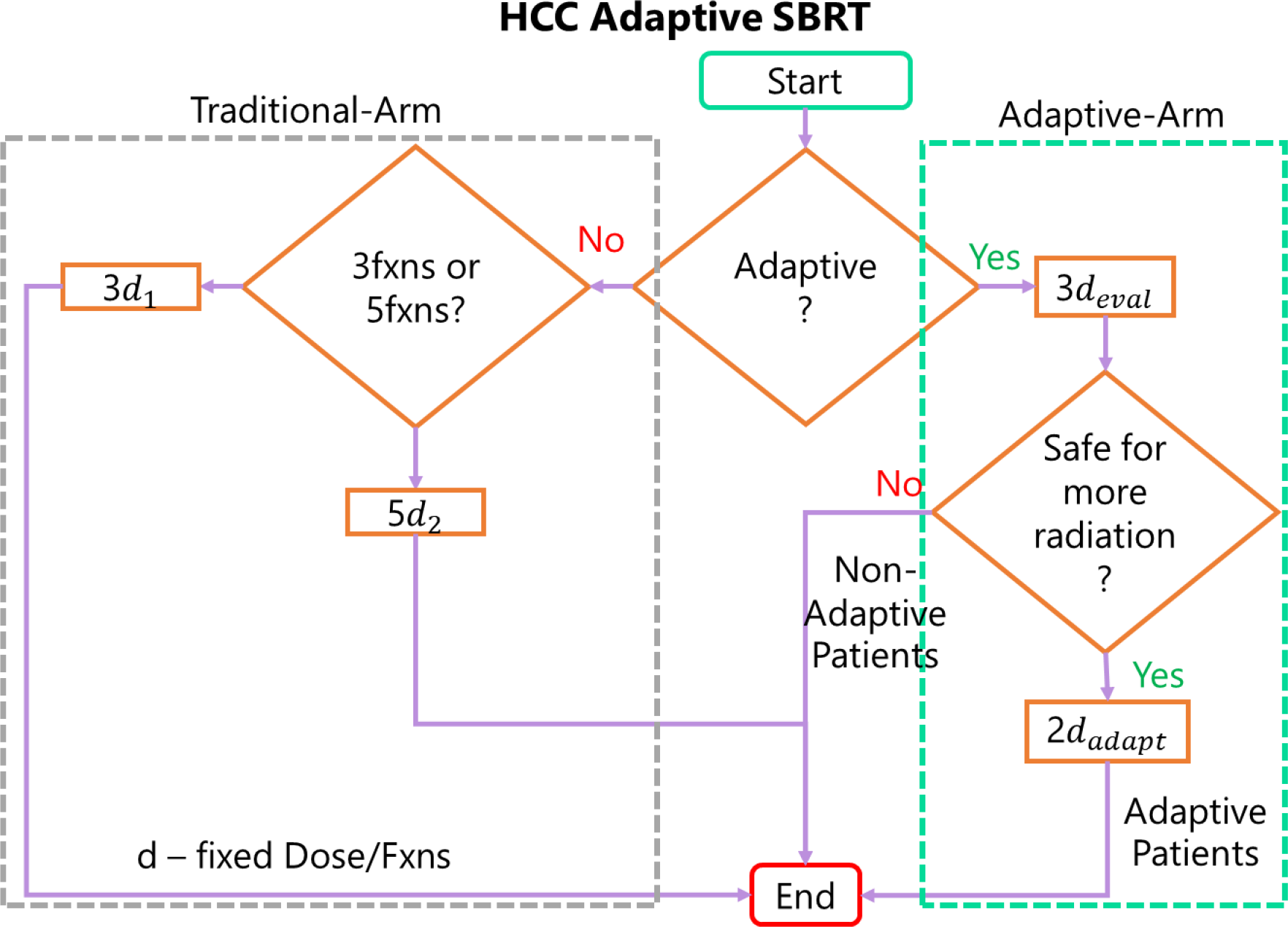
Decision-Making in HCC KBR-ASBRT. At first, patients are evaluated for the best course of treatment. Those that are not fit for adaptive SBRT receive traditional SBRT. The patients are further evaluated on whether they should receive the treatment in 3 or 5 fractions. The patients selected for Adaptive SBRT receive the first three fractions followed by a 1-month gap. After analyzing the trend in the multi-omics information, those deemed safe for further radiation recieves an adaptive dose (*d*_*adapt*_) for 2 more fractions.

As shown in the Figure S17, decision making in the Adaptive Arm is a two-step process. First, the patient is given 3 daily dose fractions and evaluated if they are safe for more radiation. Second, for those that are safe, there is a question of what the optimal adaptive dose should be. The first decision can be decided by an RTOE, while the second decision by another RTOE and an ODM.

The Non-Adaptive RTOE is trained with input from non-adaptive patient’s pre and mid treatment information and label from their outcome. The Adaptive RTOE is trained on the adaptive patient’s dataset.

#### S8.1 Data Description

Information on 292 patients with 360 tumor sites were available. The greater number of tumor sites is due to multiple tumor or recurrence. Out of that, 81 patients with 104 tumor sites had dense multi-omics data available and 71 patients with 99 tumor sites had complete information.

**Table S9:** HCC Patient Characteristics

| Variable | Category | Patient Count (n = 71) |
| --- | --- | --- |
| Sex | Male | 56 |
|  | Female | 15 |
| Age | Median (Q1–Q3) | 65 (59–75) |
|  | Range | 34–85 |
| Pre-Treatment Cirrhosis | Yes | 64 |
|  | No | 7 |
| Portal Vein Thrombosis | Yes | 12 |
|  | No | 59 |
| Pre-Treatment Number of Active Lesions | Median (Q1–Q3) | 1 (1–1) |
|  | Range | 1–4 |

**Table S10:** HCC Patient count classified by the treatment outcome

| Adaptive-Arm | Count<br>HCC<br>(n = 99) | Count<br>HCC –Non-Adaptive<br>(n = 36) | Count<br>HCC –Adaptive<br>(n = 64) |
| --- | --- | --- | --- |
| LC = 0 | 4 | 3 | 1 |
| LC = 1 | 95 | 32 | 63 |
| Adaptive-Arm | Count<br>HCC<br>(n = 71) | Count<br>HCC –Non-Adaptive<br>(n = 30) | Count<br>HCC –Adaptive<br>(n = 41) |
| LT = 0 | 54 | 20 | 34 |
| LT = 1 | 17 | 10 | 7 |
| LC: local control<br>LT: Liver Toxicity ($\geq 2$ points increase in Child Pugh Score during any point in the treatment.)<br>Total patients = 71, Non-adaptive Patient = 30, Adaptive Patients = 41,<br>For LC endpoints, patients with multiple tumor site were considered as different data points, Non-adaptive Datapoints = 36, Adaptive Datapoints = 64.<br>For LT endpoint, each patient was considered as one data point. | | | |

For the training of Adaptive RTOE, we added the 3 non-adaptive patients to the list of adaptive patients to increase the LC=0 count. To train RTOE for LC, the data from patients with multiple tumor sites were considered as different datapoints. Due to small sample size of non-adaptive patients, we considered only the second decision-making problem for the adaptive patients.

**Figure S18:**
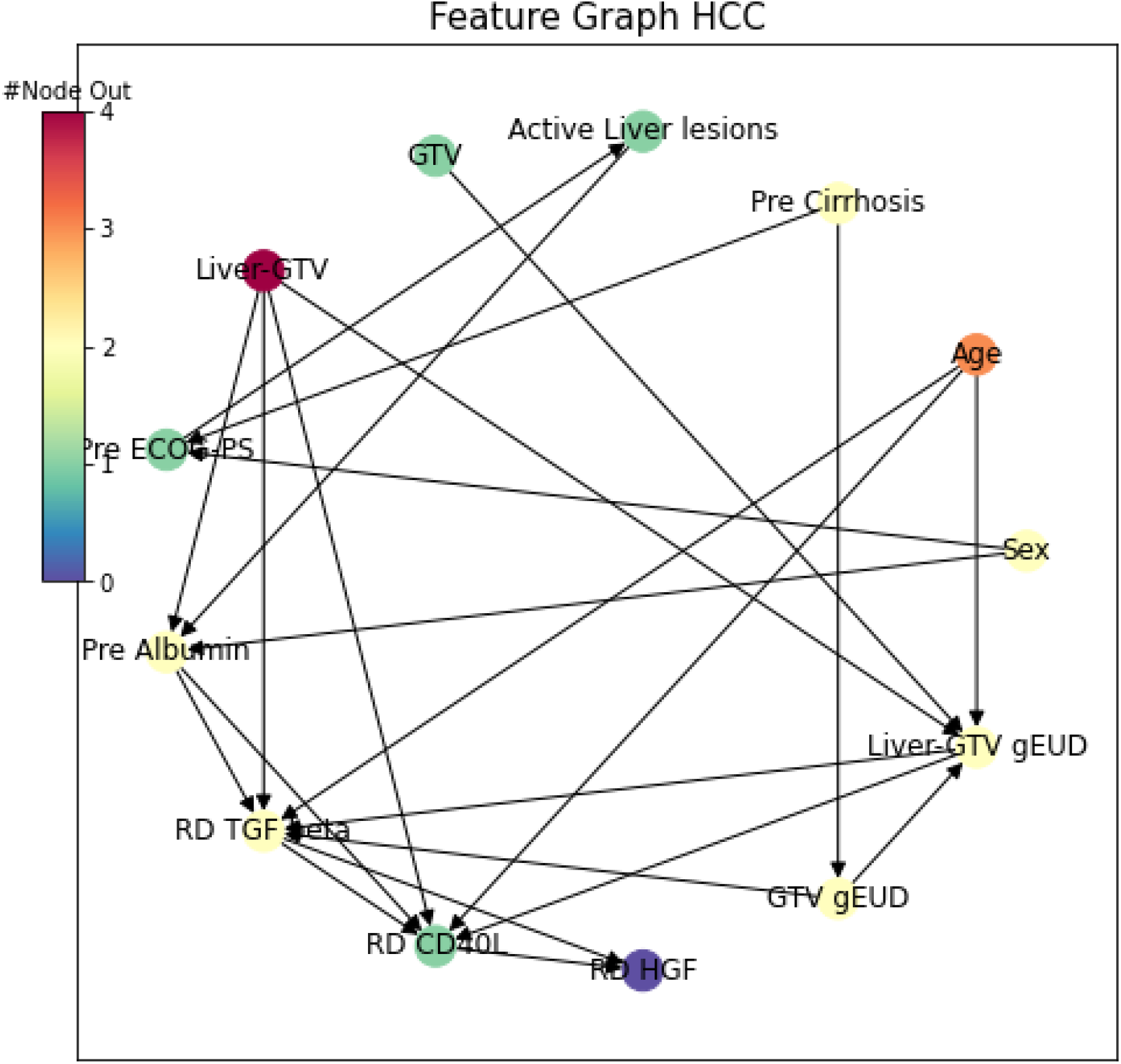
Directed graph showing the inter-relation between the HCC patient’s features. The nodes, which represent features, are color coded with the number of outgoing relationships. Pre stands for pre-treatment observation, RD and slope stands for relative difference and change in feature value between pre-treatment and mid-treatment observation, respectively.

**Figure S19:**
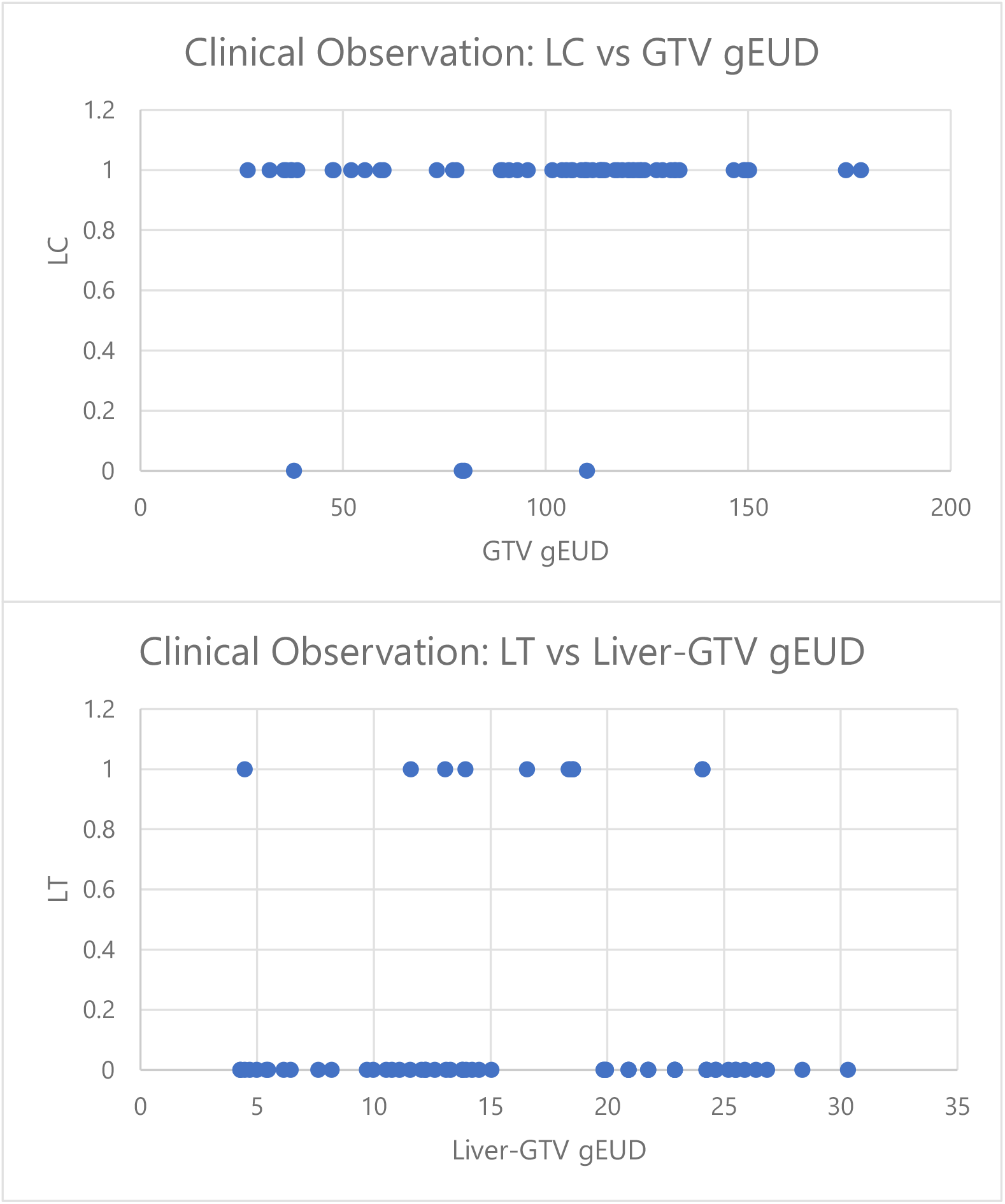
Plots showing HCC population GTV gEUD vs observed local control and Liver-GTV gEUD vs observed liver toxicity (LT). This plot captures inter-patient heterogeneity which shows patient’s diverse treatment response. Note that, for population trend for LT is opposite to expected trend for an individual patient, i.e., patients with highest Liver-GTV gEUD is not showing toxicity.

**Table S11:** HCC Patients’ feature description.

| Patient Variable | Biological/Clinical Characteristics |
| --- | --- |
| **Clinical Factors** |  |
| **Sex** | Biological male/female |
| **Age** | Pre-treatment Age |
| **Cirrhosis** | Late stage of scarring (fibrosis) of the liver |
| **ECOG-PS** | Eastern Cooperative Oncology Group Performance Status: ECOG-PS is a scale for assessing the level of function and capability of self-care. It ranges from integer 0 to 4 where PS0 indicates fully active patients while PS4 indicates patients that are completely unable for physical activity and self-care. |
| **Active Liver Lesions** | Abnormal clumps of cells in the liver |
| **Albumin** | A globular protein produced in the liver that circulates throughout human body via plasma. Its synthesis is stimulated by hormones such as insulin and inhibited by pro-inflammatory substance such as IL6 and TNF-α.<sup>18</sup> |
| **Tumor PET Imaging** |  |
| **GTV** | Gross tumor volume is the volume of the actual tumor. |
| **Liver-GTV** | Liver volume minus GTV. This provides the volume of the normal liver tissue. |
| **Dosimetry** |  |
| **GTV gEUD** | Generalized equivalent uniform dose (gEUD) of GTV converted from EQD2 (Equivalent Dose at standard 2 Gy per fraction) dose distribution using the Linear-Quadratic-Linear model. The model parameters used are $\alpha/\beta = 10$ Gy, $a=-20$, and $D_T=20$ Gy.<sup>11</sup> |
| **Liver-GTV gEUD** | Generalized equivalent uniform dose (gEUD) of liver volume minus GTV converted from EQD2 (Equivalent Dose at standard 2 Gy per fraction) dose distribution using the Linear-Quadratic-Linear model. The model parameters used are $\alpha/\beta = 2.5$ Gy, $a=1$, and $D_T=5$ Gy.<sup>11</sup> |
| **Cytokines/Signaling molecule** |  |
| **TGF-β** | Transforming growth factor beta is the prototype of TGF-β family that has diverse role in the control of cell proliferation and differentiation, wound healing and immune system, and pathology such as skeletal diseases, fibrosis, and cancer.<sup>19</sup> |
| **CD40L** | Cluster of Differentiation 40 receptor is a costimulatory molecule from the tumor necrosis factor receptor (TNF-R) family. CD40 binds with its ligand (CD40L) which is transiently expressed on T cells (immune cell) as well as other non-immune cells, under inflammatory conditions. This activates antigen presenting cells and other wide spectrum of molecular and cellular process.<sup>20</sup> |
| **HGF** | Hepatocyte growth factor is a pleiotropic cytokine required for development of several organs. It is produced after injury of the organ tissue and promotes tissue repair. HGF promotes tissue repair through inhibition of apoptosis of epithelial and endothelial cells, and by counteracting several pro-apoptotic and fibrosis factors such as TGF-β, IL-1β, IL8, TNF-α.<sup>21,22</sup> |

#### S8.2 GLoGD-GNN for Monotonic TCP/NTCP

**Figure S20:**
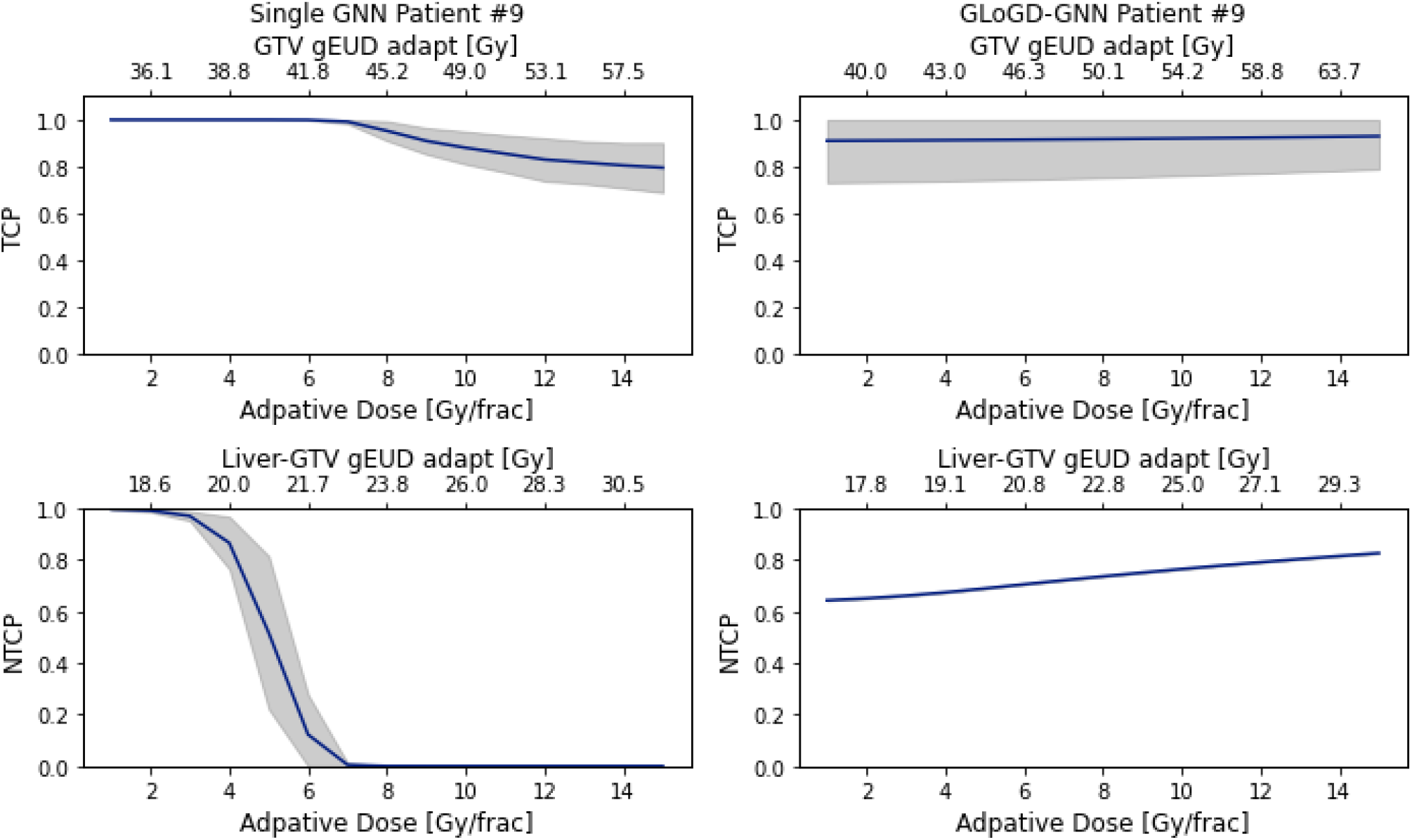
HCC outcomes estimate for adaptive dose ranging from 1 to 15 Gy/frac. The GLoGD-GNN correction successfully established a monotonic relationship for the NTCP. The model uncertainty is obtained from an ensemble of five RTOE model and presented as ±1 standard deviation. Note: the sample/patient IDs in the figures are arbitrary, unidentifiable, and meaningless and they were not known to anyone outside the research.

**Figure S21:**
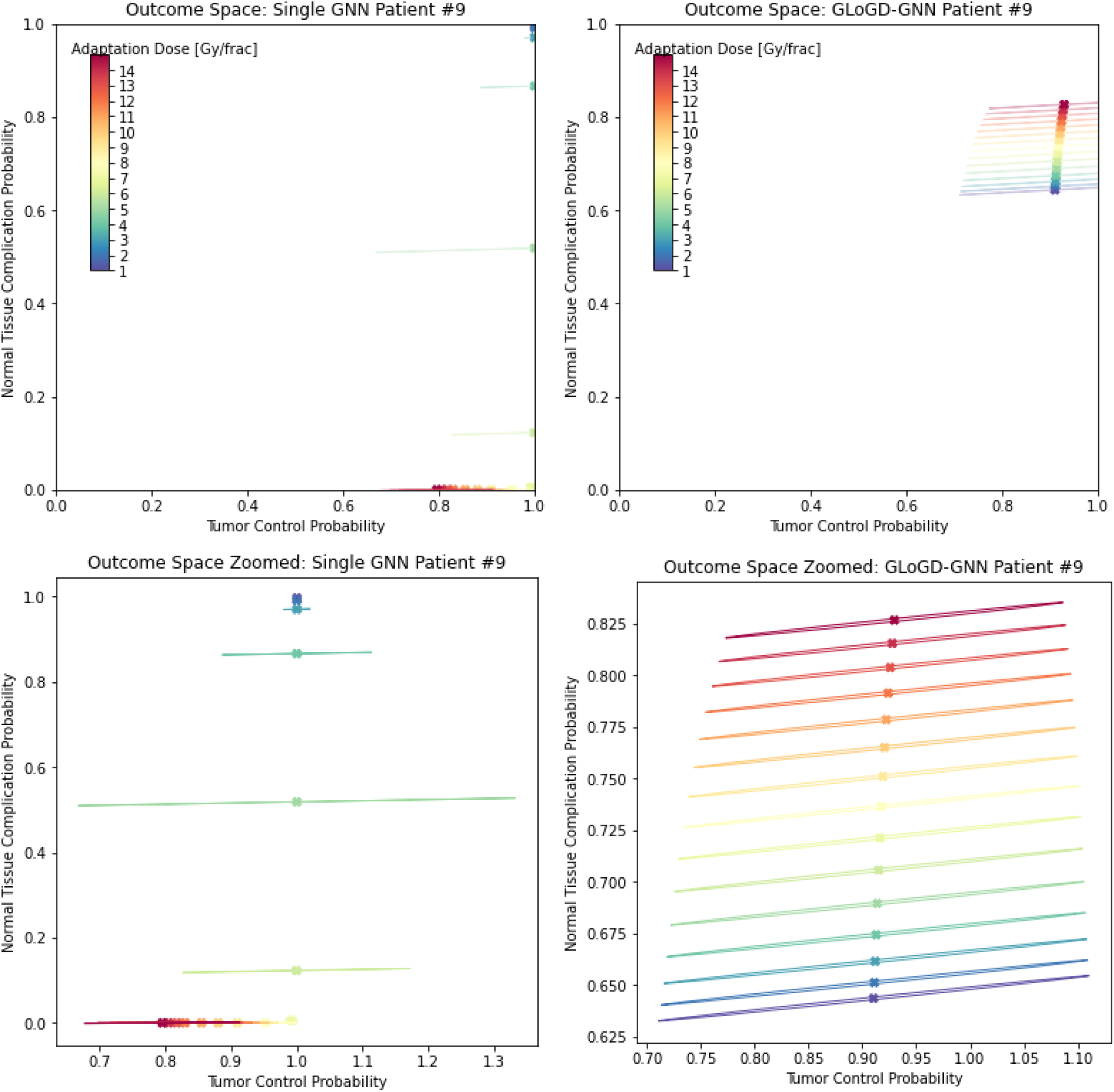
NSCLC outcomes estimate for adaptive dose ranging from 1 to 15 Gy/frac in the outcome space spanned by TCP and NTCP. Model uncertainty is obtained from an ensemble of five RTOE model and presented as an eclipse set by the Covariance matrix. In the left figure, NTCP is decreasing with increasing dose value. In the right figure, GLoGD-GNN correctly flipped the dose order i.e., NTCP is increasing with increasing dose value. Note: the sample/patient IDs in the figures are arbitrary, unidentifiable, and meaningless and they were not known to anyone outside the research.

#### S8.3 RTOE Hyper Parameter Tuning

Tables S12-S15 lists the top 10 performing HP for the HCC dataset followed by receivers operating curves (ROC) for the best performing HP (in bold) shown in Figures S22-S25. To check for the reproducibility, ROCs were generated by retraining the models with the best performing HP. The reported AUROCC values are in the mean±stdev format, where the mean AUROCC value is the area under the mean true positive rate curve, while the standard deviation is calculated from the AUROCC of 10 individual model output.

**Table S12:** Top 10 HP for **HCC Adaptive TCP** with **single-GNN**

| HP # | Opt lr | Node | Epoch | Training AUC |  | Avg Validation AUC |  |
| --- | --- | --- | --- | --- | --- | --- | --- |
|  |  |  |  | Mean | Stdev | Mean | Stdev |
| **1** | **0.005** | **64** | **200** | **0.99** | **0.00** | **0.76** | **0.31** |
| 2 | 0.0001 | 64 | 100 | 0.99 | 0.00 | 0.74 | 0.31 |
| 3 | 0.001 | 256 | 200 | 0.99 | 0.00 | 0.73 | 0.30 |
| 4 | 0.001 | 128 | 300 | 0.99 | 0.00 | 0.73 | 0.32 |
| 5 | 0.005 | 64 | 300 | 0.99 | 0.00 | 0.72 | 0.30 |
| 6 | 0.001 | 256 | 100 | 0.99 | 0.00 | 0.72 | 0.33 |
| 7 | 0.001 | 256 | 300 | 0.99 | 0.00 | 0.72 | 0.30 |
| 8 | 0.001 | 256 | 50 | 0.99 | 0.00 | 0.72 | 0.33 |
| 9 | 0.005 | 128 | 200 | 0.99 | 0.00 | 0.72 | 0.34 |
| 10 | 0.0005 | 256 | 300 | 0.99 | 0.00 | 0.71 | 0.32 |

**Figure S22:**
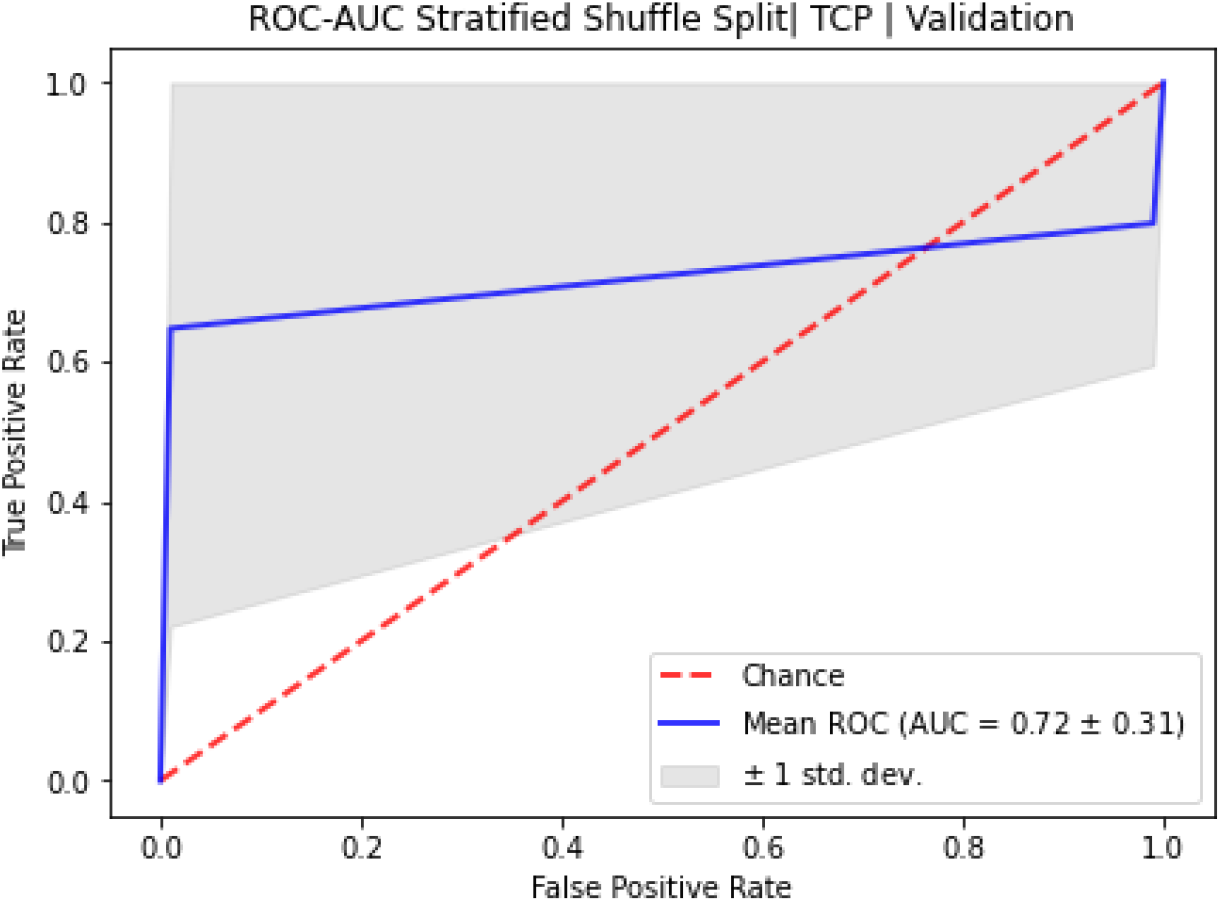
10-fold stratified shuffle 80-20 split ROC for HCC Adaptive RTOE of TCP modeled with single-GNN architecture. Note: High Validation AUC deviation and relatively flatter AUC curve is due to severe class imbalance.

**Table S13:**
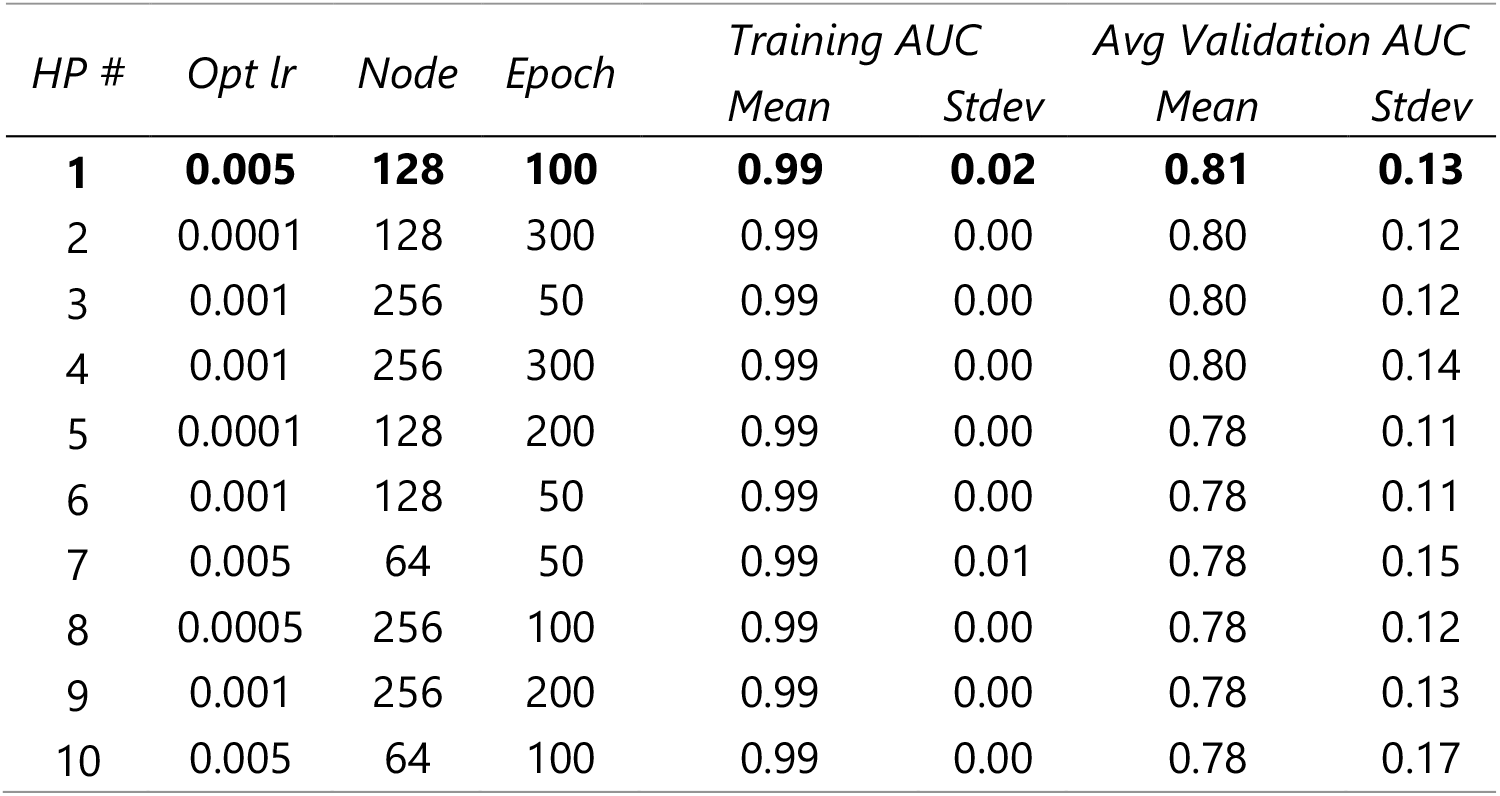
Top 10 HP for **HCC Adaptive NTCP** with **single-GNN**

**Figure S23:**
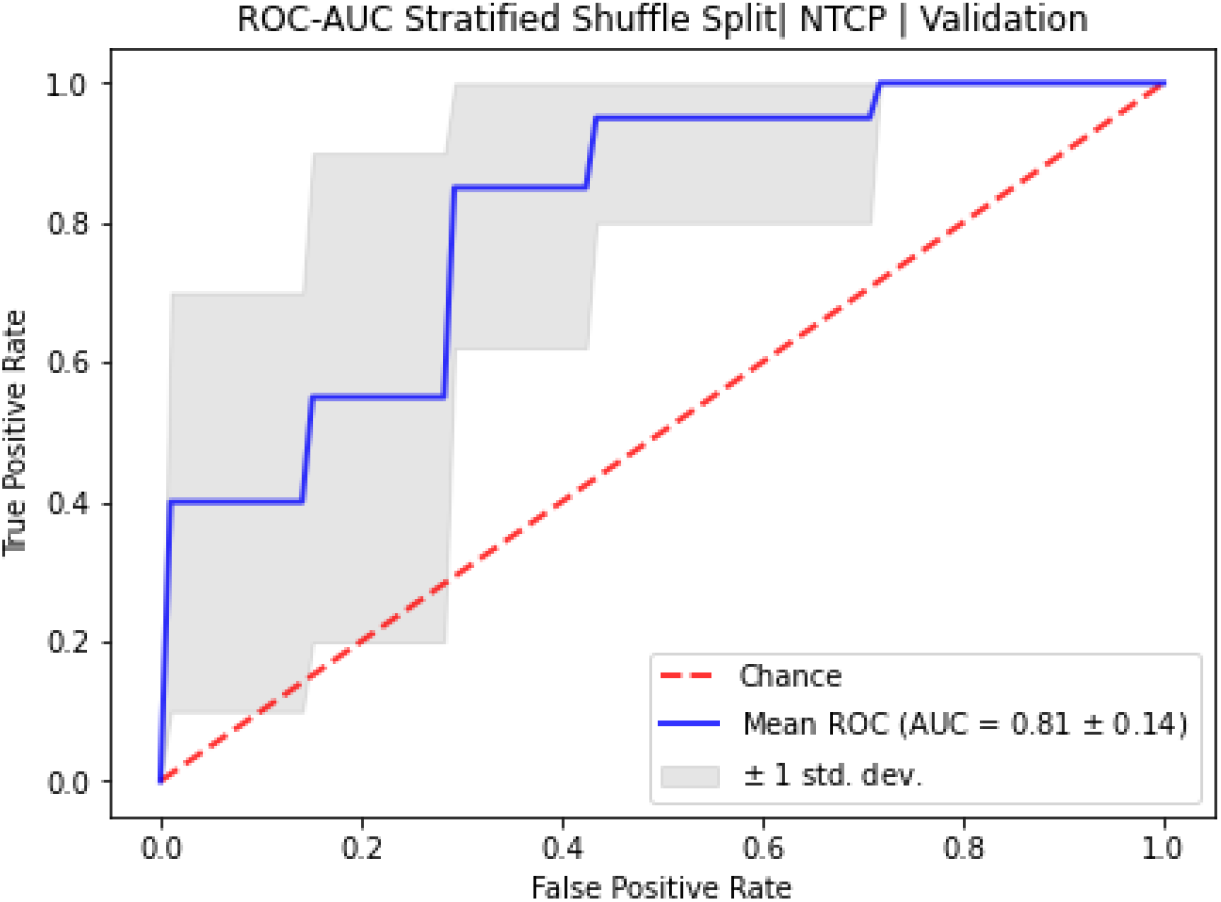
10-fold stratified shuffle 80-20 split ROC for HCC Adaptive RTOE of NTCP modeled with Single GNN architecture.

**Table S14:** Top 10 HP for **HCC Adaptive TCP** with **GloGD-GNN**

| HP # | Opt lr mu | Opt lr T | Node | Epoch | Training AUC |  | Avg Validation AUC |  |
| --- | --- | --- | --- | --- | --- | --- | --- | --- |
|  |  |  |  |  | Mean | Stdev | Mean | Stdev |
| **1** | **0.0005** | **0.0001** | **256** | **300** | **0.92** | **0.13** | **0.77** | **0.21** |
| 2 | 0.0005 | 0.0005 | 256 | 100 | 0.87 | 0.14 | 0.76 | 0.19 |
| 3 | 0.0001 | 0.0005 | 256 | 200 | 0.89 | 0.20 | 0.75 | 0.21 |
| 4 | 0.0001 | 0.001 | 256 | 400 | 0.85 | 0.22 | 0.73 | 0.21 |
| 5 | 0.0001 | 0.001 | 128 | 100 | 0.91 | 0.15 | 0.72 | 0.20 |
| 6 | 0.0001 | 0.0005 | 128 | 300 | 0.76 | 0.24 | 0.72 | 0.21 |
| 7 | 0.0001 | 0.001 | 128 | 200 | 0.81 | 0.21 | 0.71 | 0.22 |
| 8 | 0.0001 | 0.0005 | 256 | 300 | 0.94 | 0.15 | 0.71 | 0.21 |
| 9 | 0.0005 | 0.0001 | 64 | 200 | 0.99 | 0.00 | 0.71 | 0.28 |
| 10 | 0.0001 | 0.001 | 128 | 400 | 0.97 | 0.06 | 0.69 | 0.26 |

**Figure S24:**
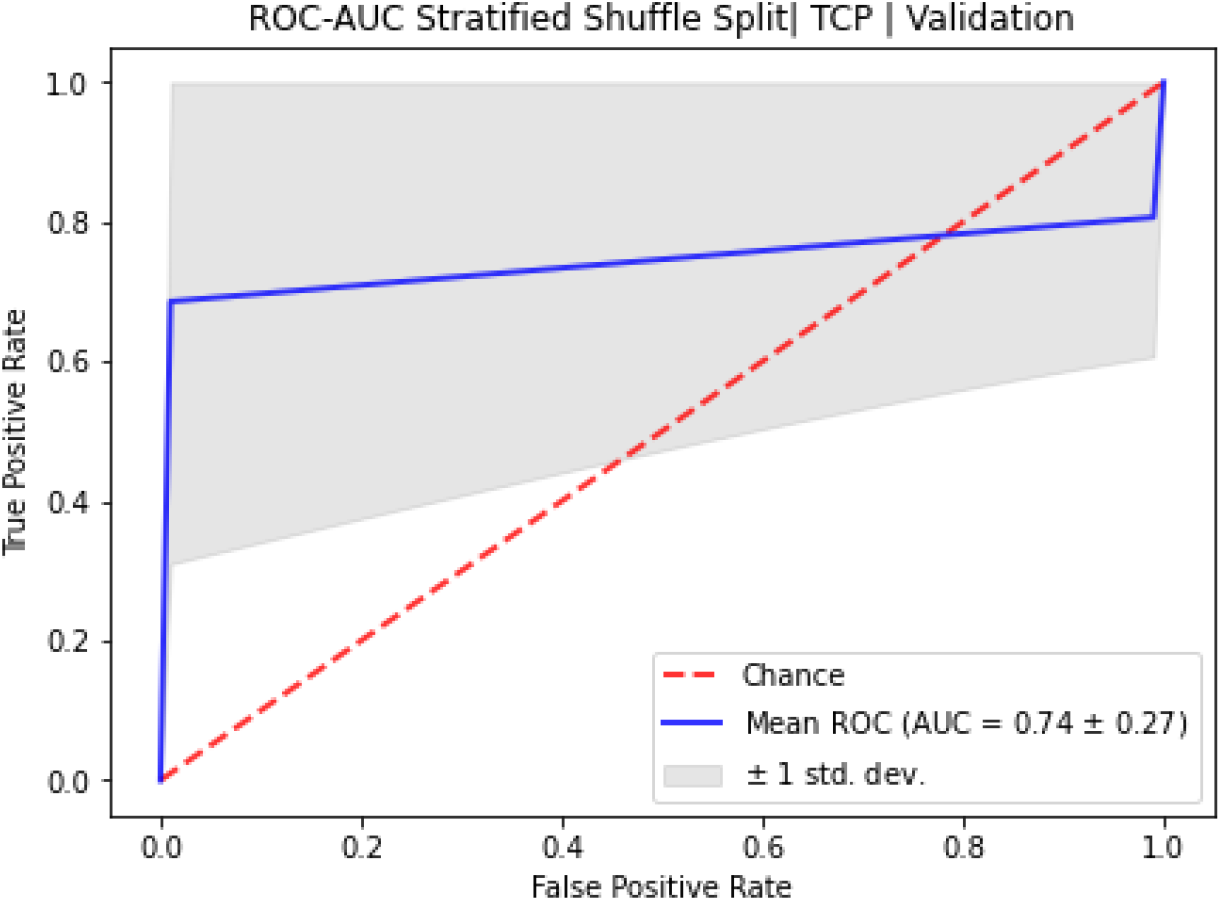
10-fold stratified shuffle 80-20 split ROC for HCC Adaptive RTOE of TCP modeled with GLoGD-GNN architecture. Note: High Validation AUC deviation and relatively flatter AUC curve is due to severe class imbalance.

**Table S15:**
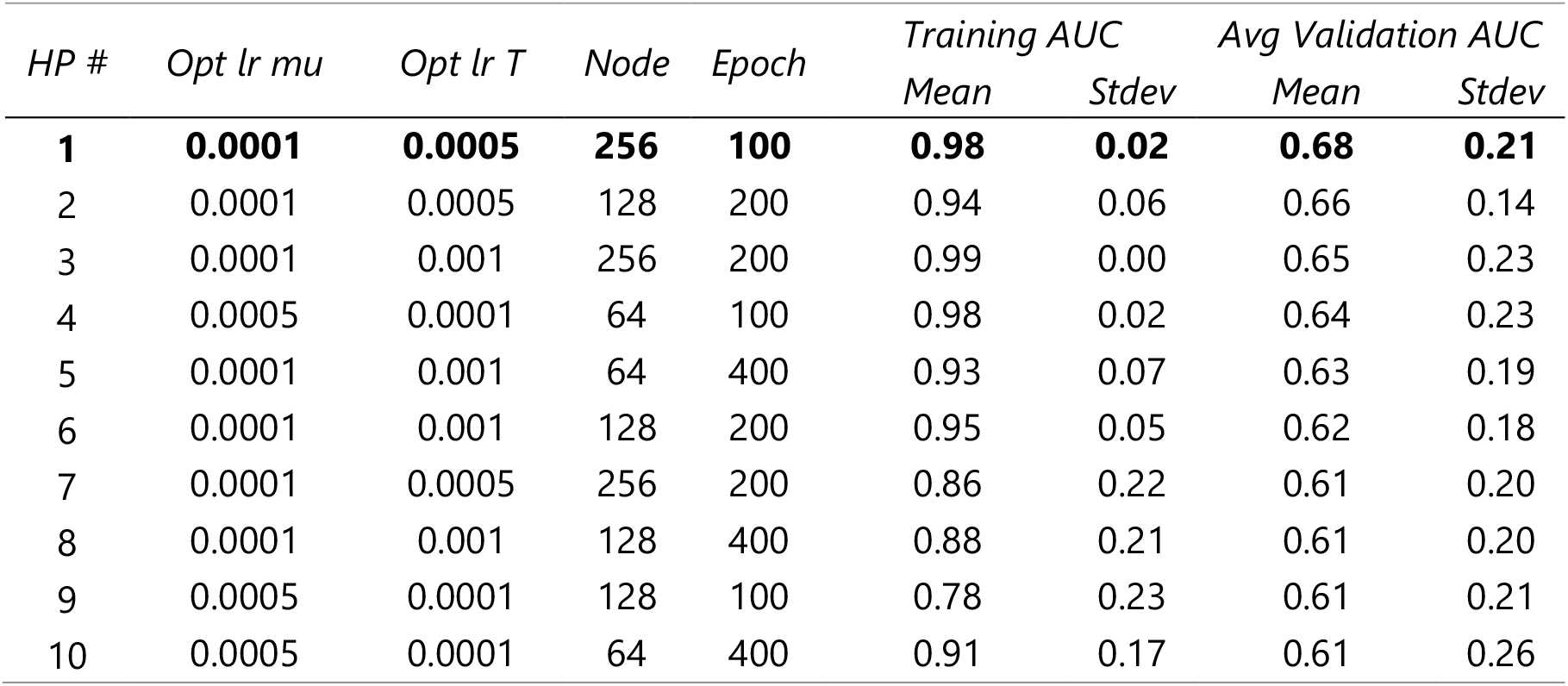
Top 10 HP for **HCC Adaptive NTCP** with **GloGD-GNN**

**Figure S25:**
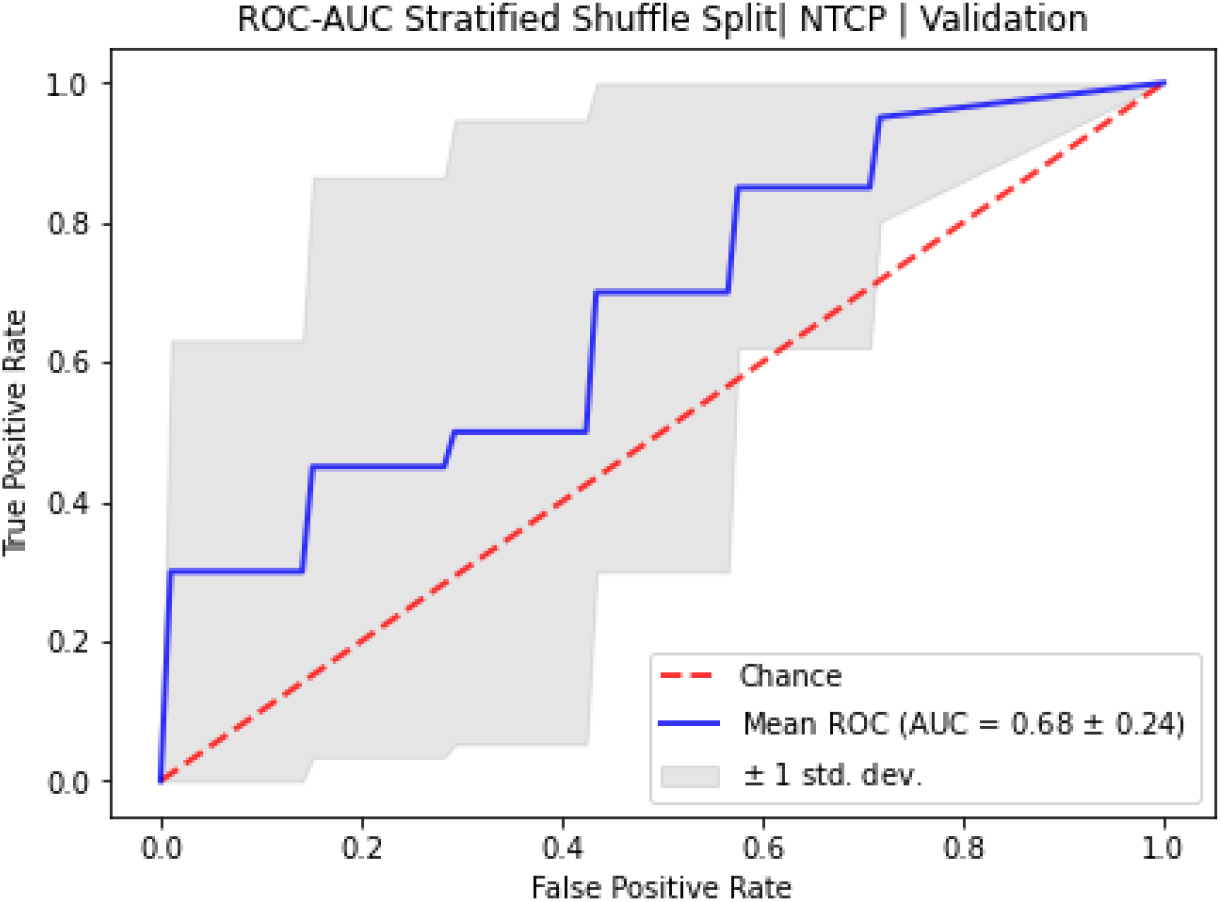
10-fold stratified shuffle 80-20 split ROC for HCC Non-Adaptive RTOE of NTCP modeled with GLoGD-GNN architecture.

#### S8.4 Synthetic Patient Generation via WGAN-GP

**Figure S26.**
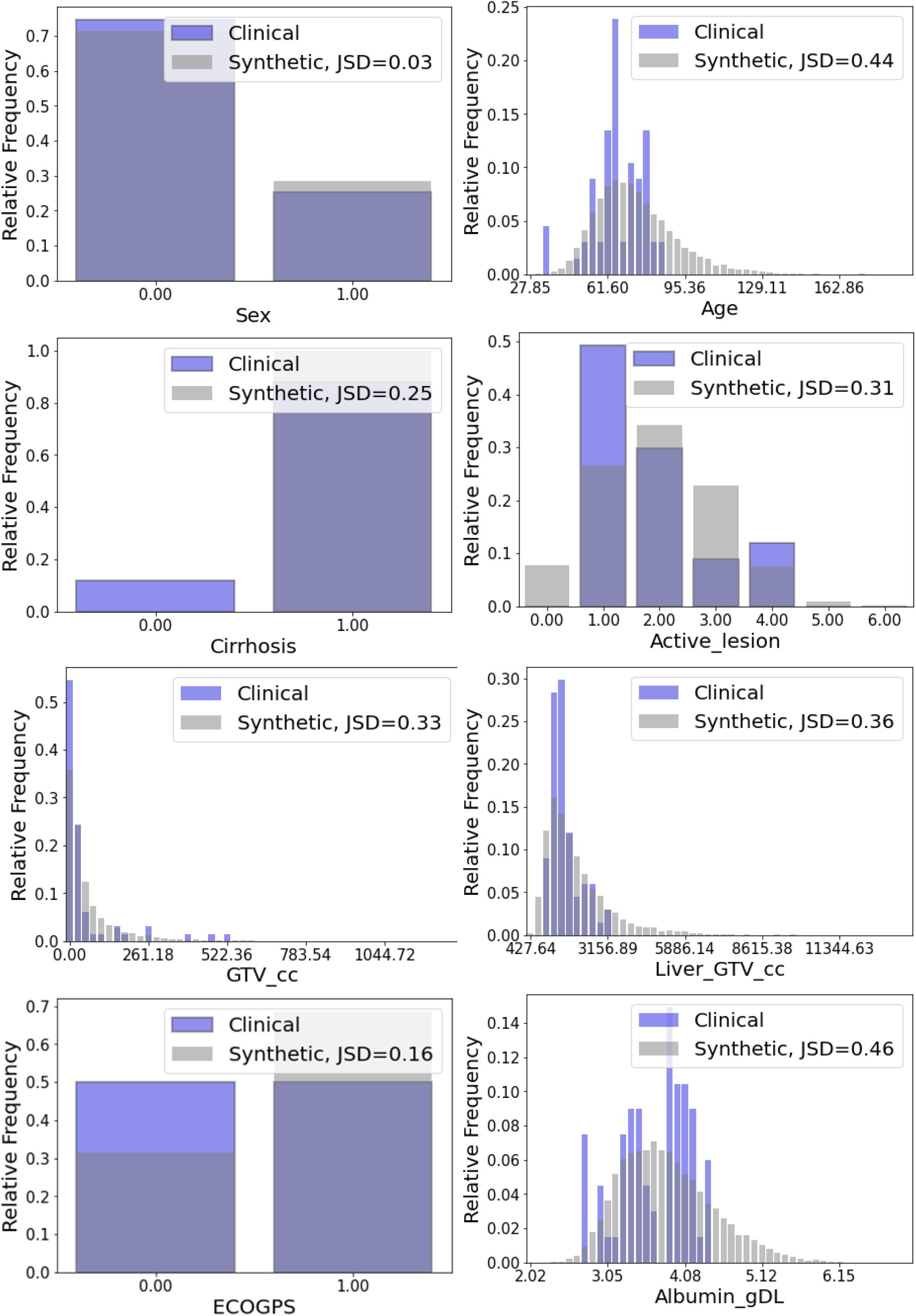
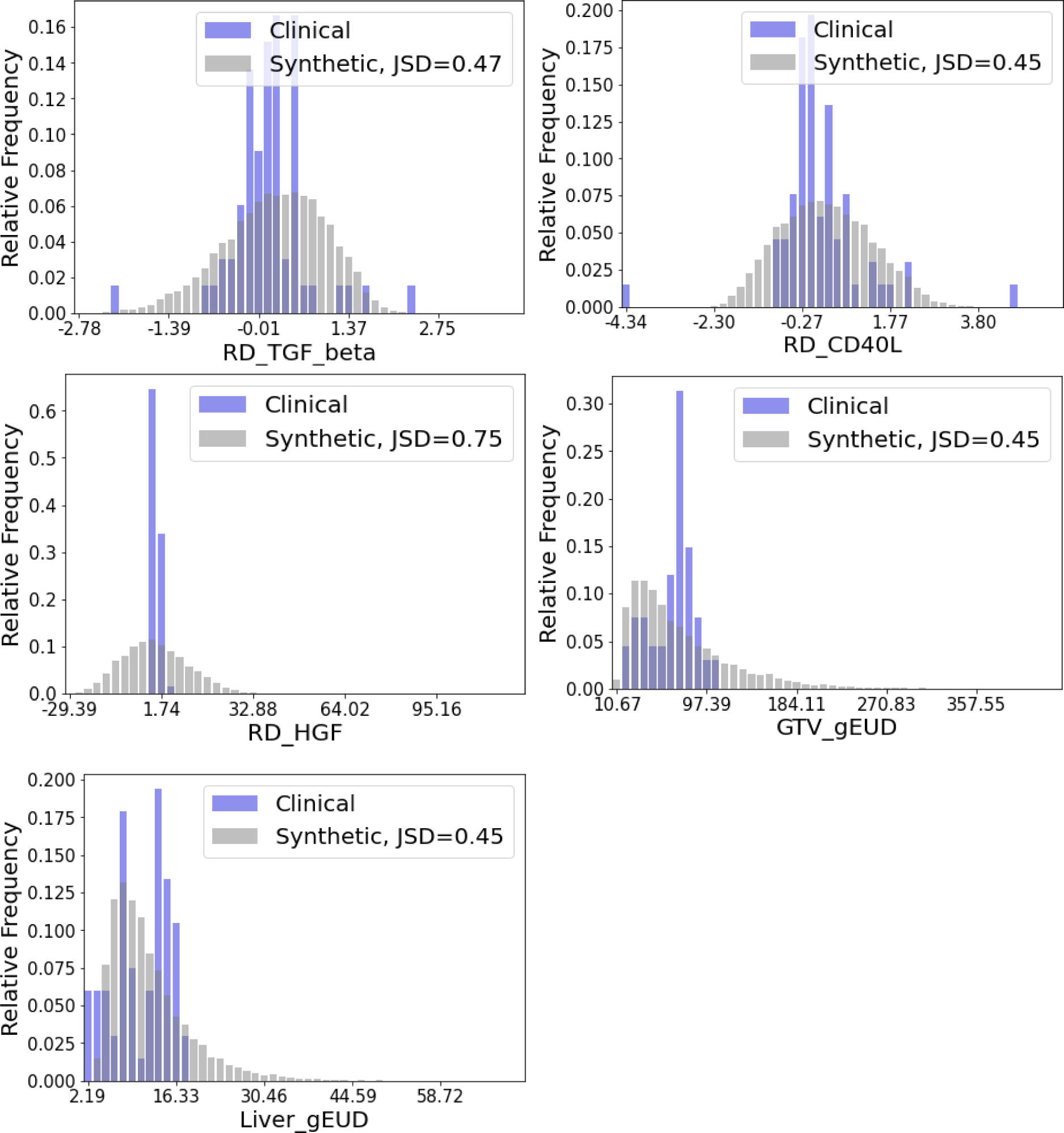
Distribution comparison of generated and original HCC dataset. Jensen Shannon Divergence metric between the distributions is provided for further insight on the differences.

#### S8.5 ODM Decision Analysis

**Table S16:** ODM Decision Analysis Results for HCC

| **Error**<br>**[Gy/frac]** | **Overall (n=64)** |  | **Positive Clinical Outcome**<br>**(n=54 (83%))** |  | **Negative Clinical Outcome (n=10 (15%))** |  |
| --- | --- | --- | --- | --- | --- | --- |
|  | RMSD | MAD | RMSD | MAD | RMSD | MAD |
| **Single GNN RTOE + DDQN ODM** | 4.75 ± 0.16 | 3.69 ± 0.16 | 4.25 ± 0.26 | 3.34 ± 0.22 | 6.78 ± 0.35 | 5.60 ± 0.38 |
| **GloGD GNN ROTE + DDQN ODM** | 2.96 ± 0.42 | 2.31 ± 0.47 | 2.79 ± 0.50 | 2.10 ± 0.54 | 4.02 ± 0.23 | 3.46 ± 0.17 |

| **Self-Evaluation**<br>**[Count (%)]** | **Overall (n=64)** |  |  | **Positive Clinical Outcome (n=54 (83%))** |  | **Negative Clinical Outcome (n=10 (15%))** |  |
| --- | --- | --- | --- | --- | --- | --- | --- |
|  | Good | Bad | Not Sure | Good | Not Sure | Good | Bad |
| **Single GNN RTOE + DDQN ODM** | 15 (23%) | 9 (14%) | 40 (62%) | 14 (26%) | 40 (74%) | 1 (10%) | 9 (90%) |
| **GloGD GNN ROTE + DDQN ODM** | 30 (46%) | 7 (11%) | 27 (42%) | 27 (50%) | 27 (50%) | 3 (30%) | 7 (70%) |

Error ± SEM | Error between average recommendation and clinical decisions| ensemble of 5 models

**Figure S27:**
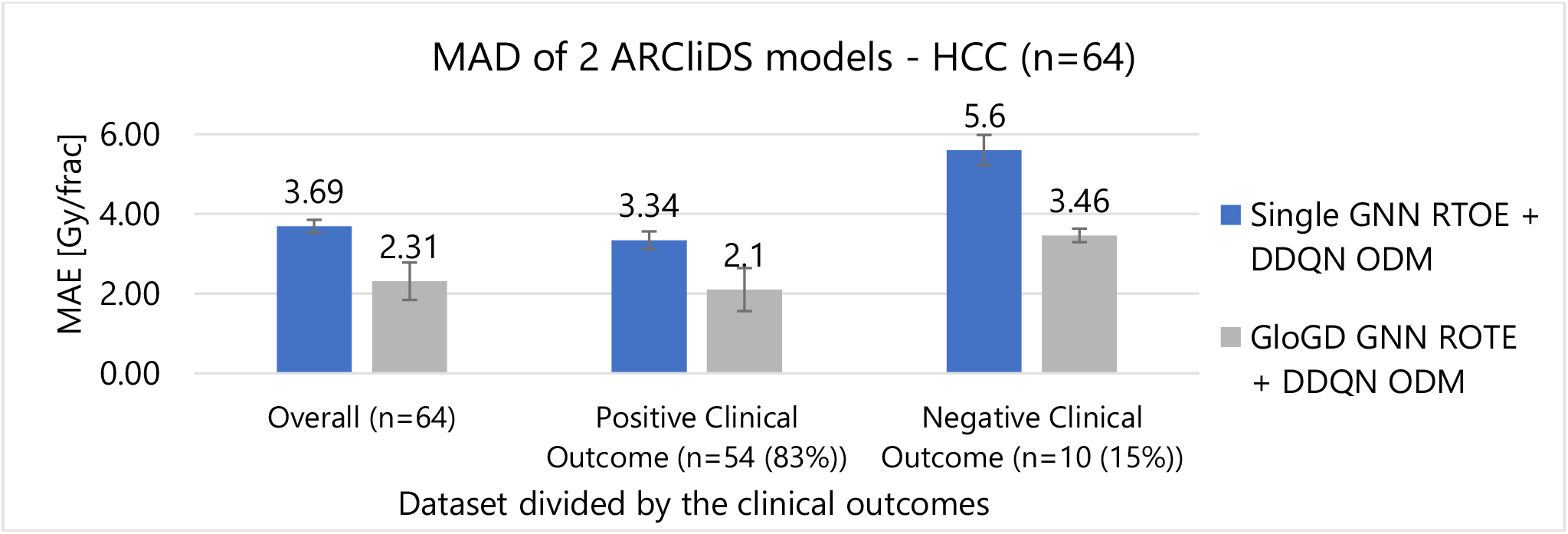
Mean Absolute Difference (MAD) of ARCliDS’s two model architecture for HCC patients grouped together according to the outcomes

##### S8.5.1 DDQN trained on Single GNN RTOE – HCC

**Figure S28:**
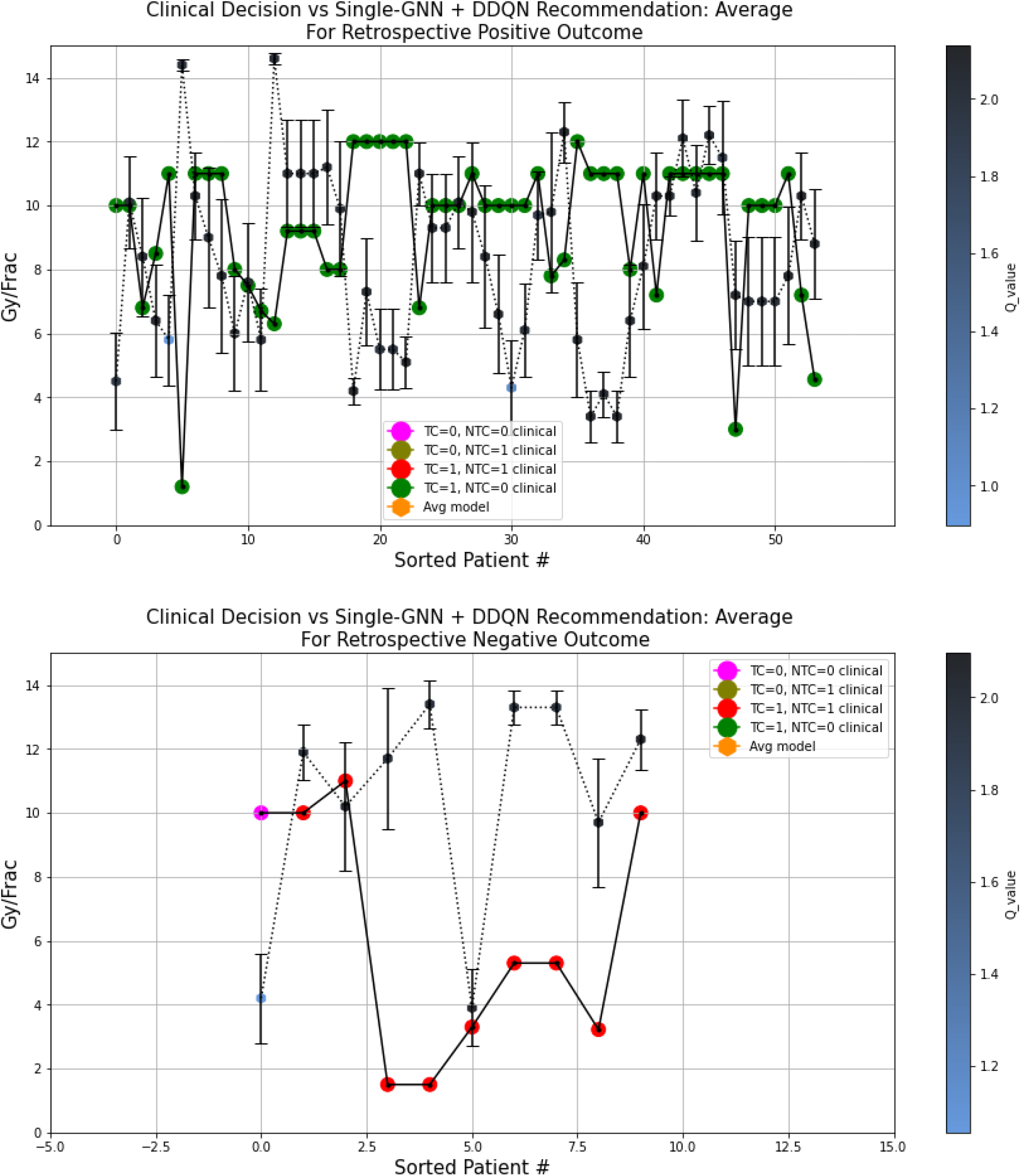
A visual comparison between the AI recommendation generated by the Single-GNN RTOE + DDQN ODM architecture and clinical decision for 2 groups of HCC patients divided according to the clinical outcomes. The clinical decisions are color coded with the outcomes and the ARCliDS recommendations are color coded with the respective q-values. Qualitatively, the q-value can be considered as the AI confidence in its recommendations.

##### S8.5.2 DDQN trained on GLoGD-GNN RTOE - HCC

**Figure S29:**
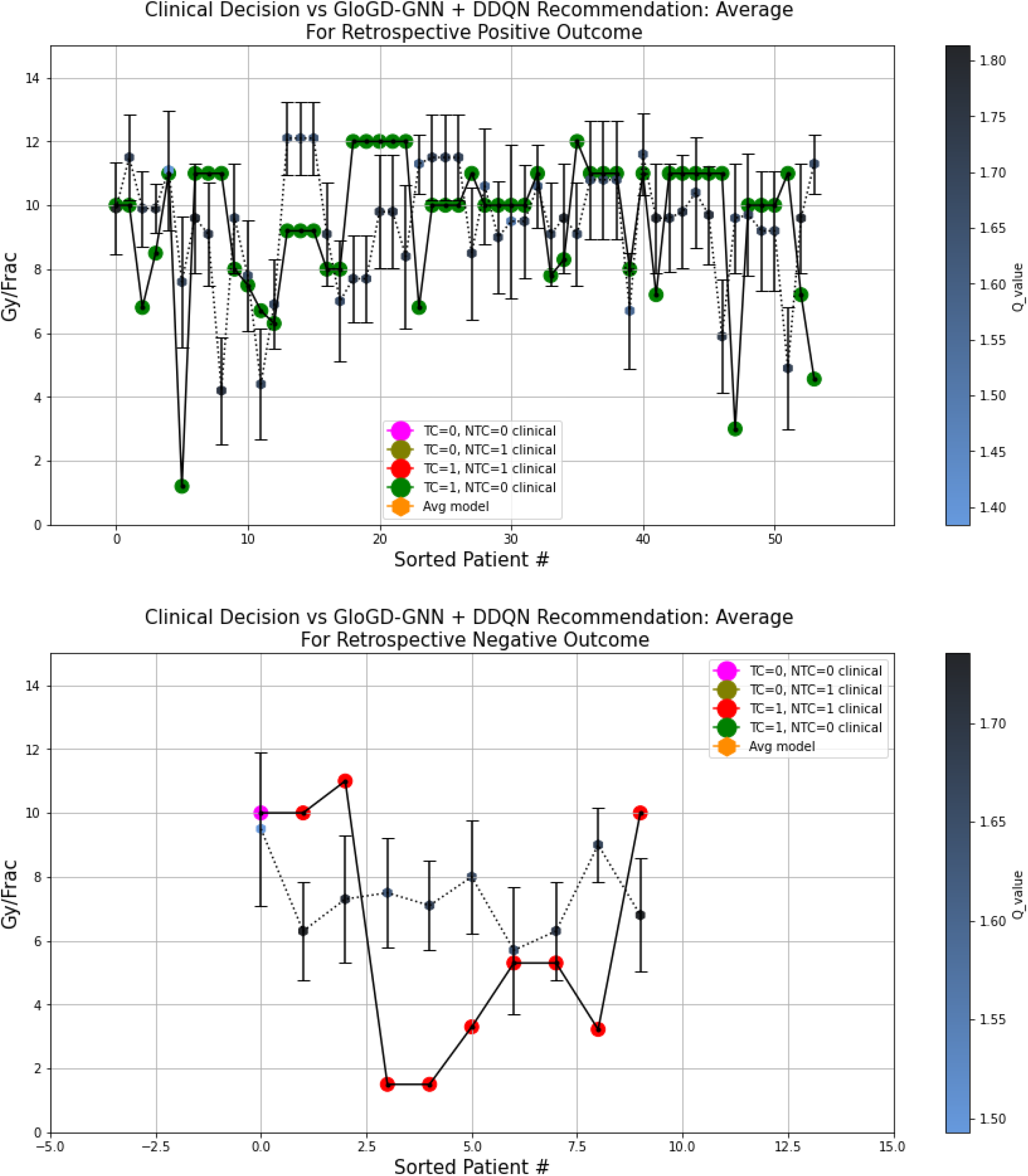
A visual comparison between the AI recommendation generated by the GloGD-GNN RTOE + DDQN ODM architecture and clinical decision for 2 groups of HCC patients divided according to the clinical outcomes. The clinical decisions are color coded with the outcomes and the ARCliDS recommendations are color coded with the respective q-values. Qualitatively, the q-value can be considered as the AI confidence in its recommendations.

##### S8.5.3 Experimentation with population-based reward goal for HCC

Due to data-related issues, as described in the Discussion Section S10, both TCP and NTCP response estimated by the GloGD-GNN ROTE were flatter than expected. Due to a flatter NTCP response, which did not span the whole probability space, the population-based reward goal severely limited the AI dose recommendation. In this section, we present plots for three different reward goals. As seen from the plots, for ntcp < 25%, the AI recommended lower dose values. The best recommendation corresponded to tcp > 90% and ntcp < 40%, however, the overall RMSD was lower than tcp > 50% and ntcp <50%.

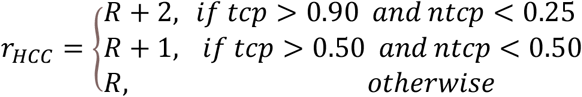

**Figure S30:**
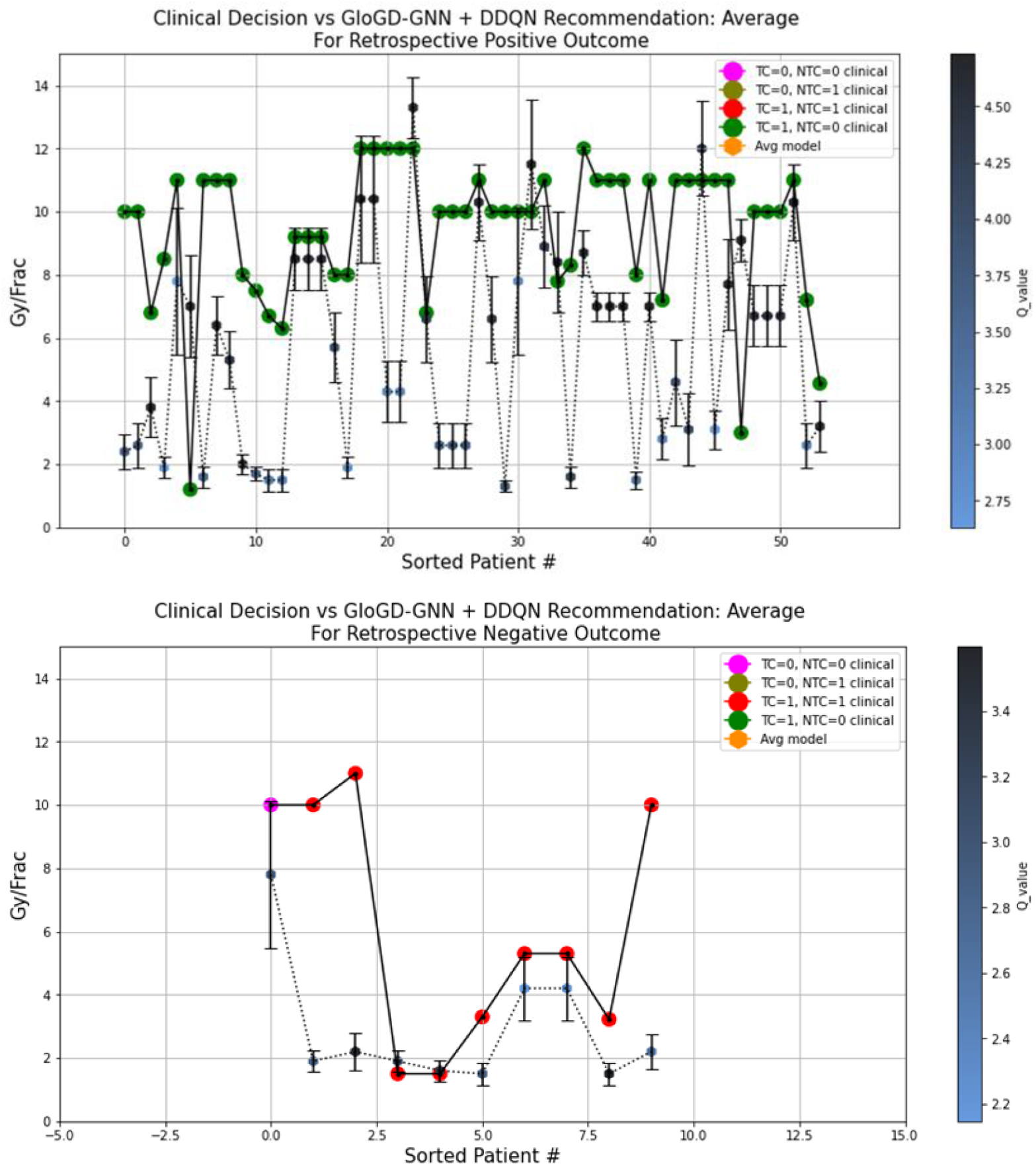
A visual comparison between the AI recommendation generated by the GloGD-GNN RTOE + DDQN ODM architecture and clinical decision for 2 groups of HCC patients divided according to the clinical outcomes. The ODM was trained with population-based reward goal of tcp > 90% and ntcp < 25%.

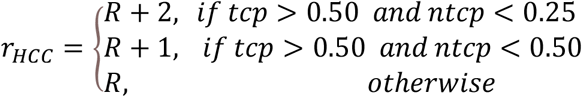

**Figure S31:**
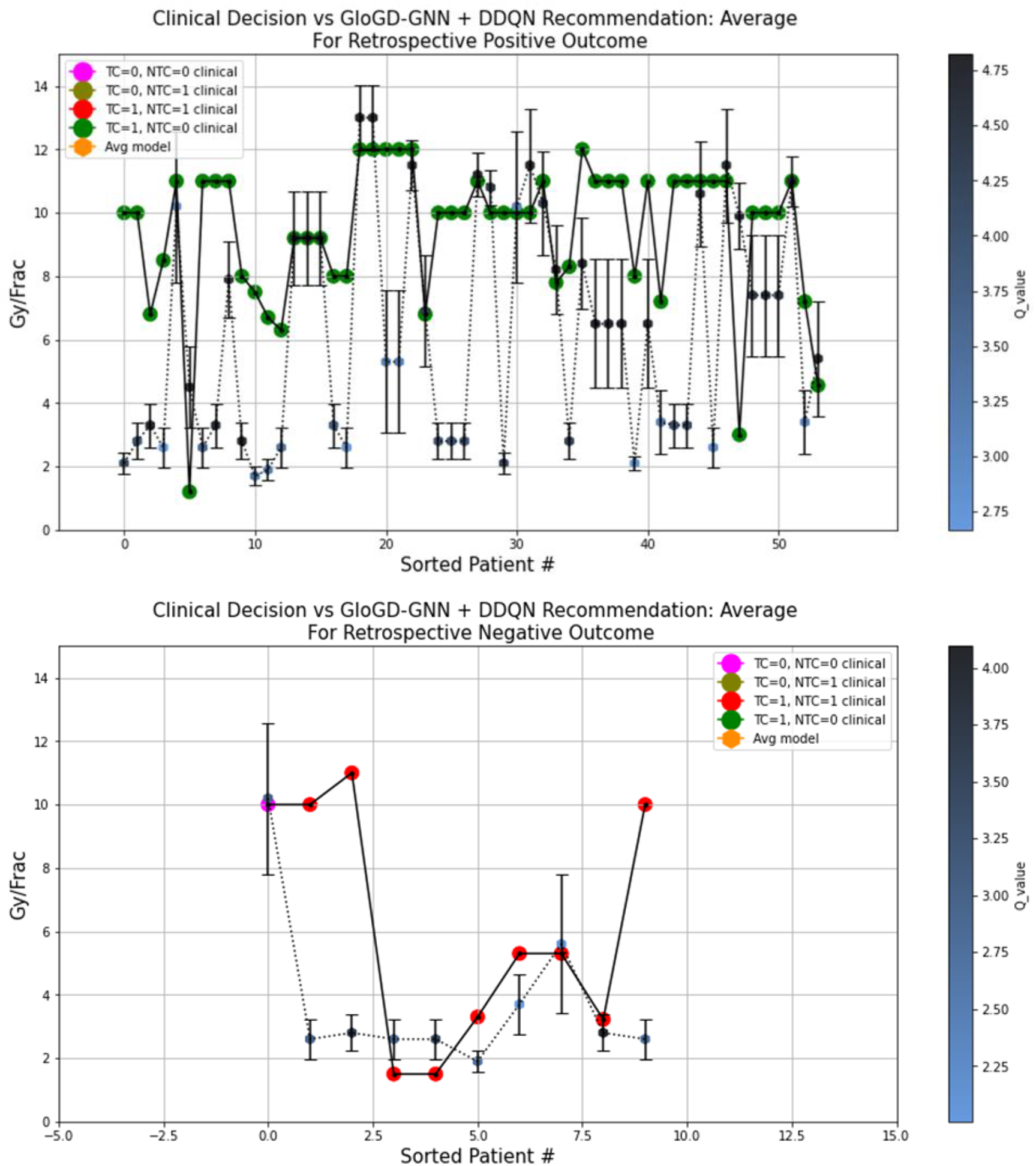
A visual comparison between the AI recommendation generated by the GloGD-GNN RTOE + DDQN ODM architecture and clinical decision for 2 groups of HCC patients divided according to the clinical outcomes. The ODM was trained with population-based reward goal of tcp > 50% and ntcp < 25%.

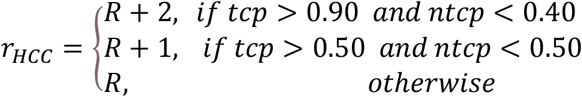

**Figure S32:**
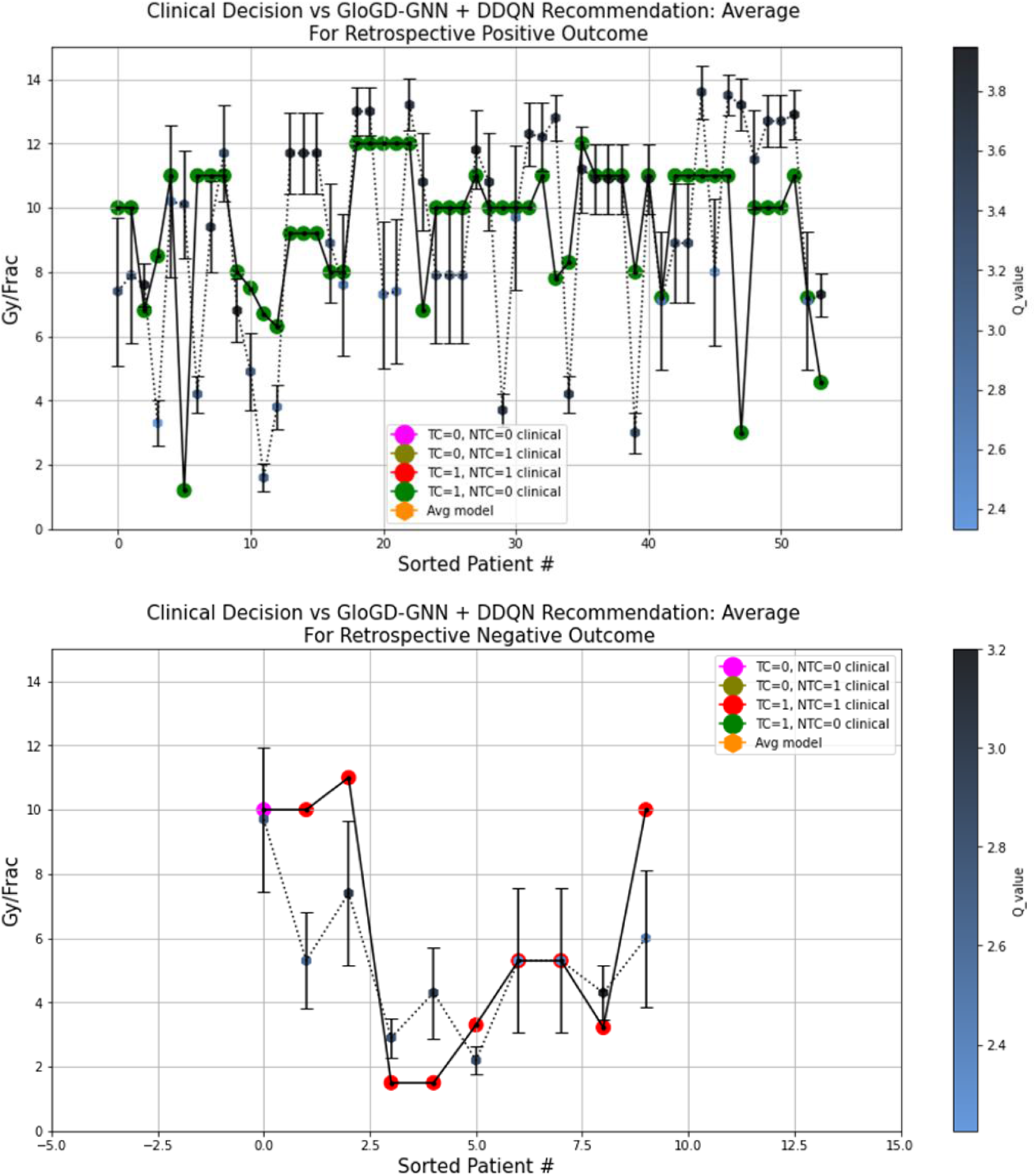
A visual comparison between the AI recommendation generated by the GloGD-GNN RTOE + DDQN ODM architecture and clinical decision for 2 groups of HCC patients divided according to the clinical outcomes. The ODM was trained with population-based reward goal of tcp > 90% and ntcp < 40%.

## References

1 Leech M, Katz MS, Kazmierska J, McCrossin J, Turner S. Empowering patients in decision-making in radiation oncology – can we do better? Mol Oncol 2020; 14: 1442–60.

2 Niraula D, Cui S, Pakela J, et al. Current status and future developments in predicting outcomes in radiation oncology. Br J Radiol 2022; published online July 28. DOI:10.1259/bjr.20220239.

3 Naqa I el, Kosorok MR, Jin J, Mierzwa M, ten Haken RK. Prospects and challenges for clinical decision support in the era of big data. JCO Clin Cancer Inform 2018; 2. DOI:10.1200/CCI.18.00002.

4 Tseng H-H, Luo Y, ten Haken RK, el Naqa I. The Role of Machine Learning in Knowledge-Based Response-Adapted Radiotherapy. Front Oncol 2018; 8. DOI:10.3389/fonc.2018.00266.

5 Niraula D, Jamaluddin J, Matuszak MM, Haken RK ten, Naqa I el. Quantum deep reinforcement learning for clinical decision support in oncology: application to adaptive radiotherapy. Sci Rep 2021; 11: 23545.

6 Sun W, Niraula D, el Naqa I, et al. Precision radiotherapy via information integration of expert human knowledge and AI recommendation to optimize clinical decision making. Comput Methods Programs Biomed 2022; 221: 106927.

7 Kosorok MR, Moodie EEM, editors. Adaptive Treatment Strategies in Practice. Philadelphia, PA: Society for Industrial and Applied Mathematics, 2015 DOI:10.1137/1.9781611974188.

8 Chakraborty B, Murphy SA. Dynamic Treatment Regimes. Annu Rev Stat Appl 2014; 1: 447–64.

9 el Naqa I, Kerns SL, Coates J, et al. Radiogenomics and radiotherapy response modeling. Phys Med Biol 2017; 62: R179–206.

10 el Naqa I, Pandey G, Aerts H, et al. Radiation Therapy Outcomes Models in the Era of Radiomics and Radiogenomics: Uncertainties and Validation. International Journal of Radiation Oncology*Biology*Physics 2018; 102: 1070–3.

11 Kamran SC, Mouw KW. Applying Precision Oncology Principles in Radiation Oncology. JCO Precis Oncol 2018; : 1–23.

12 Glide-Hurst CK, Lee P, Yock AD, et al. Adaptive Radiation Therapy (ART) Strategies and Technical Considerations: A State of the ART Review From NRG Oncology. International Journal of Radiation Oncology*Biology*Physics 2021; 109: 1054–75.

13 Kong F-M, ten Haken RK, Schipper M, et al. Effect of Midtreatment PET/CT-Adapted Radiation Therapy With Concurrent Chemotherapy in Patients With Locally Advanced Non–Small-Cell Lung Cancer. JAMA Oncol 2017; 3: 1358.

14 Jackson WC, Suresh K, Maurino C, et al. A mid-treatment break and reassessment maintains tumor control and reduces toxicity in patients with hepatocellular carcinoma treated with stereotactic body radiation therapy. Radiotherapy and Oncology 2019; 141: 101–7.

15 USFDA. Artificial Intelligence/Machine Learning (AI/ML)-Based Software as a Medical Device (SaMD) Action Plan. 2021 https://www.fda.gov/media/145022/download (accessed July 19, 2022).

16 USFDA. Proposed Regulatory Framework for Modifications to Artificial Intelligience/Machine Learning (AI/ML)-Based Software as a Medical Device(SaMD)). 2019 https://www.fda.gov/media/122535/download (accessed July 19, 2022).

17 Tseng H-H, Luo Y, Cui S, Chien J-T, ten Haken RK, Naqa I el. Deep reinforcement learning for automated radiation adaptation in lung cancer. Med Phys 2017; 44: 6690–705.

18 Goodfellow I, Bengio Y, Courville A. Deep Learning. MIT Press, 2016.

19 Hamilton WL. Graph Representation Learning. Morgan and Claypool, 2020.

20 Luo Y, McShan DL, Matuszak MM, et al. A multiobjective Bayesian networks approach for joint prediction of tumor local control and radiation pneumonitis in nonsmall-cell lung cancer (NSCLC) for response-adapted radiotherapy. Med Phys 2018; 45: 3980–95.

21 Sutton RS, Barto AG. Reinforcement Learning: An Introduction, 2nd edn. Cambridge: MIT Press, 2018.

22 Allen Li X, Alber M, Deasy JO, et al. The use and QA of biologically related models for treatment planning: Short report of the TG-166 of the therapy physics committee of the AAPM. Med Phys 2012; 39: 1386–409.

23 Issam El Naqa, editor. A Guide to Outcome Modeling In Radiotherapy and Oncology : Listening to the Data, 1st edn. Boca Raton : CRC Press, 2018.

24 Hasselt H van, Guez A, Silver D. Deep Reinforcement Learning with Double Q-Learning. In: Proceedings of the Thirtieth AAAI Conference on Artificial Intelligence. AAAI Press, 2016: 2094–100.

25 Roberts DA, Yaida S, Hanin B. The Principles of Deep Learning Theory. Cambridge University Press, 2022.

26 Gulrajani I, Ahmed F, Arjovsky M, Dumoulin V, Courville A. Improved Training of Wasserstein GANs. 2017; published online March 31.

27 Sousa F, Jourani Y, Dragan T, Beauvois S, Somoano M, van Geste D. Re-planning assessment in head and neck cancer radiotherapy: 3 years single institution experience. In: Re-planning assessment in head and neck cancer radiotherapy: 3 years single institution experience. Copenhagen: ESTRO 2022, 2022.

28 Gros SAA, Santhanam AP, Block AM, Emami B, Lee BH, Joyce C. Retrospective Clinical Evaluation of a Decision-Support Software for Adaptive Radiotherapy of Head and Neck Cancer Patients. Front Oncol 2022; 12. DOI:10.3389/fonc.2022.777793.

29 Nass SJ, Levit LA, Gostin LO, editors. Beyond the HIPAA Privacy Rule. Washington, D.C.: National Academies Press, 2009 DOI:10.17226/12458.

30 Topaloglu MY, Morrell EM, Rajendran S, Topaloglu U. In the Pursuit of Privacy: The Promises and Predicaments of Federated Learning in Healthcare. Front Artif Intell 2021; 4. DOI:10.3389/frai.2021.746497.

31 Mirza M, Osindero S. Conditional Generative Adversarial Nets. 2014; published online Nov 6.

## References

1 Luo Y, McShan DL, Matuszak MM, et al. A multiobjective Bayesian networks approach for joint prediction of tumor local control and radiation pneumonitis in nonsmall-cell lung cancer (NSCLC) for response-adapted radiotherapy. Med Phys 2018; 45: 3980–95.

2 Hamilton WL. Graph Representation Learning. Morgan and Claypool, 2020.

3 Issam El Naqa, editor. A Guide to Outcome Modeling In Radiotherapy and Oncology : Listening to the Data, 1st edn. Boca Raton : CRC Press, 2018.

4 Gulrajani I, Ahmed F, Arjovsky M, Dumoulin V, Courville A. Improved Training of Wasserstein GANs. 2017; published online March 31.

6 Ramírez MF, Huitink JM, Cata JP. Perioperative Clinical Interventions That Modify the Immune Response in Cancer Patients. Open J Anesthesiol 2013; 03: 133–9.

7 Schaue D, Kachikwu EL, McBride WH. Cytokines in Radiobiological Responses: A Review. Radiat Res 2012; 178: 505–23.

8 Warltier DC, Laffey JG, Boylan JF, Cheng DCH. The Systemic Inflammatory Response to Cardiac Surgery. Anesthesiology 2002; 97: 215–52.

9 Mahasittiwat P, Yuan S, Xie C, et al. Metabolic tumor volume on PET reduced more than gross tumor volume on CT during radiotherapy in patients with non-small cell lung cancer treated with 3DCRT or SBRT. J Radiat Oncol 2013; 2: 191–202.

10 Carrier-Vallieres M. Radiomics: enabling factors towards precision medicine. 2018.

11 Issam El Naqa, editor. A Guide to Outcome Modeling In Radiotherapy and Oncology : Listening to the Data, 1st edn. Boca Raton : CRC Press, 2018.

12 Hildebrandt MAT, Komaki R, Liao Z, et al. Genetic Variants in Inflammation-Related Genes Are Associated with Radiation-Induced Toxicity Following Treatment for Non-Small Cell Lung Cancer. PLoS One 2010; 5: e12402.

13 Chang JS, Wrensch MR, Hansen HM, et al. Nucleotide excision repair genes and risk of lung cancer among San Francisco Bay Area Latinos and African Americans. Int J Cancer 2008; 123: 2095–104.

14 Kiyohara C, Yoshimasu K. Genetic polymorphisms in the nucleotide excision repair pathway and lung cancer risk: A meta-analysis. Int J Med Sci 2007; : 59–71.

15 Nagpal N, Kulshreshtha R. miR-191: an emerging player in disease biology. Front Genet 2014; 5. DOI:10.3389/fgene.2014.00099.

16 Ricciuti B, Mecca C, Crinò L, Baglivo S, Cenci M, Metro G. Non-coding RNAs in lung cancer. Oncoscience 2014; 1: 674–705.

17 Lin S, Gregory RI. MicroRNA biogenesis pathways in cancer. Nat Rev Cancer 2015; 15: 321–33.

18 Caraceni P, Tufoni M, Bonavita ME. Clinical use of albumin. Blood Transfus 2013; 11 Suppl 4: s18–25.

19 Morikawa M, Derynck R, Miyazono K. TGF-β and the TGF-β Family: Context-Dependent Roles in Cell and Tissue Physiology. Cold Spring Harb Perspect Biol 2016; 8. DOI:10.1101/cshperspect.a021873.

20 Elgueta R, Benson MJ, de Vries VC, Wasiuk A, Guo Y, Noelle RJ. Molecular mechanism and function of CD40/CD40L engagement in the immune system. Immunol Rev 2009; 229: 152–72.

21 Perreau M, Suffiotti M, Marques-Vidal P, et al. The cytokines HGF and CXCL13 predict the severity and the mortality in COVID-19 patients. Nat Commun 2021; 12: 4888.

22 Matsumoto K, Nakamura T. Roles of HGF as a pleiotropic factor in organ regeneration. EXS 1993; 65: 225–49.

